# Genomic and epidemiological analysis of SARS-CoV-2 viruses in Sri Lanka

**DOI:** 10.1101/2021.05.05.21256384

**Authors:** Chandima Jeewandara, Deshni Jayathilaka, Diyanath Ranasinghe, Nienyun Sharon Hsu, Dinuka Ariyaratne, Tibutius Thanesh Jayadas, Deshan Madushanka, Benjamin B. Lindsey, Laksiri Gomes, Matthew D. Parker, Ananda Wijewickrama, Malika Karunaratne, Graham S. Ogg, Thushan I. de Silva, Gathsaurie Neelika Malavige

## Abstract

**Background:** In order to understand the molecular epidemiology of SARS-CoV-2 in Sri Lanka, since March 2020, we carried out genomic sequencing overlaid on available epidemiological data until April 2021.

**Methods:** Whole genome sequencing was carried out on diagnostic sputum or nasopharyngeal swabs from 373 patients with COVID-19. Molecular clock phylogenetic analysis was undertaken to further explore dominant lineages.

**Results:** The B.1.411 lineage was most prevalent, which was established in Sri Lanka and caused outbreaks throughout the country until March 2021. The estimated time of the most recent common ancestor of this lineage was 29^th^ June 2020 (95% lower and upper bounds 23rd May to 30^th^ July), suggesting cryptic transmission may have occurred, prior to a large epidemic starting in October 2020. Returning travellers were identified with infections caused by lineage B.1.258, as well as the more transmissible B.1.1.7 lineage, which has replaced B.1.411 to fuel the ongoing large outbreak in the country.

**Conclusions:** The large outbreak that started in early October, is due to spread of a single virus lineage, B.1.411 until the end of March 2021, when B.1.1.7 emerged and became the dominant lineage.

## Introduction

The severe acute respiratory syndrome coronavirus-2 (SARS-CoV-2) has emerged as the leading cause of mortality in several countries in the world. As of the 24^th^ of May 2021, 167 million cases and 3.45 million deaths have been reported worldwide [1]. Due to the emergence of variants of concern, the World Health Organization has recommended whole genomic sequencing of the SARS-CoV-2 viruses within countries regularly and systematically for early identification of such variants [2].

The first patient infected with SARS-CoV-2 in Sri Lanka was reported on the 27^th^ January 2020, who was a foreign national, with the first Sri Lankan patient reported on the 10^th^ of March 2020 [3]. In the following six months (March to September), the spread of the virus was largely contained with only 3111 reported cases, of which 38.8% were imported [4]. However, there was a surge in the number of cases with discovery of a new cluster in early October 2020 in a clothing factory in the district adjacent to Colombo (Gampaha). This was followed by rapid spread of SARS-CoV-2 within the Colombo Municipality region (CMC), fish markets and subsequently to the whole country. This outbreak continued to evolve, with infections being reported in all regions of the country, but the numbers gradually declined by end of March 2021, with the number of daily cases been reported falling to 200 to 300 cases per day[5]. The number of daily cases remained <250 cases/day until mild-April 2021, when a gradual and steep rise in the number of cases was seen with daily infections rising over 1000/day by the last week of April to over 2500/day by mid-May [6].

We carried out SARS-CoV-2 sequencing from isolates collected throughout the different phases of the pandemic in order to determine the molecular epidemiology of SARS-CoV-2 in Sri Lanka, including current circulation of viruses with mutations that may confer greater transmissibility and/or threaten the efficacy of vaccines.

## Methods

### Patients

Whole genome sequencing was carried out on diagnostic sputum or nasopharyngeal swabs from 373 patients with COVID-19, where cycle threshold (Ct) values of the quantitative SARS-CoV-2 real-time PCR was <30. Ethical approval for the study was obtained by the Ethics Review Committee of the University of Sri Jayewardenepura.

### Viral RNA Extraction

Viral RNA was extracted using QIAamp viral RNA mini kit (Qiagen, USA), SpinStarTM Viral Nucleic Acid Extraction kit 1.0 (ADT Biotech, Malaysia) or FastGene RNA Viral Kit, (Nippon Genetics, Germany) according to manufacturer’s instructions. Presence of ORF1ab gene and N gene of SARS-CoV-2 was detected with Novel Coronavirus (2019-Ncov) Nucleic Acid Diagnostic Kit (Sansure Biotec) and S gene was detected by Taqpath COVID-19 RT PCR kit (Applied biosystems) by real time RT PCR in ABI 7500 real time PCR system (Applied Biosystems, USA).

### Library Preparation and Next Generation sequencing (NGS)

Library preparation was attempted using either the TruSeq Stranded Total RNA Library Prep Gold (Illumina, San Diego, USA) or the The AmpliSeq for Illumina SARS-CoV-2 Community Panel, in combination with AmpliSeq for Illumina library prep, index, and accessories (Illumina, San Diego, USA). Shotgun metagenomic sequencing workflow was used for four initial samples, while for the remainder (n= 369) a targeted RNA/cDNA amplicon assay was used (Supplementary methods).

### Phylogenetic Analysis of SARS-CoV-2 sequences

Sequences with ≤10x median read depth and ≤86% genome coverage were taken forward for further analysis. GISAID accession numbers, location, and the clades of sequences used are shown in Supplementary table 1. 373 sequences from Sri Lanka were combined with 3200 representative global sequences. The representative sample of global sequences was obtained in two steps using all available data on GISAID up until 12^th^ May 2021. The first step included randomly selecting one sequence per country per epi week. This was then followed by a random sampling of the remaining sequences to generate a sample of 3200 sequences. All sequences were then aligned to the SARS-CoV-2 reference strain MN908947.3 using MAFFT version 7.477 (Katoh_2002). We masked alignment positions that have been previously flagged as problematic (https://github.com/W-L/ProblematicSites_SARS-CoV-2) and manually removed obvious sequencing errors and potential homoplasic positions. A maximum likelihood tree was constructed using IQ-TREE2 version 2.1.2 (Minh_2020) with the GTR+G model of nucleotide substitution (Tavaré_1986, Yang_1994) and 1000 replicates of ultrafast bootstrapping (-B 1000) and SH-aLRT branch test (-alrt 1000).

Molecular clock phylogenetic analysis was undertaken using sequences from Sri Lanka. The alignment and maximum likelihood tree construction were performed using MAFFT and IQ-TREE2 as described above. TreeTime [7] was used to infer a molecular clock phylogeny using a strict evolutionary rate of 1.1 × 10−3 substitutions/site/year (estimated by Duchene et al.[8]) and a standard deviation of 0.00004. The tree was rerooted with least-squares criteria in TreeTime. Eight samples were excluded from the analysis due to their inconsistent temporal signal. Lineages were assigned using Pangolin (version v2.4.2, lineages version 2021-04-28). Phylogenetic tree visualizations were produced using R (v3.5.3), R/ape, R/ggtree, R/ggplot2, R/ggtreeExtra, R/dplyr, R/phytools, R/tidytree. Two proportional symbol maps of Sri Lanka were plotted with GPS coordinates of the sampling locations of B.1.411 and B.1.17 sequences using R (v4.0.1), R/maps, R/ggplot2, R/ggrepel, R/cowplot and R/dplyr. Each sampling location was indicated by a coloured bubble proportionate to the number of sequences sampled within. Colombo district was zoomed into a sub map (longitude: 79.80 −79.98, latitude: 6.80 - 6.98) in order to visualize the suburbs as Colombo had the highest sampling density.

## Results

Of six samples collected in March 2020 from returning travellers and their contacts (period A, figure 1: four from Colombo district, two from Kalutara district), two belonged to lineage B.4, two to B 1.1, one to B.1 and one to B (Figure 2 and Supplementary table 1). During early April, SARS-CoV-2 spread within closed community clusters in the CMC region (period B, Figure 1). Two viruses from these clusters belonged to lineages B.4 and B.1 (Supplementary table 1). Period C in figure 1 was thought to be due to an outbreak initiated by the returning workforce from the Middle East. A sequence was obtained from only one virus, which belonged to lineage B. Again, due to detection of infected patients at the airport and mandatory quarantine of all individuals for at least 14 days, during May to June, cases appeared not to spill over to the community. However, there was a sudden surge in the number of cases in mid-July in a drug rehabilitation centre (DRC), in the North Central Province (period D, figure 1). The origin of this outbreak was not known and three lineage B.1 sequences obtained, formed a cluster separate from earlier B.1 sequences from Sri Lanka. This outbreak was also subsequently controlled.

**Figure 1:**
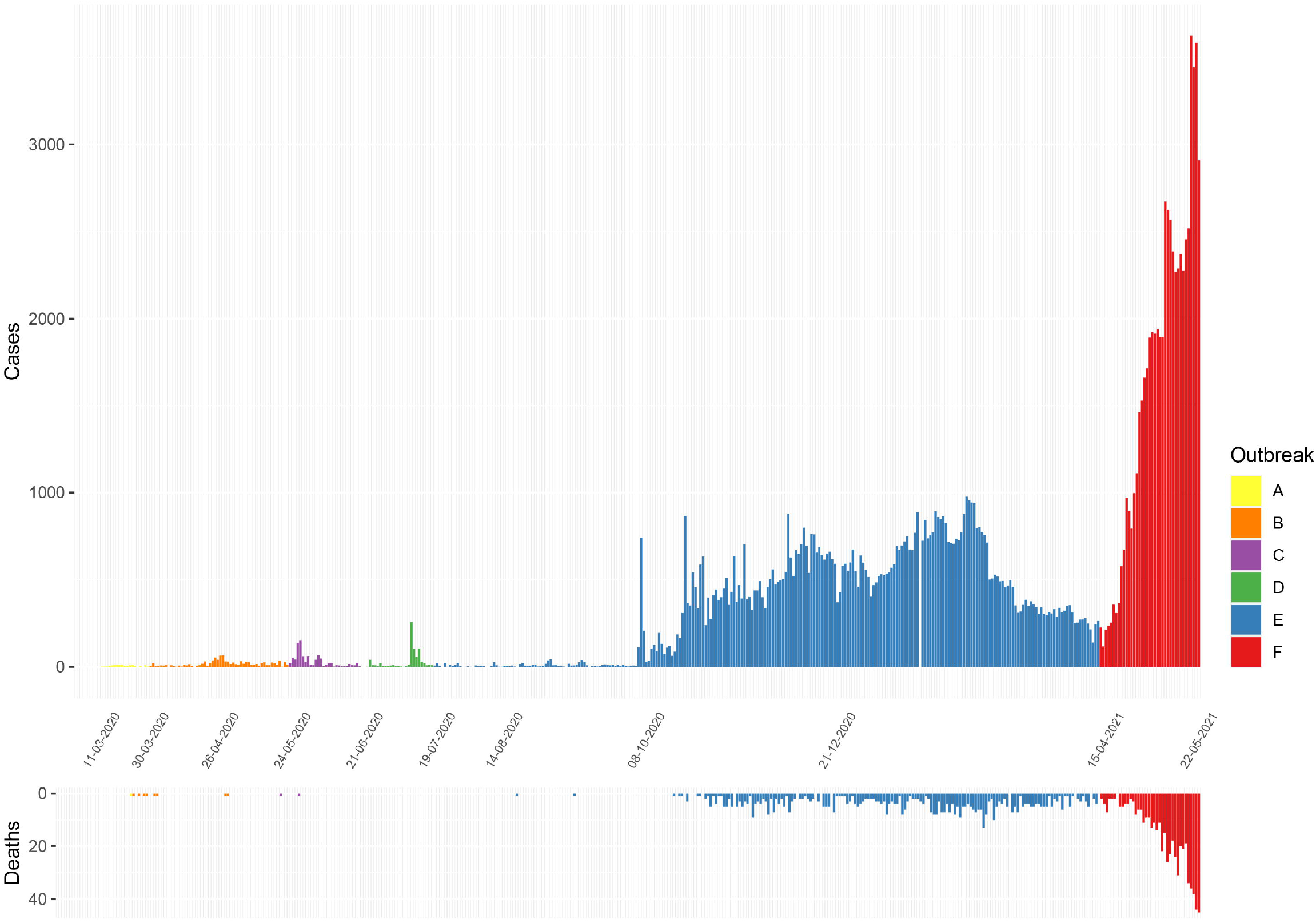
Epidemiological curves of COVID-19 cases and deaths reported in Sri Lanka from 11^th^ March 2020 to 22^nd^ May 2021. (Data from Epidemiology Unit, Ministry of Health, Sri Lanka)[3] A-initial outbreak from overseas returned, B-closed outbreak in a the Colombo Municipality area, C-returnees from Middle East, D-outbreak at the drug rehabilitation centre, E-the large outbreak that initially began in a clothing factory, F-the current ongoing outbreak.

**Figure 2.**
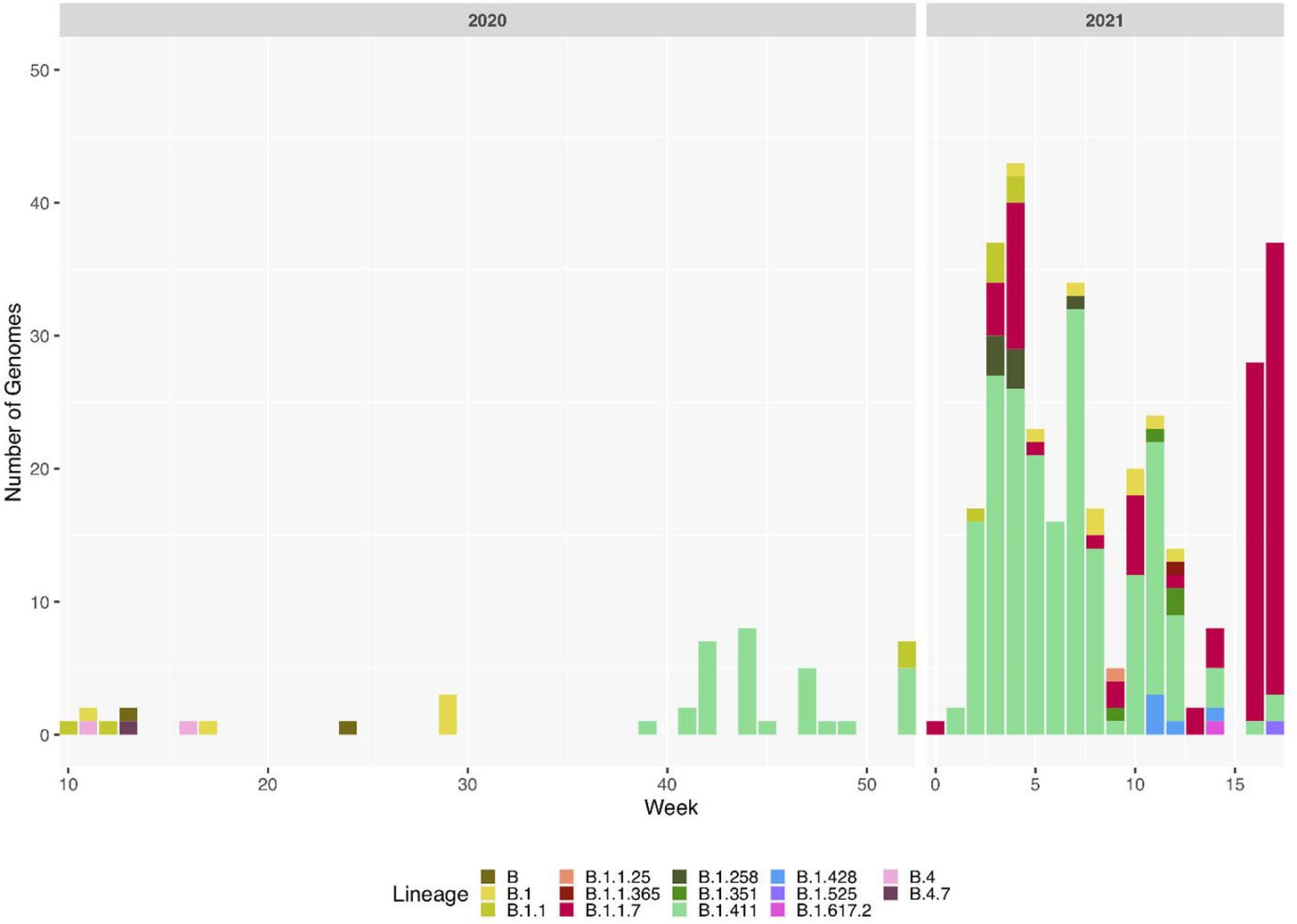
Sri Lanka sequence lineage distribution since the detection of SARS-CoV-2 in Sri Lanka. Lineage breakdown of 373 Sri Lanka samples across time. B.1.411 was first detected in week 39 in 2020 and since B.1.411 has dominated the outbreak in Sri Lanka until week 12, 2021. B.1.1.7 was detected at the beginning of 2021 and its prevalence has rapidly increased in Sri Lanka.

Sri Lanka then had a period of approximately two months during August and September, where no locally acquired infections were reported [3]. In early October, an outbreak occurred in a clothing factory that heralded a wave of infections, which continued to the end of March 2021. This outbreak in early October was soon followed by widespread transmission in many factories, fish markets island wide, in the highly populated CMC region in Colombo and subsequently to many areas in the country (period E, figure 1). Following a period of intense spread in the CMC for 4 months, the number of cases gradually declined from January 2021 to early April 2021. However, a rapid rise in the number of cases were seen from mid-April 2021 (period F, Figure 1), with the numbers exponentially increasing as of 24^th^ May 2021.

### Outbreak with the B.1.411 lineage

We sequenced 361 samples taken during October 2020 to 30^th^ April 2021, to determine how the outbreak evolved and establish if there were any new introductions to fuel ongoing infections. 231 of these 361 viruses were classified into a novel lineage B.1.411, which appears to have arisen in Sri Lanka and was responsible for the outbreak of SARS-CoV-2 in Sri Lanka from October 2020 to early April 2021 (Figure 2 and 3). Based on TimeTree analysis (Figure 4), the estimated time of the most recent common ancestor (tMRCA) of the B.1.411 lineage emerged around the 29^th^ of June 2020 (95% lower and upper bounds 23rd May to 30th July). The distribution of B.1.411 cases throughout the country is shown in Figure 5.

**Figure 3.**
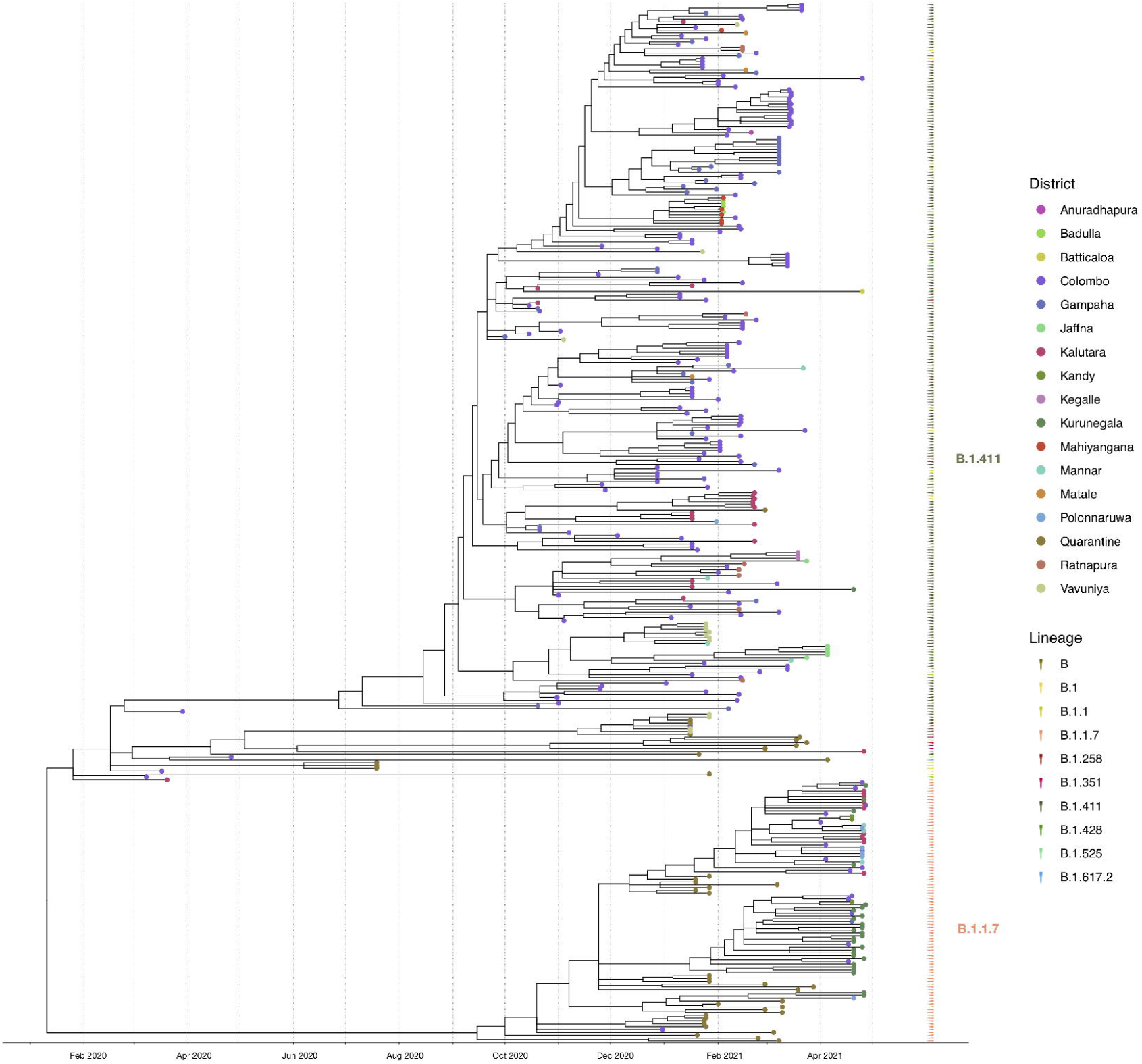
Phylogenetic analysis of Sri Lankan sequences. Maximum likelihood tree of 373 Sri Lanka samples and a representative sample of global sequences obtained from GISAID. Tips are coloured by Pango lineages, and the external layer on the right indicates the outbreak period associated with each Sri Lanka sample. The tree was estimated using IQTree2 (GTR maximum likelihood model and +G heterogeneity rate).

**Figure 4.**
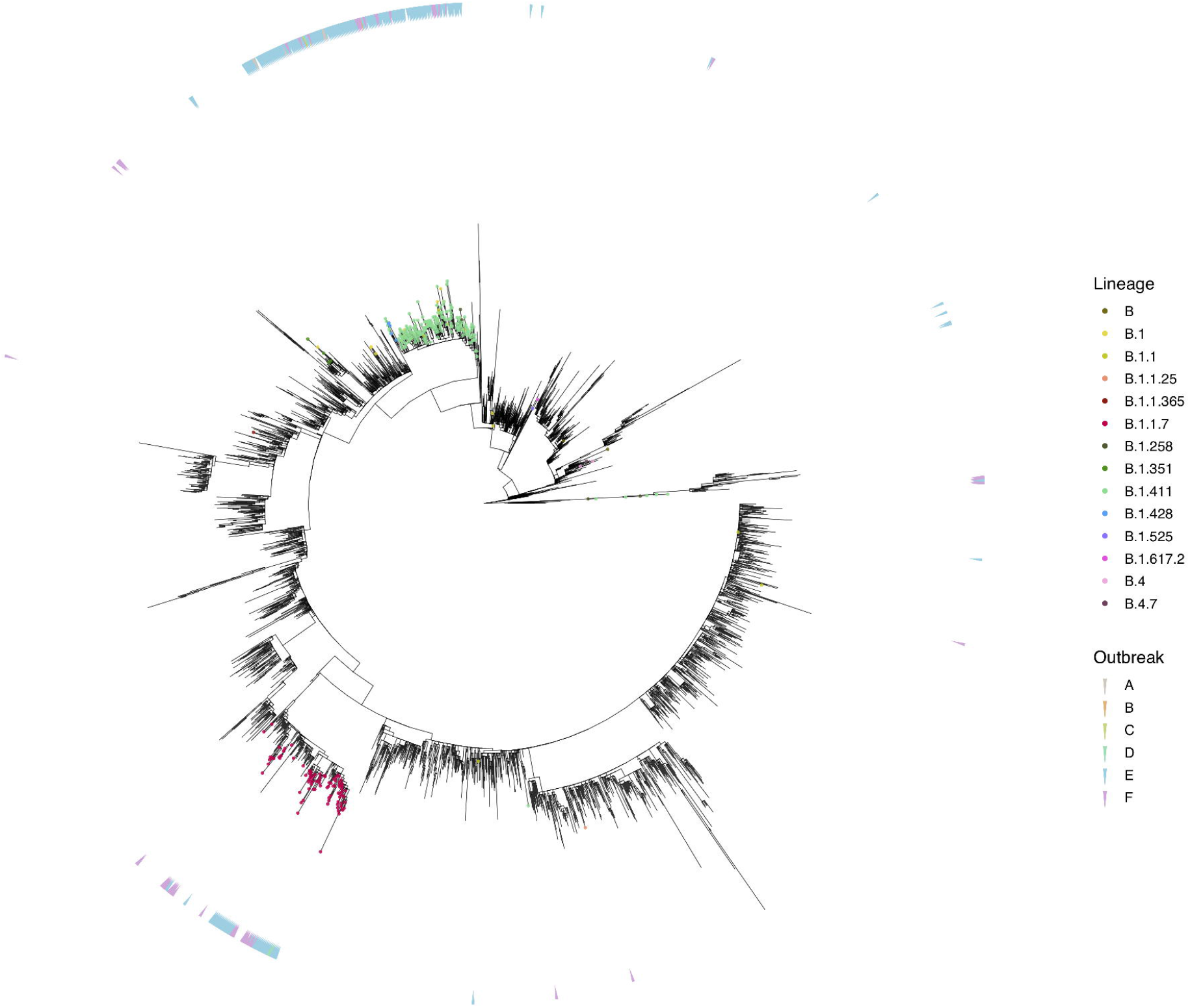
Molecular clock phylogeny of Sri Lankan samples. Tips are coloured by the sample location, and the external layer on the right shows the sample lineages. The molecular clock was inferred using TreeTime with an evolutionary rate of 1.1 × 10−3 substitutions/site/year and a standard deviation of 0.00004.

**Figure 5.**
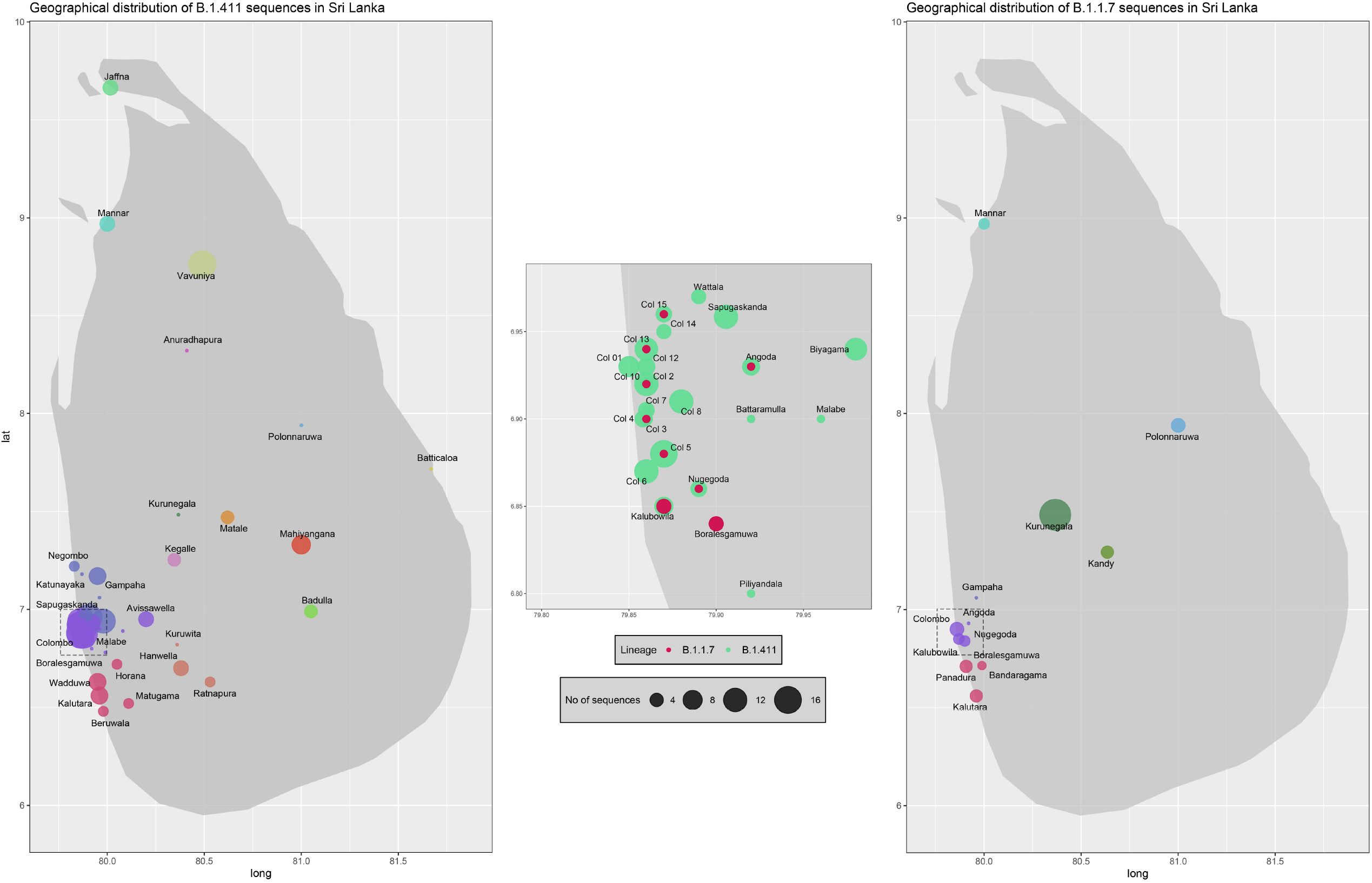
Geographical distribution of SARS-CoV-2 B.1.411 and B.1.17 lineage infections in Sri Lanka sampled from October 2020 to April 2021. Radius of each bubble accounts for the number of sequences reported from its representing district and the colour code based on the location is adopted from the figure 4. Colombo district is expanded in the middle to visualize its suburbs.

NSP12 was the most mutated region in B.1.411 lineage followed by the spike protein and NSP2. The predominant mutations in the spike protein were D614G seen in 228/231 sequences andH1159Y mutation seen in 215/231 sequences. M666I mutation was the second most abundant mutation in B.1.411 lineage with 224 occurrences in the NSP12 region (Supplementary table 2). Importantly, one B.1.411 genome sequenced in mid-February had the E484K mutation while four sequences from quarantine centres had acquired N439K in the spike protein along with the other B.1.411 lineage defining mutations. Also, we observed that the ΔH69/V70 deletion in the spike had co-occurred in three of those with the N439K mutation, which were identified from individuals in quarantine facilities. The other mutations seen in B.1.411 lineage are, T116I in NSP2, L37F in NSP6, P323L and M666I in NSP12 and T205I in the N protein (Supplementary table 2). A phylogenetic tree of all sequences in our analysis is shown in Figure 2.

### Outbreak with the B.1.1.7 lineage

Twenty seven infections sampled between January and late March 2021 belonged to the B.1.1.7 lineage. The first case of these was an imported infection on the 02^nd^ of January 2021, with a further 26 imported cases detected between January and March 2021. Except one case infection that was seen in the community and another infection seen in a fisherman in March 2021, all other cases were identified within quarantine facilities during this period (Figure 2). After a gradual decline in the number of cases by the end of March, a rapid surge in the number of cases were seen since mid-April 2021. From the 78 samples sequenced from 26^th^ March to 30^th^ April, 66 were found to be of the B.1.1.7 variant (Figure 2 and 4). By end of April, B.1.1.7 was the predominant variant circulating in many parts of the country (Figure 5).

### Other SARS-CoV-2 variants detected

Twenty infections sampled from the community and quarantine centres were classified into UK dominant B.1.258 (n=7), Danish lineage B.1.428 (n=5) and B.1.351 lineage, first described in South Africa (n=4). The rest of the sequences identified were B.1.1.25 (n=1), B.1.1.365 (n=1), B.1.525 (n=1) and B.1.617.2 (n=1). Both variants of concern (B.1.617.2 and B.1.351) were reported from returnees from India and Middle East.

## Discussion

We report the first description of SARS-CoV-2 molecular epidemiology in Sri Lanka from March 2020 to 30^th^ April 2021. The virus strains identified in March 2020 belonged to clades B.1, B.2, B 1.1 and B.4, demonstrating that SARS-CoV-2 strains were introduced to Sri Lanka from multiple locations [9, 10]. Sri Lanka underwent a national lockdown very early in the pandemic on the 20^th^ of March 2020, when only 66 patients with SARS-CoV-2 were confirmed. This lockdown, which continued until mid-May, managed to contain the outbreak and prevent community transmission, except within isolated community clusters. A further contained outbreak occurred in mid-July within a drug rehabilitation centre (DRC). Sequencing of a limited number of these samples showed that this outbreak was due to viruses belonging to lineage B.1 but which were distinct to the former B.1 samples. The outbreak in the DRC was also subsequently controlled and Sri Lanka did not report any cases of locally acquired infection during the months of August and September. Small numbers of reported cases were from imported infections only.

A large outbreak was abruptly discovered in early October after a clothing factory employee presented with pneumonia caused by SARS-CoV-2, which was followed by the emergence of a large second wave. We report that these were due to a lineage first described in samples from Sri Lanka, B.1.411, that dispersed throughout the country. The molecular clock analysis revealed that this lineage most likely emerged in late June 2020 and therefore, it is possible that the virus was circulating in the community for several months before leading to the large outbreak that started in October. This highlights the potential for cryptic community transmission leading to a national epidemic wave even in the face of strict quarantine rules for returning travellers.

The B.1.411 Sri Lankan lineage has a unique spike mutation H1159Y in the C terminal region, which was seen in 215/231 viruses belonging to this lineage. The significance of this mutation is unknown. Also, the P323L mutation in NSP12 region, which is known to have co-evolved with D614G mutation was seen in 201/231 of the B.1.411 genomes [11]. Even though there is no direct correlation between P323L mutation and infectivity, given the fact that this mutation is widespread and almost 100% co-existent with D614G, some argue that this mutation could contribute to the enhanced viral replication and infectivity seen in D614G dominant strains [11].

Most importantly, number of B.1.411 genomes carrying the E484K (n=1) and N439K (n=4) mutations in the spike protein were detected from both community and quarantine centres between January to mid-February 2021, demonstrating the potential for this lineage to evolve mutations that may evade antibody responses. Even though E484K mutation is predominantly seen in B.1.351 and P.1 lineages, recent evidence indicates introduction of this mutation into other lineages such as B.1.1.7 and B.1.243 [12]. N439K mutation in the receptor binding motif (RBM) of the spike protein is known to enhance the binding affinity of the spike protein to human ACE2 receptor and increase the resistance against several neutralizing monoclonal antibodies [13]. In addition, three of those genomes showed the S:H69/V70 mutation, which often co-occurs in the RBM with amino acid replacements such as N439K. This also is shown to associate with increased infectivity [14]. None of these were identified within the community.

The other more frequent mutations were T166I in NSP2, L37F in NSP6, and T205I in N-protein. The L37F mutation in NSP6 is thought to render the NSP6 protein less stable and therefore, compromise the function of NSP6 [15]. The other mutations have been frequently reported in many other SARS-CoV-2 lineages [16] while the other changes that were detected in the amino acids have not been associated with increased or reduced virulence.

Since the emergence of the ‘second wave’ of SARS-CoV-2 infections in early October 2020, all repatriation from overseas was stopped for a few months and subsequently, restarted in December 2020. Along with this, viruses of many lineages were identified within the quarantine centers where overseas returnees were housed. Importantly, the B.1.1.7 variant, which has been associated with higher transmissibility [17] was initially identified within these quarantine centres, but later from the community from samples sequenced from April 2021. The introduction and spread of the B.1.1.7 led to an exponential rise in the number of cases, along with the number of deaths[18]. Despite strict quarantine for returning travellers, where several imported B.1.1.7 cases were detected, a period of relative quiescence has been followed by an explosive increase in cases across the country. During a period of one month, it appears that the B.1.1.7 lineage had almost completely replaced the circulating B.1.411 due to is higher transmissibility [17]. The introduction of B.1.1.7 has also resulted in higher case fatality rates (CFRs). For instance, the COVID-19 until to 31^st^ of March was 0.63%, whereas since the 1^st^ of April the CFRs has risen to 0.89%, and is 1.09% during 15^th^ to 21^st^ May [18]. The rise in the CFRs could be due to increase in the number of patients requiring health care or possibly due to the increase in viral virulence [19].

In summary, the viruses identified in March 2020, appear to be predominantly introduced by multiple sources such as from Europe and the Middle East and these strains were responsible for the subsequent outbreaks that were seen in Sri Lanka until July. The large outbreak that started in early October, appears to be due to spread of a single virus lineage, B.1.411 until to end of March 2021. The current exponential rise in case numbers appears to be due to the introduction of B.1.1.7 into the community, rapidly displacing the previous circulating B.1.411. As SARS-CoV-2 vaccine rollout commences in Sri Lanka, ongoing genomic surveillance for variants of concern will be vital.

## Data Availability

All data is available within the manuscript and the supporting information files.

## Acknowledgement

We thank Dr. Julian Villabonas-Arenas for sharing some scripts for viral sequence analysis.

