## Supplementary table 1 for "Genomic and epidemiological analysis of SARS-CoV-2 viruses in Sri Lanka"

**Supplementary Table 1. Metadata of 373 SARS-CoV2 samples sequenced in Sri Lanka between March 2020 to April 2021.**

| # | Sequence ID | GISAID Accession ID | Outbreak Period | Pango Lineage | Clade | Collection Month | AA Substitutions |
|---|---|---|---|---|---|---|---|
| 1 | CoV53 | EPI_ISL_428671 | A | B.1.1 | GR | Mar-20 | N_R203K, N_G204R, NSP12_P323L, Spike_D614G |
| 2 | Cov38 | EPI_ISL_428670 | A | B.4 | O | Mar-20 | NSP14_M315I, NSP2_V198I, NS3_I263M, NSP6_L37F, NSP15_S288F, N_A398S |
| 3 | CoV91 | EPI_ISL_428672 | A | B.1 | G | Mar-20 | NSP3_M494I, NSP12_P323L, Spike_D614G, M_A2V |
| 4 | COV194 | EPI_ISL_525478 | A | B.1.1 | GR | Mar-20 | NS6_I60V, N_R203K, N_G204R, NSP12_P323L, Spike_D614G |
| 5 | CoV486 | EPI_ISL_428673 | A | B | O | Mar-20 | NS3_G251V |
| 6 | COV503 | EPI_ISL_525479 | A | B.4.7 | O | Mar-20 | N_P199S, NSP2_V198I, NSP2_R27C, NS8_G8stop, NS3_M260T, NSP14_D345G, NSP4_M33I, NSP6_L37F |
| 7 | C142 | EPI_ISL_525474 | B | B.4 | O | Apr-20 | NSP14_M315I, NSP2_V198I, NS3_A98S, NS3_I263M, NS7b_H42Y, NSP6_L37F, NSP15_S288F, N_A398S |
| 8 | 1885 | EPI_ISL_525476 | B | B.1 | G | Apr-20 | NS6_I60V, NSP12_D893B, NSP3_P125L, NSP12_P323L, Spike_D614G, NSP5_V157I |
| 9 | 7066 | EPI_ISL_525481 | C | B | O | Jun-20 | NSP2_V198I, NSP2_R27C, NSP12_D893B, E_A41T, N_S186F, Spike_T478R, NSP4_M33I, NSP6_L37F, Spike_D80Y, NSP12_D208B |
| 10 | KK224 | EPI_ISL_525488 | D | B.1 | GH | Jul-20 | Spike_G769V, NS3_V90F, NSP3_P74S, NS3_Y156H, NSP12_K91R, NS3_Q57H, NSP5_A234V, NSP2_T85I, NSP2_F93V, NSP12_A706T, Spike_D614G |
| 11 | KK230 | EPI_ISL_525489 | D | B.1 | GH | Jul-20 | NSP3_S377I, Spike_G769V, NS3_V90F, NSP3_P74S, NS3_Y156H, Spike_P1162L, NS3_Q57H, NSP5_A234V, NSP3_V50A, NSP2_T85I, NSP2_K384E, NSP12_P323L, NSP12_A706T, Spike_D614G |
| 12 | KK57 | EPI_ISL_525486 | D | B.1 | GH | Jul-20 | NSP3_S377I, Spike_G769V, NS3_V90F, NSP3_P74S, NS3_Y156H, NS3_Q57H, NSP5_A234V, NSP2_T85I, NSP12_A706T, Spike_D614G, N_Q418H |
| 13 | SL-MIN01 | EPI_ISL_602564 | E | B.1.411 | GH | Oct-20 | NSP12_D445G, NSP12_M666I, NS8_Q18stop, Spike_H1159Y, N_T205I, NSP2_T166I, NS3_Q57H, NSP2_T85I, NSP12_P323L, Spike_D614G, NSP6_L37F |
| 14 | SL-CMC9932 | EPI_ISL_602572 | E | B.1.411 | O | Oct-20 | NSP12_M666I, Spike_H1159Y, NSP2_T166I, NS3_Q57H, NSP2_T85I, NSP12_P323L, NSP6_L37F |
| 15 | SL-CMC9950 | EPI_ISL_602573 | E | B.1.411 | GH | Oct-20 | NSP12_M666I, Spike_H1159Y, NSP2_T166I, NS3_Q57H, NSP2_T85I, Spike_D614G, NSP6_L37F |
| 16 | SL-BIN33 | EPI_ISL_602575 | E | B.1.411 | O | Oct-20 | NSP2_ins211TSX, NSP2_E210D, NSP12_M666I, NSP2_T166I, NSP3_Y519N, Spike_L841I, NSP5_Q306R, NSP12_P323L, Spike_D614G, NSP2_S211H, NSP6_L37F |
| 17 | SL-BIN44 | EPI_ISL_602576 | E | B.1.411 | O | Oct-20 | NSP12_M666I, NS8_Q18stop, Spike_H1159Y, N_T205I, NSP2_T166I, NS3_Q57H, NSP2_T85I, NSP2_R222C, NSP12_P323L, Spike_D614G, NSP6_L37F |
| 18 | SL-WB04 | EPI_ISL_602566 | E | B.1.411 | GH | Oct-20 | NSP12_M666I, NS8_Q18stop, Spike_H1159Y, N_T205I, NSP2_T166I, NS3_Q57H, NSP10_P107del, NSP2_T85I, NSP10_V108del, NSP12_P323L, Spike_D614G |
| 19 | SL-WB28 | EPI_ISL_602567 | E | B.1.411 | GH | Oct-20 | NSP12_M666I, Spike_H1159Y, NSP2_T166I, NS3_Q57H, NSP2_T85I, NSP12_P323L, Spike_D614G, NSP6_L37F |
| 20 | SL-CMC11595 | EPI_ISL_602565 | E | B.1.411 | GH | Oct-20 | NSP12_M666I, NS8_Q18stop, Spike_F92H, Spike_S94P, Spike_H1159Y, NSP2_T166I, NS3_Q57H, NSP2_T85I, NSP12_P323L, Spike_D614G, NSP6_L37F, Spike_A93Y |

| | | | | | | | |
|---|---|---|---|---|---|---|---|
| 21 | SL-CSTH3262 | EPI_ISL_602568 | E | B.1.411 | GH | Oct-20 | NSP12_M666I, NS8_Q18stop, Spike_H1159Y, NSP2_T166I, NS3_Q57H, NSP2_T85I, NSP12_P323L, Spike_D614G, NSP6_L37F |
| 22 | SL-GQC12 | EPI_ISL_602574 | E | B.1.411 | GH | Oct-20 | NSP12_M666I, NS8_Q18stop, Spike_H1159Y, N_T205I, NSP2_T166I, NS3_Q57H, NSP2_T85I, NSP12_P323L, Spike_D614G, NSP6_L37F |
| 23 | cov3563 | EPI_ISL_668454 | E | B.1.411 | GH | Nov-20 | NSP12_D445G, NSP12_M666I, NS8_Q18stop, Spike_H1159Y, N_T205I, NSP2_T166I, NS3_Q57H, NSP14_P203L, NSP12_P323L, Spike_D614G, NSP6_L37F |
| 24 | cov3576 | EPI_ISL_668455 | E | B.1.411 | G | Nov-20 | NSP12_M666I, NS8_Q18stop, Spike_H1159Y, N_T205I, NSP2_T166I, NSP12_P323L, Spike_D614G |
| 25 | 7589 | EPI_ISL_668447 | E | B.1.411 | O | Nov-20 | NSP12_M666I, NS8_Q18stop, Spike_H1159Y, N_T205I, NSP2_T166I, NS3_Q57H, NSP12_P323L, Spike_D614G, NSP6_L37F |
| 26 | 14902 | EPI_ISL_668448 | E | B.1.411 | O | Nov-20 | NSP12_M666I, NS8_Q18stop, N_T205I, NSP2_T166I, Spike_D614G, NSP6_L37F |
| 27 | CM10 | EPI_ISL_668453 | E | B.1.411 | G | Nov-20 | NSP12_M666I, Spike_H1159Y, N_T205I, NSP2_T166I, NSP12_P323L, Spike_D614G, NSP6_L37F |
| 28 | 15372 | EPI_ISL_668449 | E | B.1.411 | GH | Nov-20 | NSP12_M666I, Spike_H1159Y, N_T205I, NSP2_T166I, NS3_Q57H, NSP2_T85I, NS7a_A79V, Spike_D614G, NSP6_L37F |
| 29 | 15386 | EPI_ISL_668450 | E | B.1.411 | GH | Nov-20 | NSP12_D445G, NSP12_M666I, NS8_Q18stop, Spike_H1159Y, N_T205I, NSP2_T166I, NS3_Q57H, NS8_E106stop, Spike_D614G, NSP6_L37F |
| 30 | 15899 | EPI_ISL_668451 | E | B.1.411 | G | Nov-20 | NSP12_D445G, NSP12_M666I, Spike_H1159Y, N_T205I, NSP2_T166I, NSP2_T85I, NSP12_P323L, Spike_D614G, NSP6_L37F |
| 31 | 17272 | EPI_ISL_668452 | E | B.1.411 | O | Nov-20 | NSP12_M666I, NS8_Q18stop, Spike_H1159Y, N_T205I, NSP2_T166I, NS3_Q57H, Spike_D614G, NSP6_L37F, Spike_Q14H |
| 32 | CMC34108 | EPI_ISL_792547 | E | B.1.411 | GH | Nov-20 | NSP12_M666I, NS8_Q18stop, Spike_H1159Y, N_T205I, NSP2_T166I, NS3_Q57H, Spike_D614G, N_A134V, NSP6_L37F |
| 33 | cov5260 | EPI_ISL_792552 | E | B.1.411 | O | Nov-20 | NSP12_M666I, NS8_Q18stop, Spike_H1159Y, N_T205I, NSP2_T166I, NS3_Q57H, Spike_D614G |
| 34 | CMC35812 | EPI_ISL_792548 | E | B.1.411 | GH | Nov-20 | NSP12_M666I, NS8_Q18stop, Spike_H1159Y, N_T205I, NS3_Q57H, NSP12_P323L, Spike_D614G |
| 35 | DKP18 | EPI_ISL_792554 | E | B.1.411 | GH | Nov-20 | NSP2_L71F, NSP12_M666I, NS8_Q18stop, Spike_H1159Y, N_T205I, NSP2_T166I, NS3_Q57H, NSP2_T85I, NSP12_P323L, Spike_D614G, NSP6_L37F |
| 36 | DKP5 | EPI_ISL_792553 | E | B.1.411 | GH | Nov-20 | NSP2_L71F, NSP12_M666I, Spike_H1159Y, N_T205I, NS3_Q57H, Spike_D614G |
| 37 | CMC37019 | EPI_ISL_792549 | E | B.1.411 | GH | Nov-20 | NSP12_M666I, NS8_Q18stop, Spike_H1159Y, NS6_N39T, N_T205I, NSP2_T166I, NS3_Q57H, Spike_F186L, Spike_D614G |
| 38 | w65 | EPI_ISL_792556 | E | B.1.411 | O | Dec-20 | NSP12_M666I, Spike_H1159Y, N_T205I, NSP2_T166I, NS3_Q57H, Spike_D614G |
| 39 | CMC60514 | EPI_ISL_1717040 | E | B.1.1 | GH | Dec-20 | NSP12_D445G, NSP13_S80G, NSP12_M666I, Spike_H1159Y, NSP2_T166I, NS3_Q57H, NSP13_D260Y, NSP12_P323L, Spike_D614G, NSP6_L37F, Spike_D178G |
| 40 | CMC60597 | EPI_ISL_1717042 | E | B.1.1 | GH | Dec-20 | NSP12_D445G, NSP13_S80G, NSP12_M666I, NS8_Q18stop, Spike_H1159Y, N_T205I, NS3_L106R, NS3_Q57H, NSP13_D260Y, NSP12_P323L, Spike_D614G, NSP6_L37F, Spike_D178G |
| 41 | CMC60508 | EPI_ISL_1717039 | E | B.1.411 | GH | Dec-20 | NSP12_D445G, NSP12_M666I, NS8_Q18stop, NSP14_D496Y, Spike_H1159Y, Spike_E154K, N_T205I, NSP2_T166I, NS3_Q57H, NSP2_T85I, Spike_D253N, NSP12_P323L, Spike_D614G, NSP6_L37F |

| | | | | | | | |
|---|---|---|---|---|---|---|---|
| 42 | CMC60513 | EPI_ISL_792550 | E | B.1.411 | O | Dec-20 | NSP13_S80G, NSP12_M666I, NS8_Q18stop, Spike_H1159Y, N_T205I, NSP2_T166I, NS3_L106R, NS3_Q57H, NSP13_D260Y, Spike_D614G, Spike_D178G |
| 43 | CMC60515 | EPI_ISL_1717041 | E | B.1.411 | G | Dec-20 | NSP12_D445G, NSP13_S80G, NSP12_M666I, NS8_Q18stop, Spike_H1159Y, N_T205I, NSP2_T166I, NS3_L106R, NSP2_T85I, NSP13_D260Y, NSP12_P323L, Spike_D614G, NSP6_L37F, Spike_D178G |
| 44 | NP47 | EPI_ISL_1717092 | E | B.1.411 | O | Dec-20 | NSP12_D445G, NSP12_M666I, NS8_Q18stop, Spike_H1159Y, N_T205I, NSP2_T166I, NSP2_T85I, NSP12_P323L, Spike_D614G, N_A134V, NSP6_L37F, NSP3_L1096I |
| 45 | NP55 | EPI_ISL_1717093 | E | B.1.411 | GH | Dec-20 | NSP12_D445G, NSP12_M666I, NS8_Q18stop, Spike_H1159Y, N_T205I, NSP2_T166I, NS3_Q57H, NSP12_P323L, Spike_D614G, N_A134V, NSP6_L37F, NSP3_L1096I |
| 46 | ECB1 | EPI_ISL_792555 | E | B.1.1.7 | GR | Jan-21 | NS8_Q27stop, NSP3_T183I, Spike_T716I, NSP6_S106del, N_R203K, Spike_A570D, Spike_N501Y, NSP3_I1412T, NS8_R52I, Spike_P681H, NSP3_L1700F, NSP3_P402L, NSP3_T1501I, NSP6_G107del, NSP3_A890D, Spike_D1118H, NSP6_F108del, NS8_Y73C, N_G204R, NSP12_P323L, Spike_D614G, N_D3L, Spike_S982A, N_S235F |
| 47 | CMC64284 | EPI_ISL_792551 | E | B.1.411 | GH | Jan-21 | NSP12_M666I, NS8_Q18stop, Spike_H1159Y, N_T205I, NSP2_T166I, NS3_Q57H, M_K14I, M_L16Q, M_K15M, NSP12_P323L, Spike_D614G |
| 48 | CMC66839 | EPI_ISL_1717043 | E | B.1.411 | G | Jan-21 | NSP12_M666I, Spike_H1159Y, N_T205I, NSP2_T166I, NSP2_T85I, NSP12_P323L, NSP3_Q1884H, Spike_D614G, NSP6_L37F |
| 49 | CMC68909 | EPI_ISL_1717044 | E | B.1.411 | GH | Jan-21 | NSP12_M666I, NS8_Q18stop, Spike_H1159Y, N_T205I, NSP2_T166I, NS3_Q57H, NSP2_T85I, NSP12_P323L, NSP3_Q1884H, Spike_D614G, NSP6_L37F |
| 50 | CoV7137 | EPI_ISL_872478 | E | B.1.411 | GH | Jan-21 | NSP12_M666I, NS8_Q18stop, Spike_H1159Y, N_T205I, NSP2_T166I, NS3_Q57H, N_M210V, NSP12_P323L, Spike_D614G, NSP6_L37F |
| 51 | N22 | EPI_ISL_1717091 | E | B.1.411 | O | Jan-21 | NSP12_D445G, NS7a_G38stop, NSP12_M666I, NS8_Q18stop, Spike_H1159Y, NSP4_A380V, NSP2_T166I, NS3_Q57H, NSP12_P323L, NS8_Q72H, Spike_D614G, NSP6_L37F |
| 52 | CMC69934 | EPI_ISL_1717046 | E | B.1.1 | GH | Jan-21 | NSP12_D445G, NSP12_M666I, NS8_Q18stop, Spike_H1159Y, NS3_W131C, N_T205I, NSP2_T166I, NS3_Q57H, NSP2_T85I, NSP12_P323L, Spike_D614G, NSP6_L37F, Spike_C1247F |
| 53 | CMC69602 | EPI_ISL_1717045 | E | B.1.411 | GH | Jan-21 | NSP12_D445G, NSP12_M666I, NS8_Q18stop, Spike_H1159Y, N_T205I, NSP2_T166I, NS3_Q57H, NSP2_T85I, NSP12_P323L, Spike_D614G, NSP6_L37F |
| 54 | P2 | EPI_ISL_1717099 | E | B.1.411 | GH | Jan-21 | NSP12_D445G, NSP12_M666I, NS8_Q18stop, Spike_H1159Y, NSP3_M1556L, N_T205I, NSP2_T166I, NS3_Q57H, NSP2_T85I, NS7a_I3F, NSP12_P323L, Spike_D614G, NSP6_L37F, Spike_A222V |
| 55 | P4 | EPI_ISL_1717100 | E | B.1.411 | GV | Jan-21 | NSP12_D445G, NSP12_M666I, NS8_Q18stop, Spike_H1159Y, NSP3_M1556L, N_T205I, NS7a_I3F, NSP12_P323L, Spike_D614G, NSP6_L37F, Spike_A222V |
| 56 | P8 | EPI_ISL_862725 | E | B.1.411 | GH | Jan-21 | NSP12_M666I, NS8_Q18stop, Spike_H1159Y, N_T205I, NSP2_T166I, NS3_Q57H, NSP2_T85I, NSP12_P323L, Spike_D614G, NSP6_L37F, Spike_A222V |
| 57 | CMC70666 | EPI_ISL_1717047 | E | B.1.411 | GH | Jan-21 | NSP14_Y361C, NSP6_T172A, NSP12_M666I, Spike_H1159Y, N_T205I, NSP2_T166I, NS3_Q57H, NSP6_ins171MTARTVYDDG, NS3_T175I, NSP12_P323L, Spike_D614G, NSP6_L37F |

| | | | | | | | |
|---|---|---|---|---|---|---|---|
| 58 | CMC70852 | EPI_ISL_872394 | E | B.1.411 | GH | Jan-21 | NSP2_M404V, NSP12_M666I, NS8_Q18stop, Spike_H1159Y, N_T205I, NSP2_T166I, NS3_Q57H, NSP2_T85I, NSP7_A80V, Spike_L1063F, NSP12_P323L, Spike_D614G, NSP6_L37F |
| 59 | ABG16 | EPI_ISL_862718 | E | B.1.411 | O | Jan-21 | NSP12_M666I, NS8_Q18stop, Spike_H1159Y, N_T205I, NSP2_T166I, NS3_Q57H, Spike_A684V, Spike_D614G, NSP6_L37F, NSP3_P654S |
| 60 | ABG34 | EPI_ISL_862719 | E | B.1.411 | G | Jan-21 | NSP12_D445G, N_K373R, NSP12_M666I, Spike_T716I, NS8_Q18stop, NSP3_I617V, Spike_H1159Y, N_T205I, NSP2_T166I, Spike_D614G, NSP6_L37F |
| 61 | CA78693 | EPI_ISL_1717026 | E | B.1.411 | GH | Jan-21 | NSP12_M666I, NS8_Q18stop, Spike_H1159Y, N_T205I, NSP2_T166I, NS3_Q57H, NSP2_T85I, NS3_K67R, NSP12_P323L, NSP2_V308L, Spike_D614G, NSP6_L37F |
| 62 | CA78694 | EPI_ISL_1717027 | E | B.1.411 | G | Jan-21 | NSP12_D445G, NSP12_M666I, Spike_H1159Y, N_T205I, NSP2_T166I, NSP12_P323L, Spike_D614G, NSP6_L37F |
| 63 | MVP03 | EPI_ISL_872567 | E | B.1.411 | O | Jan-21 | NSP3_P985L, NSP12_M666I, NS8_Q18stop, Spike_H1159Y, N_T205I, NSP2_T166I, NS3_Q57H, NSP2_T85I, NSP9_T24I, NSP12_P323L, Spike_D614G, NSP6_L37F |
| 64 | CA79689 | EPI_ISL_1717028 | E | B.1.411 | GH | Jan-21 | NSP12_M666I, NS8_Q18stop, Spike_H1159Y, N_T205I, NS3_Q57H, NSP12_P323L, Spike_A684V, Spike_D614G, Spike_T1238S, NSP6_L37F, NSP3_P654S |
| 65 | CMC72385 | EPI_ISL_1718051 | E | B.1.411 | GH | Jan-21 | NSP14_R213H, NSP14_R212H, NSP14_L209del, NSP12_M666I, NSP14_D211del, NSP14_T215K, Spike_H1159Y, N_T205I, NSP2_T166I, NSP14_C210del, NSP14_C208del, NSP12_P323L, Spike_D614G, NSP6_L37F, NSP14_F217del |
| 66 | VQC30 | EPI_ISL_862727 | E | B.1.258 | G | Jan-21 | NSP13_H290Y, NSP9_M101I, NSP12_V720I, NSP6_E195D, NSP12_P323L, Spike_D614G |
| 67 | VQC32 | EPI_ISL_862716 | E | B.1.258 | O | Jan-21 | Spike_H69del, NSP2_T175N, NSP13_H290Y, NSP13_A598S, NSP9_M101I, NSP12_V720I, Spike_V70del, NSP14_S134F, NSP6_E195D, Spike_D614G |
| 68 | CMC72613 | EPI_ISL_1717048 | E | B.1.411 | G | Jan-21 | NSP12_M666I, Spike_H1159Y, N_T205I, Spike_D614G, NSP6_L37F |
| 69 | CV80518 | EPI_ISL_1717070 | E | B.1.411 | G | Jan-21 | Spike_H69del, NSP13_H290Y, NSP13_A505P, NSP3_I1683T, Spike_N439K, NSP13_A598S, NSP9_M101I, NSP12_V720I, Spike_V70del, NSP14_S134F, NSP6_E195D, NSP12_P323L, Spike_D614G, NS7b_A15S |
| 70 | CV80604 | EPI_ISL_1717071 | E | B.1.411 | G | Jan-21 | Spike_H69del, NSP13_H290Y, NSP3_I1683T, NSP9_M101I, NSP12_V720I, Spike_V70del, NSP6_E195D, NSP12_P323L, Spike_D614G |
| 71 | CV80630 | EPI_ISL_1717072 | E | B.1.411 | G | Jan-21 | Spike_H69del, NSP13_H290Y, NSP13_A505P, Spike_N439K, NSP13_A598S, NSP9_M101I, NSP12_V720I, Spike_V70del, NSP14_S134F, NSP6_E195D, NSP12_P323L, Spike_D614G |
| 72 | VQC14 | EPI_ISL_862726 | E | B.1.411 | G | Jan-21 | NSP13_H290Y, NSP13_A505P, NSP13_A598S, NSP9_M101I, NSP12_V720I, NSP14_S134F, NSP6_E195D, Spike_D614G |
| 73 | CA81471 | EPI_ISL_1717031 | E | B.1.1 | GH | Jan-21 | NSP12_D445G, NSP12_M666I, NS8_Q18stop, Spike_H1159Y, N_T205I, NSP2_T166I, NS3_Q57H, NS3_V13L, NSP2_T85I, NSP3_V929I, NSP12_P323L, Spike_D614G, NSP6_L37F |
| 74 | CA81615 | EPI_ISL_1717032 | E | B.1.1 | G | Jan-21 | NSP12_D445G, NSP12_M666I, NS8_Q18stop, NSP14_P203F, Spike_H1159Y, NSP2_T166I, NSP12_P323L, Spike_D614G, NSP2_S138L, NSP6_L37F |
| 75 | A7528 | EPI_ISL_862717 | E | B.1.411 | O | Jan-21 | NSP12_M666I, NS8_Q18stop, Spike_H1159Y, N_T205I, NSP2_T166I, NS3_Q57H, NSP12_P323L, NSP3_A488V, Spike_D614G, NSP6_L37F, Spike_G1124V |

| | | | | | | | |
|---|---|---|---|---|---|---|---|
| 76 | CA81375 | EPI_ISL_1718049 | E | B.1.411 | O | Jan-21 | NSP12_D445G, NSP12_M666I, NS8_Q18stop, Spike_H1159Y, NSP2_T166I, NS3_V13L, NSP2_T85I, NSP3_V929I, NSP12_P323L, Spike_D614G, NSP6_L37F |
| 77 | CA81378 | EPI_ISL_1718050 | E | B.1.411 | GH | Jan-21 | NSP12_D445G, NSP12_M666I, NS8_Q18stop, Spike_H1159Y, NSP2_T166I, NS3_Q57H, NSP12_P323L, NSP3_A488V, Spike_D614G, NSP6_L37F, Spike_G1124V |
| 78 | CA81381 | EPI_ISL_1717029 | E | B.1.411 | GH | Jan-21 | NSP12_M666I, NS8_Q18stop, Spike_H1159Y, N_T205I, NSP2_T166I, NS3_Q57H, NSP12_P323L, NSP3_A488V, Spike_D614G, NSP6_L37F, Spike_G1124V |
| 79 | CA81465 | EPI_ISL_1717030 | E | B.1.411 | O | Jan-21 | NSP12_D445G, NSP12_M666I, NS8_Q18stop, Spike_H1159Y, N_T205I, NSP2_T166I, NS3_Q57H, NSP12_P323L, Spike_D614G, NSP6_L37F, Spike_G1124V |
| 80 | CMC73322 | EPI_ISL_1717049 | E | B.1.411 | GH | Jan-21 | NSP12_D445G, NSP12_M666I, NS8_Q18stop, Spike_H1159Y, N_T205I, NSP2_T166I, NS3_Q57H, NSP7_V58A, NSP12_P323L, Spike_D614G, NSP6_L37F |
| 81 | CMC73331 | EPI_ISL_1717050 | E | B.1.411 | GH | Jan-21 | NSP12_M666I, NS8_Q18stop, Spike_H1159Y, N_T205I, NSP2_T166I, NS3_Q57H, NSP2_T85I, NSP12_P323L, NSP3_Q1884H, Spike_D614G, NSP6_L37F, NSP6_V149F, Spike_D80Y |
| 82 | CMC73350 | EPI_ISL_1717051 | E | B.1.411 | G | Jan-21 | NSP12_D445G, NSP12_M666I, NSP15_S308Y, Spike_H1159Y, N_T205I, NSP2_T166I, NSP7_V58A, NSP12_P323L, Spike_D614G, NSP6_L37F |
| 83 | CoV7350 | EPI_ISL_1717067 | E | B.1.411 | GH | Jan-21 | NSP12_D445G, N_K373R, NSP12_M666I, Spike_T716I, NS8_Q18stop, Spike_H1159Y, N_T205I, NS3_Q57H, NSP12_P323L, Spike_D614G, NSP6_L37F |
| 84 | CoV7356 | EPI_ISL_1718055 | E | B.1.411 | G | Jan-21 | NSP12_D445G, NSP12_M666I, NS8_Q18stop, Spike_H1159Y, N_T205I, NSP2_T166I, NS3_L108F, NS8_R52I, NSP12_P323L, Spike_D614G, NSP6_L37F, Spike_S640F |
| 85 | ING2052 | EPI_ISL_1717084 | E | B.1.411 | O | Jan-21 | NSP12_D445G, NSP12_M666I, NS8_Q18stop, Spike_H1159Y, N_T205I, NSP2_T166I, NS3_Q57H, NSP2_T85I, NSP12_P323L, Spike_D614G, NSP6_L37F |
| 86 | ING2084 | EPI_ISL_862720 | E | B.1.411 | GH | Jan-21 | N_D128Y, NSP12_M666I, Spike_H1159Y, N_T205I, NSP2_T166I, NSP3_L1328F, NS3_Q57H, Spike_D614G, NSP6_L37F, NSP2_E201A |
| 87 | M151 | EPI_ISL_862721 | E | B.1.411 | O | Jan-21 | Spike_P1263L, N_K373R, NSP12_M666I, Spike_T716I, NS8_Q18stop, Spike_H1159Y, N_T205I, NSP2_T166I, NS3_Q57H, NS8_V62L, Spike_D614G, NSP6_L37F |
| 88 | MAT36 | EPI_ISL_1717086 | E | B.1.411 | GH | Jan-21 | NSP12_D445G, NSP3_A1311V, NSP12_M666I, NS8_Q18stop, NSP4_A231V, Spike_H1159Y, NSP2_T166I, NS3_Q57H, NSP12_P323L, Spike_D614G |
| 89 | MAT38 | EPI_ISL_862724 | E | B.1.411 | GH | Jan-21 | NSP12_M666I, NS8_Q18stop, Spike_H1159Y, N_T205I, NSP2_T166I, NS3_Q57H, NSP12_P323L, Spike_D614G, NSP6_L37F |
| 90 | MAT39 | EPI_ISL_1718059 | E | B.1.411 | O | Jan-21 | NSP3_A1311V, NSP12_M666I, Spike_H1159Y, N_T205I, NSP12_P323L, Spike_D614G, NSP6_L37F |
| 91 | MAT43 | EPI_ISL_1717087 | E | B.1.411 | GH | Jan-21 | NSP12_D445G, NSP3_A1311V, NSP12_M666I, NS8_Q18stop, Spike_H1159Y, N_T205I, NSP2_T166I, NS3_Q57H, NSP12_P323L, Spike_D614G, NSP6_L37F |
| 92 | MVP46 | EPI_ISL_872568 | E | B.1.411 | GH | Jan-21 | NSP12_M666I, NS8_Q18stop, Spike_H1159Y, N_T205I, NSP2_T166I, NS3_Q57H, NSP2_T85I, NSP3_S1443F, NSP12_P323L, Spike_D614G, NSP6_L37F |

| | | | | | | | |
|---|---|---|---|---|---|---|---|
| 93 | QA31 | EPI_ISL_1717101 | E | B.1.1.7 | O | Jan-21 | Spike_H69del, NS8_Q27stop, NSP3_T183I, NS8_K68stop, NSP6_S106del, N_R203K, Spike_A570D, Spike_N501Y, NSP3_I1412T, NS8_R52I, Spike_P681H, Spike_Y144del, NSP6_G107del, NSP3_A890D, Spike_D1118H, NSP6_F108del, NS8_Y73C, N_G204R, Spike_V70del, NSP12_P323L, Spike_D614G, N_D3L, Spike_S982A |
| 94 | QA71 | EPI_ISL_1717102 | E | B.1.1.7 | GR | Jan-21 | NS8_Q27stop, NSP3_T183I, NSP3_A890D, NSP6_G107del, Spike_T716I, NS8_K68stop, NSP6_S106del, N_R203K, Spike_D1118H, NSP6_F108del, NS8_Y73C, N_G204R, NSP3_I1412T, NS8_R52I, NSP12_P323L, Spike_P681H, Spike_D614G, Spike_Y144del, N_D3L, Spike_S982A, N_S235F |
| 95 | QA72 | EPI_ISL_1717103 | E | B.1.1.7 | GRY | Jan-21 | Spike_H69del, NS8_Q27stop, NSP3_T183I, Spike_T716I, NS8_K68stop, NSP6_S106del, N_R203K, Spike_A570D, Spike_N501Y, NSP3_I1412T, NS8_R52I, Spike_P681H, Spike_Y144del, NSP6_G107del, NSP3_A890D, Spike_D1118H, NSP6_F108del, NS8_Y73C, N_G204R, Spike_V70del, NSP12_P323L, Spike_D614G, N_D3L, Spike_S982A |
| 96 | CMC74873 | EPI_ISL_1718052 | E | B.1.411 | O | Jan-21 | NSP6_L33M, NSP6_ins35VF, NSP12_M666I, NS8_Q18stop, Spike_H1159Y, N_T205I, NSP2_T166I, NSP6_F34V, NSP6_W31Y, NS3_Q57H, NSP2_T85I, NSP6_Q30E, NSP12_P323L, Spike_D614G, NSP6_L37F, NSP6_T29P |
| 97 | NR5 | EPI_ISL_1717098 | E | B.1.1.7 | GRY | Jan-21 | Spike_H69del, NS3_L15F, NS8_Q27stop, NSP3_T183I, NSP6_S106del, N_R203K, Spike_A570D, NSP13_K460R, NSP4_F17L, Spike_N501Y, NSP3_I1412T, NS8_R52I, Spike_Y144del, NSP3_A890D, NSP6_G107del, Spike_D1118H, NSP6_F108del, NS8_Y73C, N_G204R, Spike_V70del, NSP12_P323L, Spike_D614G, N_D3L, Spike_S982A, N_S235F |
| 98 | CMC75126 | EPI_ISL_1717052 | E | B.1.411 | O | Jan-21 | NSP12_D445G, NSP12_M666I, Spike_H1159Y, N_T205I, NSP2_T166I, NS3_Q57H, NSP12_Q822H, NS3_T175I, NS3_E102K, NSP12_P323L, Spike_D614G, NSP6_L37F, Spike_T76I |
| 99 | CMC75184 | EPI_ISL_1717053 | E | B.1.411 | GH | Jan-21 | NSP12_D445G, NSP12_M666I, NS8_Q18stop, Spike_H1159Y, N_T205I, NS3_Q57H, NSP12_P323L, Spike_D614G, NSP6_L37F |
| 100 | CA85022 | EPI_ISL_1717033 | E | B.1.1 | GH | Jan-21 | NSP12_D445G, NSP12_M666I, Spike_H1159Y, N_T205I, NSP2_T166I, NS3_Q57H, NSP12_P323L, Spike_A684V, Spike_D614G, NSP6_L37F, NSP3_P654S |
| 101 | CMC75810 | EPI_ISL_1717054 | E | B.1.258 | G | Jan-21 | NSP12_D445G, NSP12_M666I, Spike_T240I, Spike_H1159Y, N_T205I, NSP2_T166I, NSP2_T85I, NSP14_S503L, NSP12_P323L, Spike_D614G, NSP6_L37F |
| 102 | NR1 | EPI_ISL_1717094 | E | B.1.411 | G | Jan-21 | N_P199S, NSP3_P1261S, NS3_G174V, NSP14_P203L, NSP3_S1206L, NS3_Y264H, N_V72I, NSP12_P323L, Spike_D614G, NSP2_V447F, Spike_T76I |
| 103 | CMC76417 | EPI_ISL_1717056 | E | B.1 | GH | Jan-21 | NSP12_M666I, NS8_Q18stop, Spike_H1159Y, N_T205I, NSP2_T166I, NSP2_L410F, NS3_Q57H, Spike_L5F, Spike_D614G, NSP6_L37F |
| 104 | CMC76300 | EPI_ISL_1717055 | E | B.1.258 | GH | Jan-21 | NSP12_M666I, NS8_Q18stop, Spike_H1159Y, N_T205I, NSP2_T166I, NSP2_L410F, NS3_Q57H, Spike_L5F, NSP2_T85I, Spike_D614G, NSP6_L37F |
| 105 | CMC76418 | EPI_ISL_1717057 | E | B.1.411 | GH | Jan-21 | NSP12_D445G, NSP12_M666I, NS8_Q18stop, Spike_H1159Y, N_T205I, NSP2_L410F, NS3_Q57H, Spike_L5F, NSP2_T85I, NSP12_P323L, Spike_D614G, NSP6_L37F |

| | | | | | | | |
|---|---|---|---|---|---|---|---|
| 106 | CMC76424 | EPI_ISL_1717058 | E | B.1.411 | GH | Jan-21 | NSP12_D445G, NSP12_M666I, NS8_Q18stop, Spike_H1159Y, N_T205I, NSP2_T166I, NS3_Q57H, Spike_L5F, NSP2_T85I, NSP12_P323L, Spike_D614G, NSP6_L37F |
| 107 | CV85548 | EPI_ISL_1717073 | E | B.1.411 | GH | Jan-21 | NSP12_M666I, NS8_Q18stop, Spike_H1159Y, N_T205I, NS3_Q57H, NS3_V13L, NSP2_T85I, NSP4_C296F, NSP12_P323L, Spike_D614G, NSP14_S255I, NSP6_L37F, Spike_V1176F |
| 108 | NR2 | EPI_ISL_1717095 | E | B.1.1.7 | GRY | Jan-21 | Spike_H69del, NS8_Q27stop, NSP3_T183I, Spike_T716I, NSP6_S106del, N_R203K, Spike_N501Y, NSP3_I1412T, NS8_R52I, Spike_P681H, NS7a_Q62stop, NSP6_G107del, NSP3_A890D, Spike_D1118H, NSP6_F108del, NS8_Y73C, N_G204R, Spike_V70del, NS3_T151I, Spike_N149del, NSP12_P323L, Spike_D614G, N_D3L, Spike_S982A, N_S235F |
| 109 | NR3 | EPI_ISL_1717096 | E | B.1.1.7 | O | Jan-21 | Spike_H69del, NS8_Q27stop, NSP3_T183I, NSP6_S106del, N_R203K, Spike_A570D, Spike_L5F, Spike_N501Y, NSP3_I1412T, NS8_R52I, Spike_P681H, Spike_Y144del, NS7a_Q62stop, NSP3_A890D, NSP6_G107del, Spike_D1118H, NSP6_F108del, NS8_Y73C, N_G204R, Spike_V70del, NS3_T151I, NSP12_P323L, Spike_D614G, N_D3L, Spike_S982A, N_S235F |
| 110 | CMC76839 | EPI_ISL_1717059 | E | B.1.411 | O | Jan-21 | NSP13_S80G, NSP12_M666I, NS8_Q18stop, Spike_H1159Y, N_T205I, NSP2_T166I, NS3_L106R, NS3_Q57H, NSP12_P323L, Spike_D614G, NSP6_L37F, Spike_D178G, NSP4_L353F |
| 111 | CMC76926 | EPI_ISL_1717060 | E | B.1.411 | G | Jan-21 | NSP12_D445G, NS8_F120V, NSP12_M666I, Spike_T33S, NS8_I121L, NS8_Q18stop, NSP9_V76A, Spike_H1159Y, N_T205I, NSP12_P323L, Spike_D614G |
| 112 | CMC77019 | EPI_ISL_1717061 | E | B.1.411 | G | Jan-21 | NSP12_D445G, NSP12_M666I, NS8_Q18stop, N_T205I, NSP2_T166I, NSP12_P323L, NSP5_K90R, Spike_D614G, NSP6_L37F |
| 113 | CMC77196 | EPI_ISL_1717062 | E | B.1.411 | O | Jan-21 | NSP2_L550I, N_M234V, NSP12_M666I, NS8_Q18stop, Spike_H1159Y, N_T205I, NSP2_T166I, NS3_Q57H, NSP12_P323L, Spike_D614G, NSP6_L37F, Spike_A783S |
| 114 | CVQ87412 | EPI_ISL_1717081 | E | B.1.1.7 | GRY | Jan-21 | Spike_H69del, NS8_Q27stop, NSP3_T183I, NSP6_S106del, N_R203K, Spike_A570D, Spike_L5F, NSP3_I1412T, NS8_R52I, Spike_P681H, Spike_Y144del, NS7a_Q62stop, NSP3_A890D, NSP6_G107del, Spike_D1118H, NSP6_F108del, NS8_Y73C, N_G204R, Spike_V70del, NS3_T151I, NSP12_P323L, Spike_D614G, N_D3L, Spike_S982A, N_S235F |
| 115 | CVQ87413 | EPI_ISL_1717082 | E | B.1.1.7 | GRY | Jan-21 | Spike_H69del, NS8_Q27stop, NSP3_T183I, NSP6_S106del, N_R203K, Spike_A570D, Spike_L5F, Spike_N501Y, NSP3_I1412T, NS8_R52I, Spike_Y144del, NS3_G100C, NS7a_Q62stop, NSP3_A890D, NSP6_G107del, Spike_D1118H, NSP6_F108del, NS8_Y73C, N_G204R, Spike_V70del, NS3_T151I, NSP12_P323L, Spike_D614G, N_D3L, Spike_S982A, N_S235F |
| 116 | CVQ87434 | EPI_ISL_1717083 | E | B.1.1.7 | O | Jan-21 | Spike_H69del, NS8_Q27stop, NSP3_T183I, Spike_T716I, NSP6_S106del, N_R203K, Spike_A570D, Spike_L5F, Spike_N501Y, NSP3_I1412T, NS8_R52I, Spike_P681H, Spike_Y144del, NS7a_Q62stop, NSP6_G107del, NSP3_A890D, Spike_D1118H, NSP6_F108del, NS8_Y73C, N_G204R, Spike_V70del, NS3_T151I, NSP12_P323L, Spike_D614G, N_D3L, Spike_S982A, N_S235F |

| | | | | | | | |
|---|---|---|---|---|---|---|---|
| 117 | CMC77416 | EPI_ISL_1717065 | E | B.1.258 | O | Jan-21 | NSP12_D445G, NSP12_M666I, NS8_Q18stop, Spike_H1159Y, N_T205I, NSP2_T166I, Spike_E583D, NSP2_T85I, NSP12_P323L, Spike_D614G, NSP6_L37F |
| 118 | CA86899 | EPI_ISL_1717034 | E | B.1.411 | GH | Jan-21 | NSP12_M666I, NS8_Q18stop, Spike_H1159Y, N_T205I, NSP2_T166I, NS3_Q57H, NSP2_T85I, Spike_D287N, NSP12_P323L, Spike_D614G |
| 119 | CA86905 | EPI_ISL_1717035 | E | B.1.411 | G | Jan-21 | NSP12_D445G, NSP12_M666I, NS8_Q18stop, Spike_H1159Y, N_T205I, NSP2_T85I, NSP12_P323L, Spike_A684V, Spike_D614G, NSP6_L37F, NSP3_P654S |
| 120 | CMC77309 | EPI_ISL_1717064 | E | B.1.411 | O | Jan-21 | NSP12_D445G, NSP12_M666I, NS8_Q18stop, Spike_H1159Y, N_T205I, NSP2_T166I, NSP12_P323L, Spike_D614G |
| 121 | CMC77618 | EPI_ISL_1718054 | E | B.1.411 | GH | Jan-21 | NSP12_M666I, NS8_Q18stop, Spike_H1159Y, N_T205I, NSP2_T166I, NS3_Q57H, NS3_L108F, NS8_R52I, NSP12_P323L, Spike_D614G, NSP6_L37F, Spike_S640F |
| 122 | COV7469 | EPI_ISL_1717068 | E | B.1.411 | GH | Jan-21 | NSP12_M666I, NS8_Q18stop, Spike_H1159Y, N_T205I, NSP2_T166I, NS3_Q57H, NSP2_T85I, NSP16_N235R, NSP12_P323L, Spike_D614G, NSP6_L37F, NSP16_ins234MstopM |
| 123 | COV7478 | EPI_ISL_1717069 | E | B.1.411 | GH | Jan-21 | NSP12_D445G, NSP12_M666I, NS8_Q18stop, Spike_H1159Y, NS3_W131C, N_T205I, NSP2_T166I, NS3_Q57H, NSP12_P323L, Spike_D614G, NSP6_L37F |
| 124 | CV86866 | EPI_ISL_1717074 | E | B.1.411 | GH | Jan-21 | NSP12_D445G, Spike_S98P, NSP12_M666I, Spike_H1159Y, N_T205I, NSP2_T166I, NS3_Q57H, NSP9_T24I, NSP12_P323L, Spike_D614G, NSP6_L37F |
| 125 | CV86867 | EPI_ISL_1717075 | E | B.1.411 | GH | Jan-21 | NSP12_D445G, Spike_S98P, NSP12_M666I, Spike_H1159Y, N_T205I, NSP2_T166I, NS3_Q57H, NSP2_T85I, N_V270L, NSP12_P323L, Spike_D614G, NSP6_L37F |
| 126 | CV86870 | EPI_ISL_1717076 | E | B.1.411 | GH | Jan-21 | NSP12_D445G, Spike_S98P, NSP12_M666I, NS8_Q18stop, Spike_H1159Y, N_T205I, NS3_Q57H, NSP12_P323L, Spike_D614G, NSP6_L37F |
| 127 | CV86880 | EPI_ISL_1717077 | E | B.1.411 | G | Jan-21 | NSP12_D445G, Spike_S98P, NSP12_M666I, NS8_Q18stop, Spike_H1159Y, N_T205I, NSP2_T166I, NSP12_P323L, Spike_D614G, NSP6_L37F |
| 128 | CMC77266 | EPI_ISL_1717063 | E | B.1.1 | GH | Jan-21 | NSP12_M666I, NS8_Q18stop, Spike_H1159Y, N_T205I, NSP2_T166I, NS3_Q57H, NS3_K21N, NS7a_A79V, NSP12_P323L, Spike_D614G |
| 129 | CMC77485 | EPI_ISL_1718053 | E | B.1.411 | G | Jan-21 | NSP12_D445G, NSP12_M666I, Spike_H1159Y, N_T205I, NSP2_T166I, NS3_L108F, NS8_R52I, NSP12_P323L, Spike_D614G, Spike_S640F |
| 130 | MAN3 | EPI_ISL_1718058 | E | B.1.411 | O | Jan-21 | NS3_V255del, NSP12_D445G, NSP14_A425V, NSP12_M666I, Spike_H1159Y, NSP13_P504S, NSP2_T166I, NS3_W193L, NSP3_V1936S, NSP12_P323L, Spike_T63A, Spike_D614G, NSP3_V1935L, NSP6_L37F |
| 131 | MAN4 | EPI_ISL_1717085 | E | B.1.411 | G | Jan-21 | NSP12_D445G, Spike_S98P, NSP12_M666I, NS8_Q18stop, Spike_H1159Y, N_T205I, NSP12_P323L, Spike_D614G, NSP6_L37F |
| 132 | NR4 | EPI_ISL_1717097 | E | B.1.1 | GR | Jan-21 | M_D209Y, NS8_A55V, N_R203K, NSP4_M458I, NS3_G224C, N_G204R, NS3_F114S, NSP12_P323L, Spike_D614G, NSP3_N22D |
| 133 | MULQ106 | EPI_ISL_1717088 | E | B.1.1.7 | GRY | Jan-21 | Spike_H69del, NS8_Q27stop, NSP3_T183I, Spike_T716I, NS8_K68stop, NSP6_S106del, N_R203K, Spike_A570D, Spike_N501Y, NSP3_I1412T, NS8_R52I, Spike_P681H, Spike_Y144del, NSP6_G107del, NSP3_A890D, Spike_D1118H, NSP6_F108del, NS8_Y73C, N_G204R, Spike_V70del, NSP12_P323L, Spike_D614G, N_D3L, Spike_S982A |

| | | | | | | | |
|---|---|---|---|---|---|---|---|
| 134 | MULQ112 | EPI_ISL_1717089 | E | B.1.1.7 | GR | Jan-21 | Spike_H69del, NS8_Q27stop, NSP3_T183I, NSP3_A890D, NSP6_G107del, Spike_T716I, NS8_K68stop, NS8_Q18stop, NSP6_S106del, Spike_D1118H, NSP6_F108del, NS8_Y73C, N_G204R, Spike_V70del, Spike_N501Y, NS8_R52I, NSP12_P323L, Spike_D614G, N_D3L, N_S235F |
| 135 | MULQ66 | EPI_ISL_1717090 | E | B.1.1.7 | GRY | Jan-21 | Spike_H69del, NS8_Q27stop, NSP3_T183I, Spike_T716I, NS8_K68stop, NSP6_S106del, N_R203K, Spike_A570D, Spike_N501Y, NSP3_I1412T, NSP15_T33I, NS8_R52I, Spike_P681H, Spike_Y144del, NSP6_G107del, NSP3_A890D, Spike_D1118H, NSP6_F108del, NS8_Y73C, N_G204R, Spike_V70del, NSP12_P323L, Spike_D614G, N_D3L, Spike_S982A |
| 136 | QCF12 | EPI_ISL_1717104 | E | B.1.1.7 | GR | Jan-21 | NS8_Q27stop, NSP3_T183I, NSP3_D339G, Spike_T716I, NS8_K68stop, NSP6_S106del, N_R203K, Spike_A570D, Spike_N501Y, NSP3_I1412T, NS8_R52I, Spike_P681H, N_R195I, NSP6_G107del, NSP3_A890D, Spike_D1118H, NSP3_R407I, NSP6_F108del, NS8_Y73C, N_G204R, NSP12_P323L, Spike_D614G, N_D3L, Spike_S982A |
| 137 | QCF6 | EPI_ISL_1717105 | E | B.1.1.7 | GR | Jan-21 | NS8_Q27stop, NSP3_T183I, NSP3_D339G, Spike_T716I, NS8_K68stop, NSP6_S106del, N_R203K, Spike_A570D, Spike_N501Y, NSP3_I1412T, NS8_R52I, Spike_P681H, Spike_Y144del, N_R195I, NSP6_G107del, NSP3_A890D, Spike_D1118H, NSP6_F108del, NS8_Y73C, N_G204R, NSP12_P323L, Spike_D614G, Spike_S982A |
| 138 | QCF7 | EPI_ISL_1717106 | E | B.1.1.7 | GRY | Jan-21 | Spike_H69del, NS8_Q27stop, NSP3_T183I, Spike_T716I, NS8_K68stop, NSP6_S106del, N_R203K, Spike_A570D, Spike_N501Y, NSP3_I1412T, Spike_P681H, Spike_Y144del, N_R195I, NSP6_G107del, NSP3_A890D, Spike_D1118H, NSP3_R407I, NSP6_F108del, NS8_Y73C, N_G204R, Spike_V70del, NSP12_P323L, Spike_D614G, N_D3L, Spike_S982A, N_S235F |
| 139 | CMC77697 | EPI_ISL_1717066 | E | B.1.258 | GH | Jan-21 | NSP12_D445G, N_K373R, NSP12_M666I, Spike_T716I, NS8_Q18stop, Spike_H1159Y, N_T205I, NSP2_T166I, NS3_Q57H, NSP12_P323L, Spike_D614G, NSP6_L37F |
| 140 | CV87792 | EPI_ISL_1717078 | E | B.1.411 | G | Jan-21 | NSP3_N1680D, NSP12_D445G, Spike_S98P, NSP12_M666I, NS8_Q18stop, Spike_H1159Y, N_T205I, NSP2_T166I, NSP12_P323L, Spike_D614G, NSP6_L37F |
| 141 | CV87795 | EPI_ISL_1717079 | E | B.1.411 | G | Jan-21 | NSP12_D445G, Spike_S98P, NSP12_M666I, NS8_Q18stop, Spike_H1159Y, N_T205I, NSP2_T166I, NSP9_T24I, NSP12_P323L, Spike_D614G, NSP6_L37F |
| 142 | CV87812 | EPI_ISL_1717080 | E | B.1.411 | O | Jan-21 | NSP12_D445G, Spike_S98P, NSP12_M666I, NS8_Q18stop, Spike_H1159Y, N_T205I, NSP2_T166I, NSP9_T24I, NSP12_P323L |
| 143 | CV87923 | EPI_ISL_1718056 | E | B.1.411 | G | Jan-21 | Spike_H69del, NSP13_H290Y, NSP13_A505P, NSP3_I1683T, Spike_N439K, NS7a_P99S, NSP13_A598S, NSP9_M101I, NSP12_V720I, Spike_V70del, NSP14_S134F, NSP6_E195D, NSP12_M380L, Spike_D614G |
| 144 | CV87946 | EPI_ISL_1718057 | E | B.1.411 | O | Jan-21 | Spike_N439K, NSP9_M101I, NSP12_V720I, NSP6_E195D, Spike_D614G |
| 145 | CA88751 | EPI_ISL_1717036 | E | B.1.411 | GH | Jan-21 | NSP2_S378F, NSP12_D445G, NSP12_M666I, Spike_H1159Y, N_T205I, NSP2_T166I, NS3_Q57H, NSP2_T85I, NSP12_P323L, Spike_A684V, Spike_D614G, NSP6_L37F, NSP3_P654S |
| 146 | CA89635 | EPI_ISL_1717037 | E | B.1.411 | O | Feb-21 | NSP12_M666I, NS8_Q18stop, Spike_H1159Y, N_T205I, NSP2_T166I, NS3_Q57H, NSP6_L37F |

| | | | | | | | |
|---|---|---|---|---|---|---|---|
| 147 | CA89803 | EPI_ISL_1717038 | E | B.1.411 | O | Feb-21 | NSP12_D445G, NSP12_M666I, NS8_Q18stop, Spike_H1159Y, N_T205I, NSP2_T166I, NS3_Q57H, NSP12_P323L, Spike_A684V, Spike_D614G, NSP6_L37F, NSP3_P654S |
| 148 | NR125 | EPI_ISL_1233116 | E | B.1.1.7 | G | Feb-21 | Spike_H69del, NS8_Q27stop, NSP3_T183I, Spike_T716I, NSP6_S106del, NS3_T89I, Spike_N501Y, NSP3_I1412T, NS8_R52I, Spike_P681H, NSP12_P227L, NSP3_P67L, NSP6_G107del, NSP3_A890D, NSP3_D782N, Spike_D1118H, NSP6_F108del, Spike_V70del, NSP12_P323L, NSP14_P451S, Spike_D614G, N_D3L, Spike_S982A, N_S235F |
| 149 | C105141 | EPI_ISL_1233065 | E | B.1.411 | G | Feb-21 | NSP12_D445G, NSP2_T388I, NSP12_M666I, NS8_Q18stop, Spike_H1159Y, N_T205I, NSP2_T166I, NSP2_T85I, NSP15_T48I, NSP12_P323L, Spike_D614G, NSP6_L37F, NS3_S171L |
| 150 | CK105351 | EPI_ISL_1233082 | E | B.1.411 | GH | Feb-21 | Spike_N679K, NSP12_D445G, N_T362I, NSP12_M666I, NS8_Q18stop, Spike_H1159Y, N_T205I, NSP2_T166I, NS3_Q57H, NSP2_T497K, NSP12_E254D, NSP12_P323L, Spike_D614G, NSP6_L37F |
| 151 | CK105352 | EPI_ISL_1233083 | E | B.1.411 | O | Feb-21 | Spike_N679K, NSP12_D445G, N_T362I, NSP12_M666I, NS8_Q18stop, Spike_H1159Y, N_T205I, NSP2_T166I, NS3_Q57H, NSP2_T497K, NSP2_T85I, NSP12_E254D, NSP12_P323L, Spike_D614G, NSP6_L37F |
| 152 | CK89939 | EPI_ISL_1233112 | E | B.1.411 | O | Feb-21 | NSP12_D445G, NSP12_M666I, NS8_Q18stop, Spike_H1159Y, NSP2_T166I, NS3_Q57H, Spike_L585F, NSP12_P323L, Spike_D614G, NSP6_L37F |
| 153 | CA90197 | EPI_ISL_1233115 | E | B.1.411 | GH | Feb-21 | NSP12_M666I, NS8_Q18stop, Spike_H1159Y, N_T205I, NSP2_T166I, NS3_Q57H, N_D402Y, NSP2_T85I, NSP12_P323L, Spike_D614G, NSP6_L37F |
| 154 | CA90219 | EPI_ISL_1233071 | E | B.1.411 | GH | Feb-21 | NSP12_D445G, NS8_F120V, NSP12_M666I, Spike_T33S, NS8_I121L, NS8_Q18stop, Spike_H1159Y, N_T205I, NSP2_T166I, NSP13_L581F, NS3_Q57H, NSP12_P323L, Spike_D614G, NSP6_L37F, NSP3_T1072I, NSP2_H208Y |
| 155 | CA90221 | EPI_ISL_1233072 | E | B.1.411 | GH | Feb-21 | NSP12_Q292H, NSP15_A94T, NSP12_D445G, NS8_F120V, NSP12_M666I, Spike_T33S, NS8_I121L, NS8_Q18stop, Spike_H1159Y, N_T205I, NSP2_T166I, NSP13_L581F, NS3_Q57H, NSP12_P323L, Spike_D614G, NSP6_L37F |
| 156 | CA90222 | EPI_ISL_1233073 | E | B.1.411 | GH | Feb-21 | NSP12_Q292H, NSP15_A94T, NSP12_D445G, NS8_F120V, NSP12_M666I, Spike_T33S, NS8_I121L, NS8_Q18stop, Spike_H1159Y, N_T205I, NSP2_T166I, NSP13_L581F, NS3_Q57H, NSP12_P323L, Spike_D614G, NSP6_L37F, NSP3_T1072I |
| 157 | CA90224 | EPI_ISL_1233074 | E | B.1.411 | GH | Feb-21 | NSP12_Q292H, NSP12_D445G, NS8_F120V, NSP12_M666I, Spike_T33S, NS8_I121L, NS8_Q18stop, Spike_H1159Y, N_T205I, NSP2_T166I, NSP13_L581F, NS3_Q57H, NSP2_T85I, NSP12_P323L, Spike_D614G, NSP6_L37F, NSP2_H208Y |
| 158 | CA90544 | EPI_ISL_1233075 | E | B.1.411 | GH | Feb-21 | NS3_R30H, N_T265I, NSP6_C221F, NSP12_M666I, NS8_Q18stop, Spike_H1159Y, NSP2_T166I, NS3_Q57H, NSP12_P323L, Spike_D614G, NSP6_L37F |
| 159 | CA90800 | EPI_ISL_1233120 | E | B.1.411 | O | Feb-21 | NS3_R30H, N_T265I, NSP12_D445G, NSP6_C221F, NSP12_M666I, Spike_H1159Y, N_T205I, NSP2_T166I, NS3_Q57H, NSP12_P323L, Spike_D614G, NSP6_L37F |
| 160 | CA90834 | EPI_ISL_1233076 | E | B.1.411 | G | Feb-21 | N_T265I, NSP12_D445G, NSP6_C221F, NSP12_M666I, NS8_Q18stop, Spike_H1159Y, N_T205I, NSP2_T166I, NSP2_T85I, NSP12_P323L, Spike_D614G, NSP6_L37F |
| 161 | CA90907 | EPI_ISL_1233059 | E | B.1.411 | O | Feb-21 | NSP12_M666I, NS8_Q18stop, Spike_H1159Y, N_T205I, NSP2_T166I, NS3_Q57H, NSP2_T85I, NSP5_V261F, NSP15_V320L, NSP12_P323L, Spike_D614G |

| | | | | | | | |
|---|---|---|---|---|---|---|---|
| 162 | CA91156 | EPI_ISL_1233057 | E | B.1.411 | GH | Feb-21 | N_T265I, NSP12_D445G, NSP6_C221F, NSP12_M666I, NS8_Q18stop, Spike_H1159Y, NSP2_T166I, NS3_Q57H, NSP2_T85I, NSP12_P323L, Spike_D614G, NSP6_L37F |
| 163 | CA91958 | EPI_ISL_1233078 | E | B.1 | G | Feb-21 | NS3_R30H, N_T265I, NSP12_D445G, NSP6_C221F, NSP12_M666I, NS8_Q18stop, Spike_H1159Y, N_T205I, NSP2_T85I, NSP12_P323L, Spike_D614G, NSP6_L37F |
| 164 | CA91568 | EPI_ISL_1233077 | E | B.1.411 | GH | Feb-21 | NS3_R30H, N_T265I, NSP12_D445G, NSP6_C221F, NSP12_M666I, NS8_Q18stop, Spike_H1159Y, N_T205I, NSP2_T166I, NS3_Q57H, NSP2_T85I, NSP12_P323L, Spike_D614G, NSP6_L37F |
| 165 | CA91575 | EPI_ISL_1233054 | E | B.1.411 | GH | Feb-21 | NS3_R30H, NS3_V50I, NSP12_D445G, NSP6_C221F, NSP12_M666I, NS8_Q18stop, Spike_H1159Y, N_T205I, NSP2_T166I, NS3_Q57H, NSP2_T85I, NSP12_P323L, Spike_D614G, NSP6_L37F |
| 166 | CA91924 | EPI_ISL_1233121 | E | B.1.411 | GH | Feb-21 | NS3_R30H, N_T265I, NSP12_D445G, NSP6_C221F, NSP12_M666I, Spike_H1159Y, N_T205I, NSP2_T166I, NS3_Q57H, NSP2_T85I, NSP12_P323L, Spike_D614G, NSP6_L37F |
| 167 | CA91949 | EPI_ISL_1233062 | E | B.1.411 | GH | Feb-21 | NS3_R30H, NSP6_C221F, NSP12_M666I, NS8_Q18stop, Spike_H1159Y, N_T205I, NS3_Q57H, NSP2_T85I, NSP12_P323L, Spike_D614G, NSP6_L37F |
| 168 | VAC01 | EPI_ISL_1233108 | E | B.1.411 | GH | Feb-21 | Spike_N679K, NSP12_D445G, NSP12_M666I, Spike_D936Y, NS8_Q18stop, Spike_H1159Y, N_T205I, NSP2_T166I, NS3_Q57H, NSP2_T85I, NSP12_P323L, Spike_D614G, NSP6_L37F |
| 169 | CA92688 | EPI_ISL_1233122 | E | B.1.411 | GH | Feb-21 | NSP12_D445G, NSP12_M666I, Spike_H1159Y, NSP2_T166I, NS3_Q57H, NSP2_T85I, NSP3_A667T, NSP12_P323L, Spike_D614G, NSP6_L37F |
| 170 | CMC79794 | EPI_ISL_1233084 | E | B.1.411 | GH | Feb-21 | NSP12_D445G, NSP12_M666I, NS8_Q18stop, NSP2_A159S, Spike_H1159Y, NS8_C83S, N_T205I, NSP2_T166I, NS3_Q57H, NSP12_P323L, Spike_D614G, NSP6_L37F |
| 171 | CMC80220 | EPI_ISL_1233085 | E | B.1.411 | O | Feb-21 | NSP14_K349N, NSP3_A690V, NSP12_D445G, NSP12_M666I, NS8_Q18stop, Spike_H1159Y, NSP16_R86K, NSP2_T166I, NS3_Q57H, NSP12_P323L, Spike_D614G, NSP6_L37F |
| 172 | CMC80234 | EPI_ISL_1233064 | E | B.1.411 | GH | Feb-21 | NSP4_I383F, NSP3_A690V, NSP12_D445G, NSP12_M666I, NS8_Q18stop, Spike_H1159Y, NSP16_R86K, N_T205I, NSP2_T166I, NS3_Q57H, NSP3_L1266I, NSP3_S1265del, NSP12_P323L, Spike_D614G, NSP6_L37F |
| 173 | CMC80235 | EPI_ISL_1233118 | E | B.1.411 | G | Feb-21 | NSP4_I383F, NSP3_A690V, NSP12_D445G, NSP14_G481S, NSP12_M666I, Spike_H1159Y, NSP16_R86K, N_T205I, NSP2_T166I, NSP3_L1266I, NSP3_S1265del, NSP12_P323L, Spike_D614G, NSP6_L37F |
| 174 | CMC80313 | EPI_ISL_1233086 | E | B.1.411 | G | Feb-21 | NSP14_K349N, NSP12_D445G, NSP12_M666I, NS8_Q18stop, NSP16_R86K, N_T205I, NSP2_T85I, NSP12_P323L, Spike_D614G, NSP6_L37F |
| 175 | CMC80418 | EPI_ISL_1233128 | E | B.1.411 | O | Feb-21 | NSP12_D445G, NSP12_M666I, Spike_H1159Y, NSP2_T166I, NSP3_T423I, NSP14_H455Y, NSP3_H290Y, NSP12_P323L, Spike_D614G, NSP6_L37F |
| 176 | CMC80451 | EPI_ISL_1233087 | E | B.1.411 | GH | Feb-21 | NSP12_D445G, Spike_T676A, NSP12_M666I, NS8_Q18stop, NSP10_C41S, Spike_H1159Y, N_T205I, NSP2_T166I, NS3_Q57H, NSP2_T85I, NSP12_P323L, Spike_D614G, NSP6_L37F |
| 177 | CA94250 | EPI_ISL_1233123 | E | B.1.411 | G | Feb-21 | NSP12_D445G, N_K373R, NSP12_M666I, Spike_T716I, Spike_H1159Y, N_T205I, NSP2_T166I, NSP2_T85I, NSP12_P323L, Spike_D614G, NSP6_L37F |

| | | | | | | | |
|---|---|---|---|---|---|---|---|
| 178 | CA94316 | EPI_ISL_1233079 | E | B.1.411 | O | Feb-21 | NS3_T151S, NSP12_D445G, Spike_T676A, NSP12_M666I, NS8_Q18stop, Spike_H1159Y, N_T205I, NSP2_T166I, NS3_Q57H, NSP12_P323L, Spike_D614G, M_S4F, NSP6_L37F |
| 179 | CA94371 | EPI_ISL_1233124 | E | B.1.411 | O | Feb-21 | NS3_R30H, NSP12_D445G, NSP6_C221F, NSP12_M666I, Spike_H1159Y, N_T205I, NSP2_T166I, NSP12_P323L, Spike_D614G, NSP6_L37F |
| 180 | CA94441 | EPI_ISL_1233125 | E | B.1.411 | O | Feb-21 | NSP12_D445G, NSP12_M666I, N_T205I, NSP2_T166I, Spike_V213L, NSP3_S1675I, Spike_V367F, NSP12_P323L, Spike_D614G, NSP6_L37F |
| 181 | CMC82058 | EPI_ISL_1233088 | E | B.1.411 | G | Feb-21 | NSP12_D445G, N_K373R, NSP12_M666I, Spike_T716I, NS8_Q18stop, Spike_H1159Y, N_T205I, NSP2_T166I, NSP2_T85I, NSP12_P323L, Spike_D614G, NSP6_L37F |
| 182 | CMC82201 | EPI_ISL_1239367 | E | B.1.411 | GH | Feb-21 | NSP12_M666I, NS8_Q18stop, Spike_H1159Y, NS8_T26I, N_T205I, NSP2_T166I, NS3_Q57H, NSP2_T85I, NS3_V202L, NSP13_V266L, NSP12_P323L, Spike_D614G, NSP6_L37F |
| 183 | CMC82662 | EPI_ISL_1233092 | E | B.1.411 | GH | Feb-21 | NS3_Y107C, NSP12_D445G, NSP15_S261L, NSP12_M666I, NS8_Q18stop, Spike_H1159Y, N_T205I, NSP2_T166I, NS3_Q57H, NSP2_T85I, NSP12_P323L, Spike_D614G, M_A2S, NSP6_L37F, NSP3_P654S |
| 184 | CMC82701 | EPI_ISL_1233093 | E | B.1.411 | GH | Feb-21 | NS3_R30H, N_T265I, NS3_A23S, NSP12_D445G, NSP6_C221F, NSP12_M666I, NS8_Q18stop, NSP10_V7L, Spike_H1159Y, N_T205I, NSP2_T166I, NS3_Q57H, NSP2_T85I, NSP12_P323L, Spike_D614G, NSP6_L37F |
| 185 | CMC82283 | EPI_ISL_1233089 | E | B.1.411 | G | Feb-21 | NSP12_D445G, NSP12_M666I, NS8_Q18stop, Spike_H1159Y, N_T205I, NSP2_T166I, NSP2_T85I, NSP12_P323L, Spike_D614G, NSP6_L37F |
| 186 | CMC82395 | EPI_ISL_1233090 | E | B.1.411 | G | Feb-21 | NSP12_D445G, NSP12_M666I, NS8_Q18stop, Spike_H1159Y, N_T205I, NSP2_T166I, NSP2_T85I, NS3_D199Y, NSP12_P323L, Spike_D614G, NSP6_L37F |
| 187 | CMC82819 | EPI_ISL_1233094 | E | B.1.411 | GH | Feb-21 | NSP14_K349N, NSP12_D445G, NSP12_M666I, NS8_Q18stop, Spike_H1159Y, NSP16_R86K, N_T205I, NSP2_T166I, NS3_Q57H, NSP2_T85I, NSP12_P323L, Spike_D614G, NSP6_L37F |
| 188 | CMC82904 | EPI_ISL_1233129 | E | B.1.411 | G | Feb-21 | NSP12_D445G, NSP12_M666I, Spike_H1159Y, N_T205I, NSP2_T166I, NSP12_P323L, Spike_D614G, NSP6_L37F |
| 189 | CMC83045 | EPI_ISL_1233095 | E | B.1.411 | G | Feb-21 | NSP12_D445G, NSP12_M666I, NS8_Q18stop, Spike_H1159Y, N_T205I, NSP2_T166I, NS3_V13L, NSP12_P323L, Spike_D614G |
| 190 | CMC83064 | EPI_ISL_1233096 | E | B.1.411 | O | Feb-21 | NSP12_D445G, NS8_F120V, NSP12_M666I, NS8_I121L, NS8_Q18stop, NSP9_V76A, Spike_H1159Y, N_T205I, NSP2_T166I, NSP3_V267F, NS3_Q57H, Spike_D614G, NSP6_L37F, NSP2_H208Y |
| 191 | CMC83303 | EPI_ISL_1239361 | E | B.1.411 | GH | Feb-21 | NSP12_M666I, NS8_Q18stop, Spike_H1159Y, N_T205I, NSP2_T166I, NS3_Q57H, NSP2_T85I, NS3_L108F, NS8_R52I, NSP12_P323L, Spike_D614G, NSP6_L37F, Spike_S640F |
| 192 | CMC83312 | EPI_ISL_1233097 | E | B.1.411 | GH | Feb-21 | NSP12_D445G, NSP12_M666I, NS8_Q18stop, Spike_H1159Y, N_T205I, NSP2_T166I, NS3_Q57H, NSP2_T85I, NSP12_P323L, Spike_D614G, NSP6_L37F, Spike_E748D |
| 193 | RTH6 | EPI_ISL_1233106 | E | B.1.411 | O | Feb-21 | NSP12_D445G, NSP2_T388I, NSP12_M666I, NS8_Q18stop, Spike_H1159Y, N_T205I, NSP2_T85I, NSP15_T48I, NSP12_P323L, NS3_Q185H, Spike_D614G, NSP6_L37F, NS3_S171L |

| | | | | | | | |
|---|---|---|---|---|---|---|---|
| 194 | RTH7 | EPI_ISL_1233136 | E | B.1.411 | G | Feb-21 | NSP13_P326L, Spike_N751K, Spike_H1101Y, NSP12_D445G, NSP12_M666I, N_T205I, NSP3_L862F, NSP12_P323L, Spike_D614G, NSP6_L37F |
| 195 | RTH8 | EPI_ISL_1233137 | E | B.1.411 | O | Feb-21 | NSP3_I967T, NSP12_D445G, NSP2_T388I, NSP12_M666I, Spike_H1159Y, NSP2_T166I, NS3_Q57H, NSP16_D26H, NSP3_A1280V, NSP15_T48I, NSP12_P323L, Spike_D614G, NSP6_L37F, NS3_S171L |
| 196 | CMC83757 | EPI_ISL_1233098 | E | B.1.258 | GH | Feb-21 | NSP12_D445G, NSP12_M666I, Spike_T240I, NS8_Q18stop, Spike_H1159Y, N_T205I, NSP2_T166I, NS3_Q57H, NSP14_S503L, NSP12_P323L, Spike_D614G, NSP6_L37F |
| 197 | CA99139 | EPI_ISL_1233126 | E | B.1.411 | G | Feb-21 | NSP12_D445G, NSP12_M666I, Spike_E484K, Spike_H1159Y, N_T205I, NSP2_T166I, NSP2_T85I, NS3_V202L, NSP12_P323L, Spike_D614G, NSP6_L37F |
| 198 | CA99145 | EPI_ISL_1233080 | E | B.1.411 | GH | Feb-21 | NSP12_D445G, NSP12_M666I, NS8_Q18stop, Spike_H1159Y, N_T205I, NSP2_T166I, NS3_Q57H, NSP2_T85I, NSP12_P323L, Spike_A684V, Spike_D614G, NSP6_L37F, NSP3_P654S |
| 199 | CA99150 | EPI_ISL_1233081 | E | B.1.411 | GH | Feb-21 | NSP12_D445G, NSP12_M666I, NS8_Q18stop, Spike_H1159Y, N_T205I, NSP2_T166I, NS3_Q57H, NSP2_T85I, NSP12_P323L, Spike_A684V, Spike_D614G, NSP6_L37F, NSP3_P654S |
| 200 | CMC82521 | EPI_ISL_1233091 | E | B.1.411 | G | Feb-21 | NSP2_T170I, NSP3_L72F, NSP12_D445G, NSP12_M666I, NS8_Q18stop, Spike_H1159Y, NSP2_T166I, NSP12_D135Y, NSP6_F35L, NSP12_P323L, Spike_D614G, NSP6_L37F |
| 201 | CMC83433 | EPI_ISL_1239364 | E | B.1.411 | GH | Feb-21 | NSP12_D445G, NSP12_M666I, NS8_Q18stop, Spike_H1159Y, N_T205I, NSP2_T166I, NS3_Q57H, NSP2_T85I, NS3_L108F, NS8_R52I, NSP12_P323L, Spike_D614G, NSP3_K1804N, NSP6_L37F, Spike_S640F |
| 202 | CMC83434 | EPI_ISL_1239365 | E | B.1.411 | GH | Feb-21 | NSP12_D445G, NSP12_M666I, NS8_Q18stop, Spike_H1159Y, N_T205I, NSP2_T166I, NS3_Q57H, NS3_L108F, NS8_R52I, NSP12_P323L, Spike_D614G, NSP3_K1804N, NSP6_L37F, Spike_S640F |
| 203 | CMC83435 | EPI_ISL_1239363 | E | B.1.411 | GH | Feb-21 | NSP12_N9Y, NSP12_D445G, NSP12_M666I, Spike_H1159Y, N_T205I, NSP2_T166I, NS3_Q57H, NSP2_T85I, NS3_L108F, NSP12_ins9stop, NS8_R52I, NSP12_R10C, NSP12_P323L, Spike_D614G, NSP3_K1804N, NSP6_L37F, NSP12_L8C, Spike_S640F |
| 204 | CMC83466 | EPI_ISL_1239366 | E | B.1.411 | GH | Feb-21 | NSP12_M666I, NS8_Q18stop, Spike_H1159Y, N_R209K, N_T205I, NSP2_T166I, NS3_Q57H, NSP2_T85I, NS3_L108F, NS8_R52I, NSP12_P323L, Spike_D614G, NSP3_K1804N, NSP6_L37F |
| 205 | CMC83690 | EPI_ISL_1233113 | E | B.1.411 | GH | Feb-21 | NSP12_M666I, NS8_Q18stop, NSP13_I333V, Spike_H1159Y, N_T205I, NSP2_T166I, NS3_Q57H, NSP2_T85I, NSP12_P323L, Spike_D614G, NSP6_L37F, Spike_D178G |
| 206 | CMC83755 | EPI_ISL_1233055 | E | B.1.411 | GH | Feb-21 | NSP12_M666I, NS8_Q18stop, Spike_H1159Y, N_T205I, NSP2_T166I, NS3_Q57H, N_D402Y, Spike_V622F, NSP2_T85I, NSP12_P323L, Spike_D614G, NSP6_L37F |
| 207 | RTH2 | EPI_ISL_1239368 | E | B.1 | G | Feb-21 | Spike_L18F, NSP12_D445G, NSP12_M666I, NS8_Q18stop, Spike_H1159Y, N_T205I, NSP14_P203L, NSP2_T85I, NSP12_P323L, Spike_D614G, NSP6_L37F |
| 208 | CMC83980 | EPI_ISL_1233130 | E | B.1.411 | GH | Feb-21 | NSP12_D445G, NSP8_T148I, NSP12_M666I, Spike_H1159Y, N_T205I, NSP2_T166I, NS3_Q57H, NSP2_T85I, NSP12_P323L, Spike_D614G, NSP6_L37F, NSP3_G17C |

| | | | | | | | |
|---|---|---|---|---|---|---|---|
| 209 | CMC83982 | EPI_ISL_1233099 | E | B.1.411 | GH | Feb-21 | NSP12_D445G, NSP3_K529R, NSP8_T148I, NSP12_M666I, NS8_Q18stop, Spike_H1159Y, NSP2_T166I, NS3_Q57H, NSP2_T85I, NSP12_P323L, Spike_D614G, NS8_I58V, NSP6_L37F, NSP3_G17C |
| 210 | CMC83991 | EPI_ISL_1233100 | E | B.1.411 | GH | Feb-21 | NSP12_D445G, NSP3_K529R, NSP8_T148I, NSP12_M666I, NS8_Q18stop, Spike_H1159Y, N_T205I, NSP2_T166I, NS3_Q57H, NSP2_T85I, NSP12_P323L, Spike_D614G, NS8_I58V, NSP6_L37F |
| 211 | CMC84013 | EPI_ISL_1233056 | E | B.1.411 | GH | Feb-21 | NS7b_L32F, NSP12_M666I, NS8_Q18stop, Spike_H1159Y, N_T205I, NSP2_T166I, NS3_Q57H, NSP2_T85I, NSP12_P323L, Spike_D614G, NSP6_L37F |
| 212 | CMC84237 | EPI_ISL_1233058 | E | B.1.411 | O | Feb-21 | NSP12_M666I, NS8_Q18stop, Spike_H1159Y, N_T205I, NSP2_T166I, NS3_Q57H, NSP2_T85I, NSP12_P323L, Spike_D614G, NSP6_L37F, NS8_P93L |
| 213 | RTH1 | EPI_ISL_1239362 | E | B.1.411 | G | Feb-21 | NSP3_T1303I, NSP12_M666I, NS8_Q18stop, Spike_T859I, N_T205I, NSP2_T166I, NSP12_P323L, Spike_D614G, NSP6_L37F, NSP3_P395L, NSP8_A21V |
| 214 | RTH4 | EPI_ISL_1233105 | E | B.1.411 | G | Feb-21 | Spike_L18F, NSP12_D445G, NSP12_M666I, NS8_Q18stop, Spike_H1159Y, N_T205I, NSP2_T166I, NSP14_P203L, NSP2_T85I, NSP12_P323L, Spike_D614G, NSP6_L37F, NSP3_H1880Y |
| 215 | RTH3 | EPI_ISL_1233135 | E | B.1.411 | O | Feb-21 | NSP12_D445G, NSP12_M666I, Spike_H1159Y, NSP2_T166I, NSP3_A667T, NSP3_H290Y, NSP12_P323L, Spike_D614G |
| 216 | DMC_C_111 | EPI_ISL_1233061 | E | B.1.411 | O | Feb-21 | NSP12_D445G, NSP3_A1321V, NSP12_M666I, NS8_Q18stop, Spike_H1159Y, N_T205I, NSP2_T166I, NS3_Q57H, NSP2_T85I, NSP12_P323L, Spike_D614G, NSP6_L37F |
| 217 | RATT_C_02 | EPI_ISL_1233104 | E | B.1.411 | GH | Feb-21 | NSP3_A1105T, NSP12_D445G, NSP3_P1228S, NSP12_M666I, NS8_Q18stop, Spike_H1159Y, N_T205I, NSP2_T166I, NS3_Q57H, NSP2_T85I, NSP3_Q995H, NSP12_P323L, NS3_S92L, Spike_D614G, NSP6_L37F |
| 218 | RTH5 | EPI_ISL_1233117 | E | B.1.411 | GH | Feb-21 | NSP12_D445G, E_L39del, NSP12_M666I, Spike_H1159Y, N_T205I, NSP2_T166I, NS3_Q57H, E_C40del, NSP3_A667T, N_S201N, NSP3_H290Y, NSP12_P323L, E_A41S, Spike_D614G, NSP6_L37F |
| 219 | CN102751 | EPI_ISL_1233060 | E | B.1.411 | GH | Feb-21 | NS3_T151S, NSP12_D445G, Spike_T676A, NSP12_M666I, NS8_Q18stop, NSP1_V84del, Spike_H1159Y, NSP1_M85V, N_T205I, NSP2_T166I, NS3_Q57H, NSP2_T85I, NSP1_H83del, NSP12_P323L, Spike_D614G, NSP6_L37F, NSP1_G82del, NSP16_P236L |
| 220 | CW103180 | EPI_ISL_1233111 | E | B.1 | GH | Feb-21 | NSP7_T81I, NSP12_D445G, NS3_L41F, NSP12_M666I, NSP16_Q3L, Spike_H1159Y, N_T205I, NSP2_T166I, NS3_Q57H, NS8_E64stop, NSP2_G392R, NSP12_T85I, Spike_D614G, NSP2_K337E, NSP6_L37F |
| 221 | CW103177 | EPI_ISL_1233114 | E | B.1.411 | GH | Feb-21 | NSP7_T81I, NSP12_M666I, NS8_Q18stop, NSP16_Q3L, Spike_H1159Y, N_T205I, NSP2_T166I, NS3_Q57H, NSP2_T85I, NSP2_G392R, NSP12_T85I, NSP14_S450I, Spike_D614G, NSP2_K337E, NSP6_L37F |
| 222 | CW103178 | EPI_ISL_1233131 | E | B.1.411 | G | Feb-21 | NSP7_T81I, NSP12_D445G, NSP12_M666I, NSP16_Q3L, Spike_H1159Y, N_T205I, NSP2_T166I, NSP2_T85I, NSP2_G392R, NSP12_P323L, NSP12_T85I, NSP14_S450I, Spike_D614G, NSP2_K337E, NSP6_L37F |
| 223 | CW103259 | EPI_ISL_1233132 | E | B.1 | GH | Feb-21 | NSP7_T81I, NSP12_D445G, NS3_L41F, NSP16_Q3L, Spike_H1159Y, NSP2_T166I, NS3_Q57H, NSP2_G392R, NSP12_P323L, NSP12_T85I, NSP14_S450I, Spike_D614G, NSP2_K337E, NSP6_L37F |

| | | | | | | | |
|---|---|---|---|---|---|---|---|
| 224 | CA103286 | EPI_ISL_1233066 | E | B.1.411 | GH | Feb-21 | NSP12_D445G, NSP12_M666I, NS8_Q18stop, Spike_H1159Y, N_T205I, NSP2_T166I, NS3_Q57H, NSP14_S503L, NSP12_D284N, NSP12_P323L, Spike_D614G, NSP6_L37F |
| 225 | CA103306 | EPI_ISL_1233067 | E | B.1.411 | G | Feb-21 | NSP12_D445G, NSP12_M666I, NS8_Q18stop, Spike_H1159Y, N_T205I, NSP2_T166I, NSP2_T85I, NSP12_P323L, Spike_A684V, Spike_D614G, NSP6_L37F, NSP3_P654S |
| 226 | CW103261 | EPI_ISL_1233133 | E | B.1.411 | GH | Feb-21 | NSP7_T81I, NSP12_D445G, NS3_L41F, NSP12_M666I, NSP16_Q3L, Spike_H1159Y, N_T205I, NSP2_T166I, NS3_Q57H, NSP2_G392R, NSP12_P323L, NSP12_T85I, NSP14_S450I, Spike_D614G, NSP2_K337E, NSP6_L37F |
| 227 | CW103264 | EPI_ISL_1233101 | E | B.1.411 | G | Feb-21 | NSP7_T81I, NSP12_D445G, NSP12_M666I, NS8_Q18stop, NSP16_Q3L, Spike_H1159Y, N_T205I, NSP2_T166I, NSP2_G392R, NSP12_P323L, NSP12_T85I, NSP14_S450I, Spike_D614G, NSP2_K337E, NSP6_L37F |
| 228 | CW103267 | EPI_ISL_1233102 | E | B.1.411 | GH | Feb-21 | Spike_E180G, NSP12_D445G, NSP12_M666I, NS8_Q18stop, Spike_H1159Y, N_T205I, NSP2_T166I, NS3_Q57H, NSP2_T85I, Spike_D614G, NSP6_L37F |
| 229 | STF103572 | EPI_ISL_1233138 | E | B.1.411 | GH | Feb-21 | Spike_F1121L, NSP12_D445G, NSP12_M666I, Spike_H1159Y, N_T205I, NSP2_T166I, NS3_Q57H, NS8_V62L, NSP12_P323L, Spike_D614G, NSP6_L37F |
| 230 | CA103847 | EPI_ISL_1233068 | E | B.1.411 | G | Feb-21 | NSP12_D445G, NSP12_M666I, NS8_Q18stop, Spike_H1159Y, N_T205I, NSP2_T166I, NS3_D210Y, NSP12_P323L, Spike_D614G, NSP6_L37F |
| 231 | CA103848 | EPI_ISL_1233069 | E | B.1.411 | GH | Feb-21 | NS3_A59T, NSP12_D445G, NSP12_M666I, NS8_Q18stop, Spike_H1159Y, N_T205I, NS3_Q57H, NSP2_T85I, NSP12_P323L, Spike_D614G, NSP6_L37F |
| 232 | CA103909 | EPI_ISL_1233070 | E | B.1.411 | G | Feb-21 | N_K387R, NSP12_D445G, Spike_M1237I, NSP12_M666I, NS8_Q18stop, N_T205I, NSP2_T166I, NSP2_T85I, Spike_A879T, NSP12_P323L, Spike_D614G, NSP6_L37F |
| 233 | CK104003 | EPI_ISL_1233127 | E | B.1.411 | GH | Feb-21 | NSP12_D445G, NSP12_M666I, Spike_H1159Y, N_T205I, NSP2_T166I, NS3_Q57H, NSP2_T85I, NSP12_P323L, Spike_D614G, NSP6_L37F |
| 234 | CQ104596 | EPI_ISL_1233063 | E | B.1.1.7 | GRY | Feb-21 | Spike_H69del, NS8_Q27stop, NSP3_T183I, NSP1_V106A, NSP6_S106del, N_R203K, Spike_A570D, NSP2_L400F, Spike_N501Y, NSP3_I1412T, Spike_Y144del, NSP15_S288F, NSP3_A890D, NSP6_G107del, E_ins38CLL, Spike_D1118H, NSP6_F108del, NS8_Y73C, N_G204R, Spike_V70del, NS3_E261G, NSP12_P323L, Spike_D614G, N_D3L, N_S235F |
| 235 | SEQ01 | EPI_ISL_1233107 | E | B.1.411 | G | Feb-21 | NSP12_D445G, NSP12_M666I, NS8_Q18stop, NSP2_K456R, Spike_H1159Y, N_D144Y, NSP2_T166I, NSP12_P323L, Spike_D614G, NSP6_L37F, NS8_P93L |
| 236 | NR128 | EPI_ISL_1233134 | E | B.1.1.25 | GR | Mar-21 | Spike_V1264L, NS6_I60V, N_R203K, NSP3_K412N, N_G204R, M_F28L, Spike_P681R, NS3_K75N, NS8_V62L, NSP2_I120F, NS8_S54L, NSP12_P323L, Spike_D614G, NSP6_L37F, Spike_Q677H, Spike_W258L |
| 237 | NR123 | EPI_ISL_1233109 | E | B.1.1.7 | GRY | Mar-21 | Spike_H69del, NS8_Q27stop, NSP3_T183I, Spike_T716I, NS8_K68stop, NSP6_S106del, N_R203K, Spike_A570D, NS3_W131C, Spike_E96D, Spike_G1251V, Spike_N501Y, NSP3_I1412T, NS8_R52I, Spike_P681H, Spike_Y144del, NSP6_G107del, Spike_D1118H, NSP6_F108del, N_G204R, Spike_V70del, NSP12_P323L, Spike_D614G, N_D3L, Spike_S982A |

| | | | | | | | |
|---|---|---|---|---|---|---|---|
| 238 | NR126 | EPI_ISL_1233110 | E | B.1.1.7 | GRY | Mar-21 | Spike_H69del, NS8_Q27stop, NSP3_T183I, Spike_T716I, NS8_K68stop, NSP6_S106del, N_R203K, Spike_A570D, NS3_T89I, Spike_N501Y, NSP3_I1412T, NS8_R52I, Spike_P681H, Spike_Y144del, NSP12_P227L, NSP3_P67L, NSP6_G107del, NSP3_A890D, NSP3_D782N, Spike_D1118H, NSP6_F108del, NS8_Y73C, N_G204R, Spike_V70del, NSP12_P323L, NSP14_P451S, Spike_D614G, N_D3L, Spike_S982A |
| 239 | NR127 | EPI_ISL_1233119 | E | B.1.351 | G | Mar-21 | Spike_D215G, E_P71L, NSP3_K837N, Spike_K417N, Spike_L244del, NSP6_G107del, NSP6_S106del, Spike_E484K, N_T205I, NSP6_F108del, Spike_L242del, Spike_A701V, NSP2_T85I, Spike_D80A, Spike_N501Y, NSP12_P323L, NSP4_L264F, NSP5_K90R, Spike_D614G, Spike_A243del, NS3_S171L |
| 240 | NR124 | EPI_ISL_1233103 | E | B.1.411 | GH | Mar-21 | NSP7_T81I, NSP12_D445G, NS3_L41F, NSP12_M666I, NS8_Q18stop, Spike_H1159Y, N_T205I, NSP2_T166I, NS3_Q57H, NSP12_P323L, NSP12_T85I, NSP14_S450I, Spike_D614G, NSP6_L37F, NSP3_N922S |
| 241 | NR22 | EPI_ISL_1533846 | E | B.1.1.7 | GRY | Mar-21 | Spike_H69del, NS8_Q27stop, NSP3_T183I, Spike_T716I, NSP6_S106del, N_R203K, Spike_A570D, Spike_N501Y, NS8_R52I, Spike_P681H, Spike_Y144del, NSP3_I441L, NSP6_G107del, NSP3_A890D, Spike_D1118H, NSP6_F108del, NS8_Y73C, N_G204R, Spike_V70del, NSP12_P323L, Spike_D614G, N_D3L, Spike_S982A, N_S235F |
| 242 | CDRF36 | EPI_ISL_1533804 | E | B.1.1.7 | G | Mar-21 | Spike_H69del, NSP3_T183I, NSP6_G107del, Spike_T716I, NSP6_S106del, N_R203K, Spike_D1118H, NSP6_F108del, NS3_Q57H, N_G204R, Spike_V70del, NSP3_I1412T, NS8_R52I, NSP3_T1830I, Spike_P681H, Spike_D614G, N_D3L, Spike_S982A, N_S235F |
| 243 | CDRI1 | EPI_ISL_1533805 | E | B.1.411 | O | Mar-21 | NSP12_D445G, NSP12_M666I, NS8_Q18stop, Spike_H1159Y, N_T205I, NSP2_T166I, NSP3_L1328F, NSP2_T85I, NSP12_P323L, Spike_D614G, NSP6_L37F, NSP6_V149F, NSP2_E201A |
| 244 | CDRC2 | EPI_ISL_1533803 | E | B.1 | G | Mar-21 | NSP13_S80G, NSP12_M666I, NS8_Q18stop, Spike_H1159Y, N_T205I, NS3_L106R, NSP2_T85I, NSP12_P323L, Spike_D614G, N_D377A, Spike_D178G |
| 245 | CDRS25 | EPI_ISL_1533810 | E | B.1 | G | Mar-21 | NSP12_D445G, NSP12_M666I, N_T205I, NSP2_T166I, NSP2_T85I, NSP12_P323L, Spike_A684V, Spike_D614G, NSP6_L37F, Spike_M1229I, NSP3_P654S, NSP13_A18V |
| 246 | CDRQ4 | EPI_ISL_1533806 | E | B.1.1.7 | O | Mar-21 | NS8_Q27stop, NSP3_T183I, NSP1_V106A, Spike_T716I, NSP6_S106del, Spike_A570D, NSP2_L400F, Spike_N501Y, NSP3_I1412T, NS8_R52I, Spike_P681H, Spike_Y144del, NSP15_S288F, NSP6_G107del, NSP3_A890D, Spike_D1118H, NSP6_F108del, NS8_Y73C, NS3_E261G, NSP12_P323L, Spike_D614G, N_D3L, Spike_S982A |
| 247 | CDRS22 | EPI_ISL_1533807 | E | B.1.411 | O | Mar-21 | NSP12_D445G, NSP12_M666I, Spike_H1159Y, N_T205I, NSP2_T166I, NS3_Q57H, NSP12_P323L, Spike_A684V, Spike_D614G, NSP6_L37F, Spike_M1229I, NSP3_P654S |
| 248 | CDRS23 | EPI_ISL_1533808 | E | B.1.411 | O | Mar-21 | NSP12_D445G, NSP12_M666I, NS8_Q18stop, Spike_H1159Y, N_T205I, NSP2_T166I, NS3_Q57H, NSP2_T85I, NSP12_P323L, Spike_A684V, Spike_D614G, NSP1_E36K, NSP6_L37F, Spike_M1229I, NSP3_P654S |

| | | | | | | | |
|---|---|---|---|---|---|---|---|
| 249 | CDRS24 | EPI_ISL_1533809 | E | B.1.411 | GH | Mar-21 | NSP12_D445G, NSP12_M666I, NS8_Q18stop, Spike_H1159Y, NSP2_T166I, NS3_Q57H, NSP2_T85I, Spike_A684V, Spike_D614G, NSP6_L37F, Spike_M1229I, NSP3_P654S |
| 250 | CDRS26 | EPI_ISL_1533811 | E | B.1.411 | G | Mar-21 | NSP12_M666I, Spike_H1159Y, N_T205I, NSP2_T166I, NSP2_T85I, NSP12_P323L, Spike_A684V, Spike_D614G, NSP6_L37F, NSP3_P654S |
| 251 | CDRS27 | EPI_ISL_1533812 | E | B.1.411 | GH | Mar-21 | NSP12_D445G, NSP12_M666I, NS8_Q18stop, Spike_H1159Y, N_T205I, NSP2_T166I, NS3_Q57H, NSP2_T85I, NSP12_P323L, Spike_A684V, Spike_D614G, NSP6_L37F, Spike_M1229I, NSP3_P654S |
| 252 | CDRS28 | EPI_ISL_1533813 | E | B.1.411 | GH | Mar-21 | NSP12_D445G, NSP12_M666I, NS8_Q18stop, Spike_H1159Y, N_T205I, NSP2_T166I, NS3_Q57H, NSP2_T85I, NSP12_P323L, Spike_A684V, Spike_D614G, NSP6_L37F, Spike_M1229I, NSP3_P654S |
| 253 | CDRS29 | EPI_ISL_1533814 | E | B.1.411 | GH | Mar-21 | NSP12_D445G, NSP12_M666I, NS8_Q18stop, Spike_H1159Y, N_T205I, NSP2_T166I, NS3_Q57H, NSP2_T85I, NSP12_P323L, Spike_A684V, Spike_D614G, NSP6_L37F, Spike_M1229I, NSP3_P654S |
| 254 | CDRS30 | EPI_ISL_1533815 | E | B.1.411 | GH | Mar-21 | NSP12_D445G, NSP12_M666I, Spike_H1159Y, N_T205I, NSP2_T166I, NS3_Q57H, NSP2_T85I, NSP12_P323L, Spike_A684V, Spike_D614G, NSP6_L37F, Spike_M1229I, NSP3_P654S |
| 255 | CDRS31 | EPI_ISL_1533816 | E | B.1.411 | GH | Mar-21 | NSP12_D445G, NSP12_M666I, NS8_Q18stop, Spike_H1159Y, N_T205I, NSP2_T166I, NS3_Q57H, NSP2_T85I, NSP12_P323L, Spike_A684V, Spike_D614G, NSP6_L37F, Spike_M1229I, NSP3_P654S |
| 256 | CDRS33 | EPI_ISL_1533817 | E | B.1.411 | G | Mar-21 | NSP3_D174Y, NSP12_M666I, NS8_Q18stop, Spike_H1159Y, N_T205I, NSP2_T166I, NSP2_T85I, NSP12_P323L, Spike_A684V, Spike_D614G, NSP6_L37F, Spike_M1229I, NSP3_P654S |
| 257 | CMC86624 | EPI_ISL_1533822 | E | B.1.411 | G | Mar-21 | NSP12_D445G, NSP12_M666I, Spike_H1159Y, N_T205I, NSP2_T166I, NS3_P262S, NSP2_T85I, NSP12_P323L, Spike_D614G, NSP6_L37F |
| 258 | NR23 | EPI_ISL_1533847 | E | B.1.1.7 | O | Mar-21 | Spike_H69del, Spike_T716I, NS8_K68stop, NSP6_S106del, N_R203K, Spike_A570D, NS3_T89I, NSP5_L75F, Spike_N501Y, NSP3_I1412T, NS8_R52I, Spike_P681H, Spike_Y144del, NSP12_P227L, NSP3_P67L, NSP6_G107del, NSP3_A890D, NSP3_D782N, Spike_D1118H, NSP6_F108del, NS8_Y73C, N_G204R, Spike_V70del, NSP12_P323L, NSP14_P451S, Spike_D614G, N_D3L |
| 259 | NR24 | EPI_ISL_1533848 | E | B.1.1.7 | O | Mar-21 | NS8_Q27stop, NSP3_T183I, NS8_K68stop, NSP6_S106del, N_R203K, Spike_A570D, NS3_T89I, NSP5_L75F, Spike_N501Y, NS8_R52I, Spike_P681H, Spike_Y144del, NSP12_P227L, NSP3_P67L, NSP6_G107del, NSP3_A890D, NSP3_D782N, Spike_D1118H, NSP6_F108del, NS8_Y73C, N_G204R, NSP14_P451S, Spike_D614G, N_D3L, Spike_S982A |
| 260 | NR9 | EPI_ISL_1533851 | E | B.1.1.7 | G | Mar-21 | Spike_H69del, NS8_Q27stop, NSP3_T183I, Spike_T716I, NS8_K68stop, NSP6_S106del, Spike_A570D, Spike_N501Y, NSP3_I1412T, NS8_R52I, Spike_P681H, Spike_Y144del, NSP12_P227L, NSP3_P67L, NSP6_G107del, NSP3_A890D, NSP3_D782N, Spike_D1118H, NSP6_F108del, NS8_Y73C, Spike_V70del, NSP12_P323L, NSP14_P451S, Spike_D614G, N_D3L, Spike_S982A |
| 261 | CMC86789 | EPI_ISL_1533823 | E | B.1.411 | O | Mar-21 | NSP12_D445G, NSP12_M666I, NS8_Q18stop, Spike_H1159Y, N_T205I, N_D144Y, NSP3_T204I, NSP15_T48I, Spike_D614G, NSP6_L37F, NS8_P93L |

| | | | | | | | |
|---|---|---|---|---|---|---|---|
| 262 | CMC86811 | EPI_ISL_1533824 | E | B.1.411 | G | Mar-21 | NSP3_G777V, NSP3_G337S, NSP12_D445G, NSP12_D618N, NSP12_M666I, Spike_H1159Y, N_T205I, N_D144Y, NSP12_ins617LR, NSP2_T166I, NSP3_ins336DHNY, NSP3_A338H, NSP3_T204I, NSP2_T85I, NSP15_T48I, NSP12_P323L, Spike_D614G, NSP6_L37F, NS8_P93L, NSP12_W617F |
| 263 | CP107829 | EPI_ISL_1533839 | E | B.1.411 | GH | Mar-21 | NSP12_D445G, NSP12_M666I, NS3_E102Q, NSP16_R86K, N_T205I, NSP2_T166I, NS3_Q57H, NSP5_K90R, Spike_D614G, NSP6_L37F, NSP2_G147D |
| 264 | CP107863 | EPI_ISL_1533841 | E | B.1.411 | GH | Mar-21 | NSP3_S284C, NSP12_M666I, NS8_Q18stop, NS3_E102Q, Spike_H1159Y, NSP3_P2L, NSP16_R86K, N_T205I, NSP2_T166I, NS3_Q57H, NSP2_T85I, NSP5_K90R, Spike_D614G, NSP6_L37F, NSP2_G147D |
| 265 | CP107818 | EPI_ISL_1533838 | E | B.1.428 | GH | Mar-21 | NSP12_M666I, NS8_Q18stop, NS3_E102Q, Spike_H1159Y, NSP3_P2L, NSP16_R86K, N_T205I, NSP2_T166I, NS3_Q57H, NSP2_T85I, NSP12_P323L, NSP5_K90R, Spike_D614G, NSP6_L37F, NSP2_G147D |
| 266 | CP107831 | EPI_ISL_1533840 | E | B.1.428 | GH | Mar-21 | NSP3_S284C, NSP12_D445G, NSP12_M666I, NS8_Q18stop, NS3_E102Q, Spike_H1159Y, NSP3_P2L, NSP16_R86K, N_T205I, NSP2_T166I, NS3_Q57H, NSP2_T85I, NSP3_T864I, NSP12_P323L, NSP5_K90R, Spike_D614G, NSP6_L37F, NSP2_G147D |
| 267 | CP107951 | EPI_ISL_1533842 | E | B.1.428 | GH | Mar-21 | NSP3_S284C, NSP12_D445G, NSP12_M666I, NS8_Q18stop, NS3_E102Q, Spike_H1159Y, NSP3_P2L, NSP16_R86K, N_T205I, NSP2_T166I, NS3_Q57H, NSP2_T85I, NSP12_P323L, NSP5_K90R, Spike_D614G, NSP6_L37F, NSP2_G147D |
| 268 | CMC87036 | EPI_ISL_1533825 | E | B.1.411 | G | Mar-21 | NS3_T151S, NSP12_D445G, Spike_T676A, NSP12_M666I, Spike_H1159Y, N_T205I, NSP2_T166I, NSP12_P323L, Spike_D614G, NSP6_L37F, NSP14_P140S |
| 269 | CMC87038 | EPI_ISL_1533826 | E | B.1.411 | G | Mar-21 | NS3_T151S, NSP12_D445G, Spike_T676A, NSP12_M666I, NS8_Q18stop, Spike_H1159Y, N_T205I, NSP2_T166I, NSP12_P323L, Spike_D614G, NSP6_L37F |
| 270 | CMC87040 | EPI_ISL_1533827 | E | B.1.411 | G | Mar-21 | NS3_T151S, NSP12_D445G, Spike_T676A, NSP12_M666I, NS8_Q18stop, Spike_H1159Y, NSP2_T166I, NSP12_P323L, Spike_D614G, NSP6_L37F |
| 271 | CMC87041 | EPI_ISL_1533828 | E | B.1.411 | GH | Mar-21 | NS3_T151S, NSP12_D445G, NSP12_M666I, Spike_H1159Y, NSP2_T166I, NS3_Q57H, NSP2_T85I, NSP12_P323L, Spike_D614G, NSP6_L37F |
| 272 | CMC87046 | EPI_ISL_1533829 | E | B.1.411 | G | Mar-21 | NS3_T151S, NSP12_D445G, NSP12_M666I, NS8_Q18stop, Spike_H1159Y, N_T205I, NSP2_T166I, NSP12_P323L, Spike_D614G, NSP6_L37F |
| 273 | CMC87068 | EPI_ISL_1533830 | E | B.1.411 | O | Mar-21 | NS3_T151S, NSP12_D445G, Spike_T676A, NSP12_M666I, NS8_Q18stop, Spike_H1159Y, N_T205I, NSP2_T166I, NS3_Q57H, NSP12_P323L, Spike_D614G, NSP6_L37F |
| 274 | CMC87070 | EPI_ISL_1533831 | E | B.1.411 | O | Mar-21 | NSP10_T12I, NS3_T151S, NSP12_D445G, NSP12_M666I, Spike_H1159Y, NSP2_T166I, NS3_Q57H, NSP12_P323L, Spike_D614G, NSP6_L37F |
| 275 | CM108341 | EPI_ISL_1533821 | E | B.1.411 | O | Mar-21 | NS8_L95F, NS8_W45C, NSP12_D445G, NSP16_Q238H, NSP12_M666I, NS8_Q18stop, Spike_H1159Y, N_T205I, NSP2_T166I, NSP1_G30S, E_S55F, NSP3_S1670F, NS7a_L102P, NSP12_P323L, NSP5_K90R, Spike_D614G, NSP6_L37F |
| 276 | CMC87128 | EPI_ISL_1533832 | E | B.1.411 | G | Mar-21 | NS3_T151S, NSP12_D445G, NSP12_M666I, NS8_Q18stop, Spike_H1159Y, N_T205I, NSP2_T166I, NSP12_P323L, Spike_D614G, NSP6_L37F |
| 277 | CMC87130 | EPI_ISL_1533833 | E | B.1.411 | G | Mar-21 | NS3_T151S, NSP12_D445G, Spike_T676A, NSP12_M666I, NS8_Q18stop, Spike_H1159Y, N_T205I, NSP2_T166I, NSP12_P323L, Spike_D614G, NSP6_L37F |

| | | | | | | | |
|---|---|---|---|---|---|---|---|
| 278 | CMC87137 | EPI_ISL_1582411 | E | B.1.411 | O | Mar-21 | NS3_T151S, NSP12_D445G, Spike_T676A, NSP12_M666I, NS8_Q18stop, Spike_H1159Y, N_T205I, NSP2_T166I, NSP2_T85I, NSP12_P323L, Spike_D614G |
| 279 | CMC87140 | EPI_ISL_1533834 | E | B.1.411 | G | Mar-21 | NS3_T151S, NSP12_D445G, Spike_T676A, NSP12_M666I, Spike_H1159Y, N_T205I, NSP2_T166I, NSP12_P323L, Spike_D614G |
| 280 | CMC87144 | EPI_ISL_1533835 | E | B.1.411 | O | Mar-21 | NS3_T151S, NSP12_D445G, Spike_T676A, NSP12_M666I, NS8_Q18stop, Spike_H1159Y, N_T205I, NSP2_T166I, NSP1_G137S, NSP12_P323L, Spike_D614G, NSP6_L37F |
| 281 | CMC87190 | EPI_ISL_1533836 | E | B.1.411 | O | Mar-21 | NS3_T151S, NSP12_D445G, Spike_T676A, NSP12_M666I, Spike_H1159Y, N_T205I, NSP2_T166I, NS3_Q57H, Spike_D614G, NSP6_L37F |
| 282 | CMC87205 | EPI_ISL_1533837 | E | B.1.411 | O | Mar-21 | NS3_T151S, NSP12_D445G, Spike_T676A, NSP12_M666I, Spike_H1159Y, N_T205I, NSP2_T166I, NS3_Q57H, NS3_V55I, NSP12_P323L, Spike_D614G, NSP6_L37F |
| 283 | NR18 | EPI_ISL_1582414 | E | B.1 | O | Mar-21 | Spike_D215G, E_P71L, NSP3_K837N, Spike_K417N, Spike_L244del, NSP6_G107del, NS8_I121L, NSP6_S106del, Spike_E484K, N_T205I, NSP6_F108del, Spike_L242del, NS3_Q57H, NSP12_T120E, Spike_D80A, NSP12_P323L, Spike_D614G, NSP13_T588I, Spike_A243del, NS3_S171L |
| 284 | NR27 | EPI_ISL_1533850 | E | B.1.351 | GH | Mar-21 | NSP3_T1251I, Spike_D215G, E_P71L, NSP3_K837N, Spike_K417N, Spike_L244del, NSP6_G107del, NS8_I121L, NSP6_S106del, Spike_E484K, N_T205I, NSP6_F108del, Spike_L242del, NS3_Q57H, Spike_A701V, Spike_D80A, Spike_N501Y, NSP12_P323L, NSP5_K90R, Spike_D614G, Spike_A243del, NS3_S171L |
| 285 | NR25 | EPI_ISL_1533849 | E | B.1.1.7 | GRY | Mar-21 | Spike_H69del, NS8_Q27stop, NSP3_T183I, Spike_T716I, NS8_K68stop, NSP6_S106del, N_R203K, Spike_A570D, Spike_N501Y, NSP3_I1412T, NS8_R52I, Spike_P681H, Spike_Y144del, NSP6_G107del, NSP3_A890D, Spike_D1118H, NSP6_F108del, NSP13_S38L, NS8_Y73C, N_G204R, Spike_V70del, NSP12_P323L, Spike_D614G, NSP6_L37F, N_D3L, Spike_S982A |
| 286 | CDRW10 | EPI_ISL_1533818 | E | B.1.411 | O | Mar-21 | NSP12_D445G, NSP12_M666I, NS8_Q18stop, Spike_H1159Y, N_T205I, NSP2_T166I, NS3_Q57H, NSP2_T85I, NSP12_P323L, Spike_D614G, NSP6_L37F |
| 287 | CDRW11 | EPI_ISL_1533819 | E | B.1.411 | O | Mar-21 | NSP12_D445G, NSP12_M666I, NS8_Q18stop, Spike_H1159Y, N_T205I, NSP2_T166I, NSP12_P323L, Spike_D614G, NSP6_L37F |
| 288 | CDRW12 | EPI_ISL_1533820 | E | B.1.411 | G | Mar-21 | NSP12_D445G, NSP12_M666I, Spike_H1159Y, NSP2_T166I, NSP12_P323L, Spike_D614G, NSP6_L37F |
| 289 | NR17 | EPI_ISL_1582413 | E | B.1.1.365 | G | Mar-21 | N_L139F, NSP3_T1189I, N_S194L, Spike_E484K, NSP3_A1165V, NSP12_D92G, NSP12_P323L, Spike_D614G, NS3_G100C |
| 290 | NR26 | EPI_ISL_1582415 | E | B.1.351 | GH | Mar-21 | NSP3_T1251I, E_P71L, NSP3_K837N, Spike_K417N, NS8_I121L, NSP6_S106del, Spike_E484K, Spike_A701V, NSP3_V1933P, NSP3_V1935R, NSP3_T1938N, Spike_A243del, Spike_L244del, NSP6_G107del, N_T205I, NSP6_F108del, Spike_L242del, NS3_Q57H, NSP3_K1939I, Spike_D80A, NSP12_P323L, NSP5_K90R, Spike_D614G, NSP13_T588I, NSP3_V1936K, NS3_S171L |
| 291 | CP108816 | EPI_ISL_1533843 | E | B.1.411 | GH | Mar-21 | NSP12_D445G, NSP12_M666I, NS8_Q18stop, Spike_H1159Y, N_T205I, NSP2_T166I, NS3_Q57H, NSP2_T85I, Spike_D287N, NSP12_P323L, Spike_D614G, NSP6_L37F |

| | | | | | | | |
|---|---|---|---|---|---|---|---|
| 292 | CP108841 | EPI_ISL_1533844 | E | B.1.411 | GH | Mar-21 | NSP12_M666I, NS8_Q18stop, Spike_H1159Y, N_T205I, NSP2_T166I, NS3_Q57H, NSP2_T85I, Spike_D287N, NSP5_K90R, Spike_D614G, NSP6_L37F |
| 293 | CP108864 | EPI_ISL_1533845 | E | B.1.411 | G | Mar-21 | NSP12_D445G, NSP12_M666I, NS8_Q18stop, Spike_H1159Y, N_T205I, NSP2_T166I, Spike_D287N, NSP12_P323L, Spike_D614G, NSP6_L37F |
| 294 | CM109233 | EPI_ISL_1582410 | E | B.1.411 | G | Mar-21 | NSP12_D445G, N_K373R, NSP12_M666I, Spike_H1159Y, N_T205I, NSP2_T166I, NSP12_P323L, Spike_D614G, NSP6_L37F |
| 295 | CMC87762 | EPI_ISL_1582412 | E | B.1 | G | Mar-21 | NSP12_D445G, NS8_Q18stop, Spike_H1159Y, N_T205I, NSP2_T166I, NS3_L108F, NS8_R52I, NSP12_P323L, Spike_D614G, NSP6_L37F, Spike_S640F |
| 296 | NR28 | EPI_ISL_1970416 | E | B.1.351 | GH | Mar-21 | NSP3_T1251I, E_P71L, NSP3_K837N, Spike_K417N, NS8_I121L, NSP6_S106del, Spike_E484K, NSP13_P78S, Spike_N501Y, Spike_A243del, Spike_D215G, Spike_L244del, NSP6_G107del, N_T205I, NSP6_F108del, Spike_L242del, NS3_Q57H, NSP2_T85I, Spike_D80A, NSP12_P323L, NSP5_K90R, Spike_D614G, NSP13_T588I, NS3_S171L |
| 297 | CDR72 | EPI_ISL_1970385 | E | B.1.411 | O | Mar-21 | NSP12_M666I, Spike_H1159Y, NSP2_T166I, NSP1_G30S, NSP12_P323L, NSP5_K90R, Spike_D614G, NSP6_L37F |
| 298 | CDR71 | EPI_ISL_1970384 | E | B.1.428 | GH | Mar-21 | NS8_W45C, NSP12_D445G, NSP12_M666I, NS8_Q18stop, Spike_H1159Y, NS3_L53F, N_T205I, NSP2_T166I, NS3_Q57H, NSP1_G30S, NSP2_T85I, NSP3_S1670F, NSP12_P323L, NSP5_K90R, Spike_D614G, NSP6_L37F |
| 299 | NR29 | EPI_ISL_1970417 | E | B.1.1.7 | O | Mar-21 | NS8_Q27stop, NSP3_A890D, NSP6_G107del, Spike_T716I, NS8_K68stop, NSP6_S106del, N_R203K, Spike_A570D, Spike_D1118H, NS3_W131C, NSP6_F108del, NS8_Y73C, N_G204R, Spike_N501Y, NSP3_I1412T, NS8_R52I, NSP12_P323L, Spike_P681H, Spike_D614G, Spike_Y144del, N_D3L, Spike_S982A |
| 300 | CDR38 | EPI_ISL_1970355 | E | B.1.1.7 | GR | Apr-21 | Spike_H69del, NS8_Q27stop, NSP3_T183I, Spike_T716I, NS8_K68stop, NSP6_S106del, N_R203K, Spike_A570D, NSP3_I1412T, NS8_R52I, Spike_P681H, Spike_Y144del, NSP6_G107del, NSP3_A890D, Spike_D1118H, NSP6_F108del, N_G204R, Spike_V70del, NS3_E194D, NSP12_P323L, Spike_D614G, N_D3L, Spike_S982A, N_S235F |
| 301 | CDR39 | EPI_ISL_1970356 | E | B.1.1.7 | GR | Apr-21 | NS8_Q27stop, NSP3_T183I, NSP8_T145I, Spike_T716I, NS8_K68stop, NSP6_S106del, N_R203K, Spike_A570D, Spike_N501Y, NSP3_I1412T, NS8_R52I, Spike_P681H, Spike_Y144del, NSP6_G107del, NSP3_A890D, Spike_D1118H, NSP6_F108del, NS8_Y73C, N_G204R, NS3_E194D, NSP12_P323L, Spike_D614G, N_D3L, Spike_S982A, N_S235F |
| 302 | CDR40 | EPI_ISL_1970357 | E | B.1.1.7 | GRY | Apr-21 | Spike_H69del, NS8_Q27stop, NSP3_T183I, NSP8_T145I, Spike_T716I, NS8_K68stop, NSP6_S106del, N_R203K, Spike_A570D, Spike_N501Y, NSP3_I1412T, NS8_R52I, Spike_P681H, Spike_Y144del, NSP6_G107del, NSP3_A890D, Spike_D1118H, NSP6_F108del, NS8_Y73C, N_G204R, Spike_V70del, NS3_E194D, NSP12_P323L, Spike_D614G, N_D3L, Spike_S982A |
| 303 | CDR41 | EPI_ISL_1970358 | E | B.1.1.7 | GR | Apr-21 | NS8_Q27stop, NSP3_T183I, NSP8_T145I, NS8_K68stop, NSP6_S106del, N_R203K, Spike_A570D, Spike_N501Y, NSP3_I1412T, NS8_R52I, Spike_P681H, Spike_Y144del, NSP6_G107del, NSP3_A890D, Spike_D1118H, NSP6_F108del, NS8_Y73C, N_G204R, NS3_E194D, NSP12_P323L, Spike_D614G, NSP2_S430L, N_D3L, Spike_S982A, N_S235F |

| | | | | | | | |
|---|---|---|---|---|---|---|---|
| 304 | CDR56 | EPI_ISL_1970370 | E | B.1.411 | O | Apr-21 | NS8_W45C, NSP3_K429N, NSP12_D445G, NSP3_G145C, NSP12_M666I, NS8_Q18stop, Spike_H1159Y, NS3_L53F, N_T205I, NSP2_T166I, NS3_Q57H, NSP2_T85I, NSP3_S1670F, NSP5_K90R, Spike_D614G, NSP6_L37F, NSP12_Y546C, Spike_G261V |
| 305 | CDR57 | EPI_ISL_1970371 | E | B.1.411 | G | Apr-21 | NS8_W45C, NSP3_K429N, NSP12_D445G, NSP12_M666I, Spike_H1159Y, NSP3_K945N, N_T205I, NSP2_T166I, NSP2_T85I, NSP3_S1670F, NSP12_P323L, NSP5_K90R, Spike_D614G, NSP6_L37F |
| 306 | CDR59 | EPI_ISL_1970373 | E | B.1.411 | G | Apr-21 | NS8_W45C, NSP3_K429N, NSP12_D445G, NSP12_M666I, NS8_Q18stop, Spike_H1159Y, N_T205I, NSP2_T166I, NSP2_T85I, NSP9_G37R, NSP3_S1670F, NSP12_P323L, NSP5_K90R, Spike_D614G, NSP6_L37F |
| 307 | CDR58 | EPI_ISL_1970372 | E | B.1.428 | GH | Apr-21 | NS8_W45C, NSP3_K429N, NSP12_D445G, NSP12_M666I, Spike_H1159Y, NS3_L53F, N_T205I, NSP2_T166I, NS3_Q57H, NSP3_S1670F, NSP12_P323L, NSP5_K90R, Spike_D614G, NSP6_L37F |
| 308 | NR44 | EPI_ISL_1970418 | E | B.1.617.2 | G | Apr-21 | NS7a_L116F, N_D63G, N_R203M, NSP12_G671S, Spike_G142D, NSP2_P129L, NS3_S26L, NSP14_P46L, NSP2_R246H, Spike_P681R, Spike_R158del, NS7a_V82A, Spike_F157del, Spike_T19R, NS7a_T120I, M_I82T, NSP6_V149A, Spike_D950N, NSP3_P822L, NSP13_P77L, Spike_E156G, NSP4_A446V, NSP12_P323L, Spike_D614G, Spike_L452R, NS7a_T11K |
| 309 | CDR42 | EPI_ISL_1970359 | F | B.1.1.7 | O | Apr-21 | Spike_H69del, NS8_Q27stop, NSP3_T183I, Spike_T716I, NS8_K68stop, NSP6_S106del, N_R203K, NSP3_D110Y, Spike_A570D, NSP15_E260A, Spike_N501Y, NSP3_I1412T, NS8_R52I, NSP8_S76F, Spike_P681H, Spike_Y144del, NSP6_G107del, NSP3_A890D, Spike_D1118H, NSP6_F108del, N_G204R, Spike_V70del, NSP12_P323L, Spike_D614G, N_D3L, Spike_S982A |
| 310 | CMC88881 | EPI_ISL_1972232 | F | B.1.1.7 | GR | Apr-21 | Spike_H69del, NS8_Q27stop, NSP3_T183I, Spike_T716I, NSP6_S106del, N_R203K, NSP3_D110Y, NSP15_E260A, NSP3_R1297del, NSP3_I1412T, NS8_R52I, NSP8_S76F, Spike_P681H, NSP3_V1298del, NSP6_G107del, NSP3_A890D, Spike_D1118H, NSP6_F108del, N_G204R, Spike_V70del, NSP3_S1296del, Spike_N149del, NSP12_P323L, Spike_D614G, N_D3L, Spike_S982A, N_S235F |
| 311 | CMC88939 | EPI_ISL_1970412 | F | B.1.1.7 | GRY | Apr-21 | Spike_H69del, NS8_Q27stop, NSP3_T183I, NS8_K68stop, NSP6_S106del, N_R203K, NSP3_D110Y, Spike_A570D, NSP15_E260A, Spike_N501Y, NSP3_R1297del, NSP3_L689F, NSP3_I1412T, NSP8_S76F, Spike_P681H, Spike_Y144del, NS7b_S31L, NSP3_V1298del, NSP3_A890D, NSP6_G107del, Spike_D1118H, NSP6_F108del, NS8_Y73C, N_G204R, Spike_V70del, NSP3_S1296del, NSP12_P323L, Spike_D614G, N_D3L, Spike_S982A, N_S235F |
| 312 | CMC89508 | EPI_ISL_1972230 | F | B.1.1.7 | GR | Apr-21 | NS8_Q27stop, NSP3_T183I, Spike_T716I, NS8_K68stop, Spike_N370Y, NSP6_S106del, N_R203K, NSP14_A320V, Spike_N501Y, NSP3_I1412T, NS8_R52I, Spike_P681H, Spike_Y144del, Spike_ins370KLVPFWstopSF, NSP6_G107del, NSP3_A890D, Spike_D1118H, NSP6_F108del, NS8_Y73C, NS3_E194D, NSP12_P323L, Spike_D614G, N_D3L, Spike_S982A |

| | | | | | | | |
|---|---|---|---|---|---|---|---|
| 313 | CDR104 | EPI_ISL_1970352 | F | B.1.1.7 | GRY | Apr-21 | Spike_H69del, NS8_Q27stop, NSP3_T183I, NSP8_T145I, Spike_T716I, NS8_K68stop, NSP6_S106del, N_R203K, Spike_A570D, Spike_N501Y, NSP3_I1412T, NS8_R52I, Spike_P681H, Spike_Y144del, NSP6_G107del, NSP3_A890D, Spike_D1118H, NSP6_F108del, NS8_Y73C, N_G204R, Spike_V70del, NS3_E194D, NSP12_P323L, Spike_D614G, N_D3L, Spike_S982A, N_S235F |
| 314 | CDR105 | EPI_ISL_1970353 | F | B.1.1.7 | GR | Apr-21 | Spike_H69del, NS8_Q27stop, NSP3_T183I, Spike_T716I, NS8_K68stop, NSP3_W1196C, NSP6_S106del, N_R203K, NSP15_E260A, NSP3_I1412T, NS8_R52I, NSP8_S76F, Spike_P681H, Spike_Y144del, NSP6_G107del, NSP3_A890D, Spike_D1118H, NSP6_F108del, NS8_Y73C, N_G204R, Spike_V70del, NSP12_P323L, Spike_D614G, N_D3L, Spike_S982A, N_S235F |
| 315 | CDR106 | EPI_ISL_1970354 | F | B.1.1.7 | GRY | Apr-21 | Spike_H69del, NS8_Q27stop, NSP3_T183I, NSP8_T145I, Spike_T716I, NS8_K68stop, NSP6_S106del, N_R203K, Spike_A570D, Spike_N501Y, NSP3_I1412T, NS8_R52I, Spike_P681H, Spike_Y144del, NSP6_G107del, NSP3_A890D, Spike_D1118H, NSP6_F108del, NS8_Y73C, N_G204R, NSP15_H234Y, Spike_V70del, NS3_E194D, NSP12_P323L, Spike_D614G, N_D3L, Spike_S982A, N_S235F |
| 316 | CDR107 | EPI_ISL_1972227 | F | B.1.1.7 | G | Apr-21 | Spike_H69del, NS8_Q27stop, NSP3_T183I, NSP12_L775M, NSP3_A890D, NS8_K68stop, Spike_D1118H, NSP12_Q773H, NS8_Y73C, Spike_V70del, NS3_E194D, NSP3_I1412T, NS8_R52I, NSP12_P323L, NSP12_ins772LstopA, Spike_P681H, Spike_D614G, Spike_Y144del, NSP12_ins774TLR, N_D3L, Spike_S982A |
| 317 | CDR43 | EPI_ISL_1970360 | F | B.1.1.7 | G | Apr-21 | Spike_H69del, NS8_Q27stop, NSP3_T183I, NSP3_W1196C, NSP6_S106del, NSP3_D110Y, Spike_A570D, NSP15_E260A, NSP3_R1297del, NSP3_I1412T, NSP8_S76F, Spike_P681H, Spike_Y144del, NSP3_V1298del, NSP3_A890D, NSP6_G107del, Spike_D1118H, NSP6_F108del, Spike_V70del, NSP3_S1296del, E_ins38CLX, NSP12_P323L, Spike_D614G, N_D3L, Spike_S982A |
| 318 | CDR44 | EPI_ISL_1970361 | F | B.1.1.7 | O | Apr-21 | NS8_Q27stop, NSP3_T183I, Spike_T716I, NS8_K68stop, NSP6_S106del, N_R203K, NSP3_D110Y, NSP15_E260A, Spike_N501Y, NSP3_R1297del, NSP3_I1412T, NS8_R52I, NSP8_S76F, Spike_P681H, Spike_Y144del, NSP3_V1298del, NSP6_G107del, NSP3_A890D, Spike_D1118H, NSP6_F108del, NS8_Y73C, N_G204R, NSP3_S1296del, NSP12_P323L, Spike_D614G, N_D3L, Spike_S982A, N_S235F |
| 319 | CMC90016 | EPI_ISL_1970413 | F | B.1.1.7 | G | Apr-21 | Spike_H69del, NS8_Q27stop, NSP3_T183I, Spike_T716I, NS8_K68stop, NSP6_S106del, Spike_A570D, NSP15_E260A, Spike_N501Y, NSP3_R1297del, NSP3_I1412T, NS8_R52I, NSP8_S76F, Spike_P681H, Spike_Y144del, NSP3_V1298del, NSP6_G107del, NSP3_A890D, Spike_D1118H, NSP6_F108del, NS8_Y73C, Spike_V70del, NSP3_S1296del, NSP12_P323L, Spike_D614G, N_D3L, Spike_S982A, N_S235F |

| | | | | | | | |
|---|---|---|---|---|---|---|---|
| 320 | CDR45 | EPI_ISL_1970362 | F | B.1.1.7 | GRY | Apr-21 | Spike_H69del, NS8_Q27stop, NSP3_T183I, Spike_T716I, NS8_K68stop, NSP6_S106del, N_R203K, NSP3_D110Y, Spike_A570D, NSP15_E260A, Spike_N501Y, NSP3_I1412T, NS8_R52I, NSP8_S76F, Spike_P681H, Spike_Y144del, NSP6_G107del, NSP3_A890D, Spike_D1118H, NSP6_F108del, NS8_Y73C, N_G204R, Spike_V70del, NSP12_P323L, Spike_D614G, N_D3L, Spike_S982A, N_S235F |
| 321 | CDR46 | EPI_ISL_1970363 | F | B.1.1.7 | GRY | Apr-21 | Spike_H69del, NSP3_T183I, Spike_T716I, NS8_K68stop, NSP6_S106del, N_R203K, NSP3_D110Y, Spike_A570D, NSP15_E260A, NSP3_M560I, Spike_N501Y, NSP3_I1412T, NS8_R52I, NSP8_S76F, Spike_P681H, Spike_Y144del, NSP6_G107del, NSP3_A890D, Spike_D1118H, NSP6_F108del, NS8_Y73C, N_G204R, Spike_V70del, NSP12_P323L, Spike_D614G, N_D3L, Spike_S982A, N_S235F |
| 322 | CDR47 | EPI_ISL_1972233 | F | B.1.1.7 | GR | Apr-21 | Spike_H69del, NS8_Q27stop, NSP3_T183I, Spike_T716I, NSP12_Q5H, NSP3_W1196C, NSP6_S106del, N_R203K, NSP3_D110Y, Spike_A570D, NSP15_E260A, NSP12_F7L, NSP12_N9I, Spike_N501Y, NSP3_R1297del, NSP12_S6stop, NSP3_I1412T, NS8_R52I, NSP8_S76F, Spike_P681H, NSP12_V11M, NSP3_V1298del, NSP6_G107del, NSP3_A890D, Spike_D1118H, NSP6_F108del, NS8_Y73C, N_G204R, NSP12_R10A, Spike_V70del, NSP12_G13T, NSP3_S1296del, NSP12_L8K, NSP12_P323L, NSP12_V14R, Spike_D614G, N_D3L, Spike_S982A, N_S235F |
| 323 | CDR48 | EPI_ISL_1970364 | F | B.1.1.7 | O | Apr-21 | NS8_Q27stop, NSP3_T183I, Spike_T716I, NS8_K68stop, NSP3_W1196C, NSP6_S106del, N_R203K, NSP3_D110Y, Spike_A570D, NSP15_E260A, Spike_N501Y, NSP3_I1412T, NS8_R52I, NSP8_S76F, Spike_P681H, Spike_Y144del, NSP6_G107del, NSP3_A890D, Spike_D1118H, NSP6_F108del, NS8_Y73C, N_G204R, NSP12_P323L, Spike_D614G, N_D3L, Spike_S982A |
| 324 | CDR49 | EPI_ISL_1970365 | F | B.1.1.7 | O | Apr-21 | Spike_R21I, NS8_Q27stop, Spike_T716I, NS8_K68stop, NSP3_W1196C, NSP6_S106del, N_R203K, NSP3_D110Y, Spike_A570D, NSP15_E260A, Spike_N501Y, NSP3_I1412T, NS8_R52I, NSP8_S76F, Spike_P681H, Spike_Y144del, Spike_Q14H, NS3_R134C, NSP6_G107del, NSP3_A890D, Spike_D1118H, NSP6_F108del, NS8_Y73C, N_G204R, NSP12_P323L, Spike_D614G, N_D3L, Spike_S982A |
| 325 | CDR50 | EPI_ISL_1970366 | F | B.1.1.7 | GRY | Apr-21 | Spike_H69del, NS8_Q27stop, NSP3_T183I, NSP8_T145I, Spike_T716I, NS8_K68stop, NSP6_S106del, N_R203K, Spike_A570D, Spike_N501Y, NSP3_I1412T, NS8_R52I, Spike_P681H, Spike_Y144del, NSP15_G246C, NSP6_G107del, NSP3_A890D, Spike_D1118H, NSP6_F108del, NS8_Y73C, N_G204R, Spike_V70del, NS3_E194D, NSP12_P323L, Spike_D614G, N_D3L, Spike_S982A, N_S235F |

| | | | | | | | |
|---|---|---|---|---|---|---|---|
| 326 | CDR51 | EPI_ISL_1972229 | F | B.1.1.7 | GR | Apr-21 | Spike_H69del, NS8_Q27stop, NSP3_T183I, Spike_T716I, NS8_K68stop, NSP6_S106del, N_R203K, NSP3_D110Y, Spike_A570D, NSP15_E260A, NSP2_K110R, Spike_ins370LVPFWstopSF, Spike_N501Y, NSP3_R1297del, NSP3_I1412T, NS8_R52I, NSP8_S76F, Spike_P681H, NSP3_V1298del, NSP6_G107del, NSP3_A890D, Spike_D1118H, NSP6_F108del, N_G204R, Spike_V70del, NSP3_S1296del, NSP12_P323L, Spike_D614G, Spike_S982A, Spike_N370K, N_S235F |
| 327 | CDR53 | EPI_ISL_1970368 | F | B.1.1.7 | GR | Apr-21 | NSP3_T183I, Spike_T716I, NS8_K68stop, NSP6_S106del, N_R203K, NSP4_A231V, NS3_T89I, NSP15_E260A, Spike_N501Y, NSP3_I1412T, Spike_P681H, Spike_Y144del, NSP12_P227L, NSP6_G107del, NSP3_A890D, NSP3_D782N, Spike_D1118H, NSP6_F108del, NS8_Y73C, N_G204R, NSP12_P323L, Spike_D614G, N_D3L, Spike_S982A |
| 328 | CDR54 | EPI_ISL_1972234 | F | B.1.1.7 | O | Apr-21 | Spike_H69del, NS8_Q27stop, NSP3_T183I, Spike_T716I, NS8_K68stop, NSP6_S106del, Spike_A570D, NSP15_E260A, NSP3_I1412T, NS8_R52I, NSP8_S76F, Spike_P681H, Spike_Y144del, NS7a_T14I, NSP6_G107del, NSP3_A890D, Spike_D1118H, NSP6_F108del, NS8_Y73C, NSP3_T779I, Spike_V70del, NSP12_P323L, Spike_D614G, N_D3L, Spike_S982A |
| 329 | CDR55 | EPI_ISL_1970369 | F | B.1.1.7 | GRY | Apr-21 | Spike_H69del, NS8_Q27stop, NSP3_T183I, Spike_T716I, NS8_K68stop, NSP6_S106del, N_R203K, NSP3_D110Y, Spike_A570D, NSP15_E260A, Spike_N501Y, NSP3_I1412T, NS8_R52I, NSP8_S76F, Spike_P681H, Spike_Y144del, NS7a_T28I, NSP6_G107del, NSP3_A890D, Spike_D1118H, NSP6_F108del, NS8_Y73C, N_G204R, Spike_V70del, NSP12_P323L, Spike_D614G, N_D3L, Spike_S982A |
| 330 | CDR60 | EPI_ISL_1970374 | F | B.1.1.7 | GRY | Apr-21 | Spike_H69del, NS8_Q27stop, NSP3_T183I, Spike_T716I, NS8_K68stop, NSP6_S106del, N_R203K, Spike_A570D, NSP15_E260A, Spike_N501Y, NSP3_I1412T, NS8_R52I, NSP8_S76F, Spike_P681H, Spike_Y144del, NSP6_G107del, NSP3_A890D, Spike_D1118H, NSP6_F108del, NS8_Y73C, NSP12_A699S, N_G204R, Spike_V70del, NSP12_P323L, Spike_D614G, N_D3L, Spike_S982A |
| 331 | CDR61 | EPI_ISL_1970375 | F | B.1.1.7 | GRY | Apr-21 | Spike_H69del, NS8_Q27stop, NSP3_T183I, NSP8_T145I, Spike_T716I, NS8_K68stop, NSP6_S106del, N_R203K, Spike_A570D, Spike_N501Y, NS6_W27L, NS8_R52I, Spike_P681H, Spike_Y144del, NSP6_G107del, NSP3_A890D, N_P46S, Spike_D1118H, NSP6_F108del, NS8_Y73C, N_G204R, Spike_V70del, NS3_E194D, Spike_D614G, N_D3L, Spike_S982A |
| 332 | CDR62 | EPI_ISL_1970376 | F | B.1.1.7 | O | Apr-21 | Spike_H69del, NS8_Q27stop, NSP3_T183I, Spike_T716I, NS8_K68stop, NSP6_S106del, N_R203K, NSP3_D110Y, Spike_A570D, NSP15_E260A, NSP2_K110R, Spike_N501Y, NSP3_I1412T, NS8_R52I, NSP8_S76F, Spike_P681H, Spike_Y144del, NSP6_G107del, NSP3_A890D, Spike_D1118H, NSP6_F108del, NS8_Y73C, N_G204R, Spike_V70del, NSP12_P323L, Spike_D614G, N_D3L, Spike_S982A, N_S235F |

| | | | | | | | |
|---|---|---|---|---|---|---|---|
| 333 | CDR63 | EPI_ISL_1970377 | F | B.1.1.7 | GRY | Apr-21 | Spike_H69del, NS8_Q27stop, NSP3_T183I, Spike_T716I, NS8_K68stop, NSP6_S106del, N_R203K, NSP15_E260A, NSP2_K110R, Spike_N501Y, NSP3_I1412T, NS8_R52I, NSP8_S76F, Spike_P681H, Spike_Y144del, NSP6_G107del, NSP3_A890D, Spike_D1118H, NSP6_F108del, NS8_Y73C, N_G204R, Spike_V70del, NSP12_P323L, Spike_D614G, N_D3L, Spike_S982A |
| 334 | CDR79 | EPI_ISL_1970392 | F | B.1.1.7 | GRY | Apr-21 | Spike_H69del, NS8_Q27stop, NSP3_T183I, Spike_T716I, NS8_K68stop, NSP6_S106del, N_R203K, NSP4_A231V, Spike_A570D, NS3_T89I, Spike_N501Y, NSP3_I1412T, NS8_R52I, Spike_Y144del, NSP12_P227L, Spike_L141F, NSP3_P67L, NSP6_G107del, NSP3_A890D, Spike_D1118H, NSP6_F108del, NS8_Y73C, N_G204R, NSP3_T275A, Spike_V70del, NSP12_P323L, NSP14_P451S, Spike_D614G, N_D3L, Spike_S982A, N_S235F |
| 335 | CDR52 | EPI_ISL_1970367 | F | B.1.411 | O | Apr-21 | NSP12_D445G, N_D144N, NSP12_M666I, NSP6_L125F, Spike_H1159Y, N_T205I, NSP2_T166I, NSP2_T85I, NSP12_P323L, Spike_D614G, NSP6_L37F |
| 336 | CDR68 | EPI_ISL_1970381 | F | B.1.1.7 | G | Apr-21 | Spike_H69del, NS8_Q27stop, NSP3_T183I, NSP8_T145I, Spike_T716I, NSP6_S106del, Spike_A570D, NSP14_A320V, Spike_N501Y, NSP3_I1412T, NS8_R52I, Spike_P681H, Spike_Y144del, NSP6_G107del, NSP3_A890D, Spike_D1118H, NSP6_F108del, NS8_Y73C, Spike_V70del, NS3_E194D, NSP12_P323L, Spike_D614G, N_D3L, Spike_S982A |
| 337 | CDR67 | EPI_ISL_1970380 | F | B.1.1.7 | O | Apr-21 | Spike_H69del, NS8_Q27stop, NSP3_T183I, NSP8_T145I, Spike_T716I, NS8_K68stop, NSP6_S106del, N_R203K, Spike_A570D, Spike_N501Y, NSP3_I1412T, NS8_R52I, Spike_P681H, NS7a_R78C, Spike_Y144del, NSP6_G107del, NSP3_A890D, Spike_D1118H, NSP6_F108del, NS8_Y73C, N_G204R, Spike_V70del, NS3_E194D, NSP12_P323L, Spike_D614G, N_D3L, Spike_S982A |
| 338 | CDR74 | EPI_ISL_1970387 | F | B.1.1.7 | GRY | Apr-21 | Spike_H69del, NS8_Q27stop, NSP3_T183I, NSP8_T145I, Spike_T716I, NS8_K68stop, NSP6_S106del, N_R203K, Spike_A570D, Spike_N501Y, NSP3_I1412T, NS8_R52I, Spike_P681H, NS7a_R78C, Spike_Y144del, NSP6_G107del, NSP3_A890D, Spike_D1118H, NSP6_F108del, NS8_Y73C, N_G204R, Spike_V70del, NS3_E194D, NSP12_P323L, Spike_D614G, N_D3L, Spike_S982A, N_S235F |
| 339 | CDR75 | EPI_ISL_1970388 | F | B.1.1.7 | GRY | Apr-21 | Spike_H69del, NS8_Q27stop, Spike_T716I, NS8_K68stop, NSP6_S106del, N_R203K, Spike_A570D, Spike_N501Y, NSP3_I1412T, NS8_R52I, Spike_P681H, NS7a_R78C, Spike_Y144del, NSP6_G107del, Spike_D1118H, NSP6_F108del, NS8_Y73C, N_G204R, Spike_V70del, NS3_E194D, NSP12_P323L, Spike_D614G, Spike_S982A |
| 340 | CDR76 | EPI_ISL_1970389 | F | B.1.1.7 | O | Apr-21 | Spike_H69del, NS8_Q27stop, NSP3_T183I, NSP8_T145I, Spike_T716I, NS8_K68stop, NSP6_S106del, N_R203K, Spike_N501Y, NSP3_I1412T, NS8_R52I, Spike_P681H, Spike_Y144del, NSP6_G107del, NSP3_A890D, Spike_D1118H, NSP6_F108del, NS8_Y73C, N_G204R, Spike_V70del, NS3_E194D, NSP12_P323L, Spike_D614G, Spike_S982A, N_S235F |
| 341 | CDR77 | EPI_ISL_1970390 | F | B.1.1.7 | GR | Apr-21 | NSP6_G107del, Spike_T716I, NSP6_S106del, N_R203K, Spike_A570D, Spike_D1118H, NSP6_F108del, N_G204R, NS3_E194D, NSP3_I1412T, Spike_P681H, Spike_D614G, N_D3L, Spike_S982A, N_S235F |

| | | | | | | | |
|---|---|---|---|---|---|---|---|
| 342 | CDR78 | EPI_ISL_1970391 | F | B.1.1.7 | GRY | Apr-21 | Spike_H69del, NS8_Q27stop, NSP3_T183I, NSP8_T145I, NS8_K68stop, NSP6_S106del, N_R203K, Spike_A570D, Spike_N501Y, NSP3_I1412T, NS8_R52I, Spike_P681H, NS7a_R78C, Spike_Y144del, NSP6_G107del, NSP3_A890D, NSP6_F108del, NS8_Y73C, N_G204R, Spike_V70del, NS3_E194D, NSP12_P323L, Spike_D614G, N_D3L, Spike_S982A |
| 343 | CDR90 | EPI_ISL_1970403 | F | B.1.1.7 | GRY | Apr-21 | Spike_H69del, NS8_Q27stop, NSP3_T183I, NSP8_T145I, Spike_T716I, NS8_K68stop, NSP6_S106del, N_R203K, Spike_A570D, Spike_N501Y, NSP3_I1412T, NS8_R52I, Spike_P681H, Spike_Y144del, NSP6_G107del, NSP3_A890D, NS7a_P99S, Spike_D1118H, NSP6_F108del, NS8_Y73C, N_G204R, Spike_V70del, NS3_E194D, NSP12_P323L, Spike_D614G, N_D3L, Spike_S982A, N_S235F |
| 344 | CDR91 | EPI_ISL_1970404 | F | B.1.1.7 | G | Apr-21 | Spike_H69del, NS8_Q27stop, NSP3_T183I, Spike_T716I, NSP6_S106del, NSP3_D110Y, NSP15_E260A, Spike_N501Y, NSP3_I1412T, NS8_R52I, NSP8_S76F, Spike_P681H, Spike_Y144del, NSP6_G107del, NSP3_A890D, Spike_D1118H, NSP6_F108del, NS8_Y73C, Spike_V70del, NSP12_P323L, Spike_D614G, N_D3L, Spike_S982A, N_S235F |
| 345 | CDR92 | EPI_ISL_1970405 | F | B.1.1.7 | GR | Apr-21 | NS8_Q27stop, NSP3_V1298del, NSP3_A890D, NSP6_G107del, Spike_T716I, NS8_K68stop, NSP6_S106del, N_R203K, Spike_A570D, Spike_D1118H, NSP6_F108del, NSP15_E260A, N_G204R, Spike_M153del, NSP3_S1296del, NSP3_R1297del, NS8_R52I, NSP8_S76F, Spike_P681H, Spike_D614G, N_D3L, Spike_S982A |
| 346 | CDR93 | EPI_ISL_1970406 | F | B.1.1.7 | O | Apr-21 | Spike_H69del, NS8_Q27stop, Spike_T716I, NS8_K68stop, NSP3_W1196C, NSP6_S106del, N_R203K, NSP3_D110Y, Spike_A570D, NSP15_E260A, Spike_N501Y, NSP3_I1412T, NS8_R52I, NSP8_S76F, Spike_P681H, Spike_Y144del, NSP6_G107del, NSP3_A890D, Spike_D1118H, NSP6_F108del, NS8_Y73C, N_G204R, Spike_V70del, NSP12_P323L, Spike_D614G, N_D3L, Spike_S982A, N_S235F |
| 347 | CDR94 | EPI_ISL_1970407 | F | B.1.1.7 | GRY | Apr-21 | Spike_H69del, NS8_Q27stop, NSP3_T183I, Spike_T716I, NS8_K68stop, NSP3_W1196C, NSP6_S106del, N_R203K, NSP3_D110Y, Spike_A570D, NSP15_E260A, Spike_N501Y, NSP3_R1297del, NSP3_I1412T, NS8_R52I, NSP8_S76F, Spike_P681H, Spike_Y144del, NSP3_V1298del, NSP6_G107del, NSP3_A890D, Spike_D1118H, NSP6_F108del, NS8_Y73C, N_G204R, Spike_V70del, NSP3_S1296del, NSP12_P323L, Spike_D614G, N_D3L, Spike_S982A, N_S235F |
| 348 | CDR95 | EPI_ISL_1970408 | F | B.1.1.7 | GR | Apr-21 | Spike_H69del, NS8_Q27stop, NSP3_T183I, Spike_T716I, NS8_K68stop, NSP3_W1196C, NSP6_S106del, N_R203K, NSP3_D110Y, Spike_A570D, NSP15_E260A, Spike_N501Y, NSP3_I1412T, NS8_R52I, NSP8_S76F, Spike_P681H, NSP6_G107del, NSP3_A890D, Spike_D1118H, NSP6_F108del, NS8_Y73C, N_G204R, Spike_V70del, Spike_D614G, N_D3L, Spike_S982A |

| | | | | | | | |
|---|---|---|---|---|---|---|---|
| 349 | CDR96 | EPI_ISL_1970409 | F | B.1.1.7 | GRY | Apr-21 | Spike_H69del, NS8_Q27stop, NSP3_T183I, Spike_T716I, NS8_K68stop, NSP6_S106del, N_R203K, Spike_A570D, NSP15_E260A, NSP2_K110R, NSP3_I1412T, NS8_R52I, NSP8_S76F, Spike_P681H, Spike_Y144del, NSP6_G107del, NSP3_A890D, Spike_D1118H, NSP6_F108del, NS8_Y73C, N_G204R, Spike_V70del, NSP9_T21I, NSP12_P323L, Spike_D614G, N_D3L, Spike_S982A, N_S235F |
| 350 | CDR97 | EPI_ISL_1972228 | F | B.1.1.7 | G | Apr-21 | Spike_H69del, NS8_Q27stop, NSP15_R138L, NSP3_T183I, NS8_K68stop, NSP6_S106del, NSP3_D110Y, NSP15_E260A, NSP5_L67F, NSP3_R1297del, NSP3_I1412T, NS8_R52I, NSP8_S76F, Spike_P681H, Spike_Y144del, NSP3_V1298del, NSP3_A890D, NSP6_G107del, Spike_D1118H, NSP6_F108del, NS8_Y73C, Spike_V70del, NSP3_S1296del, NSP12_P323L, Spike_D614G, N_D3L, Spike_S982A, NSP3_S699F |
| 351 | CMC91808 | EPI_ISL_1970414 | F | B.1.1.7 | G | Apr-21 | Spike_H69del, NS8_Q27stop, NSP3_T183I, NSP8_T145I, Spike_T716I, E_L21F, NS8_K68stop, NSP6_S106del, Spike_A570D, NSP14_A320V, Spike_N501Y, NSP3_I1412T, NS8_R52I, Spike_P681H, Spike_Y144del, NSP6_G107del, NSP3_A890D, Spike_D1118H, NSP6_F108del, NS8_Y73C, Spike_V70del, NS3_E194D, NSP12_P323L, Spike_D614G, N_D3L, Spike_S982A |
| 352 | CMC95123 | EPI_ISL_1972226 | F | B.1.1.7 | O | Apr-21 | NS3_D250E, Spike_H69del, NS8_Q27stop, NSP6_S106del, N_R203K, Spike_A570D, NSP3_G277F, NSP3_A274K, NS8_R52I, NSP3_N276M, Spike_P681H, Spike_Y144del, E_K63T, NSP2_G235C, NSP3_T275P, NSP3_A890D, NSP6_G107del, NSP6_F108del, NSP3_D1208E, N_G204R, E_N64A, Spike_V70del, NSP3_I273L, NS3_E194D, NSP12_P323L, Spike_D614G, NSP3_K280N, NSP3_Y272H, N_D3L, NSP3_P278T, NSP3_V281M, N_S235F |
| 353 | CDR66 | EPI_ISL_1970379 | F | B.1.411 | GH | Apr-21 | Spike_N679K, NSP12_D445G, NSP12_M666I, NS8_Q18stop, Spike_H1159Y, N_T205I, NS3_Q57H, NSP12_P323L, Spike_D614G, NSP6_L37F, Spike_V1040F |
| 354 | CDR70 | EPI_ISL_1970383 | F | B.1.411 | O | Apr-21 | NS3_V255del, NS8_E59stop, NSP12_D445G, NSP3_P822L, NSP12_M666I, NS8_Q18stop, Spike_H1159Y, N_T205I, NS3_Q57H, NSP14_P203L, Spike_L5F, NSP2_T85I, NS3_W193L, NSP12_P323L, Spike_D614G, NSP6_L37F, Spike_G1167R, Spike_V1122L |
| 355 | CDR100 | EPI_ISL_1970350 | F | B.1.1.7 | GRY | Apr-21 | Spike_H69del, NS8_Q27stop, NSP3_T183I, Spike_T716I, NS8_K68stop, NSP6_S106del, N_R203K, NSP4_A231V, Spike_A570D, NS3_T89I, NSP13_L581F, NSP3_I1412T, NS8_R52I, Spike_P681H, Spike_Y144del, NSP12_P227L, NSP3_P67L, NSP6_G107del, NSP3_A890D, NSP3_D782N, Spike_D1118H, NSP6_F108del, NS8_Y73C, N_G204R, NSP3_T275A, Spike_V70del, NSP5_A193V, NSP12_P323L, NSP14_P451S, Spike_D614G, N_D3L, Spike_S982A, N_S235F |
| 356 | CDR65 | EPI_ISL_1970378 | F | B.1.1.7 | G | Apr-21 | NSP8_E20K, NSP3_T183I, NSP8_T145I, NSP3_A890D, NSP6_G107del, Spike_T716I, NS8_K68stop, NSP6_S106del, Spike_D1118H, NSP6_F108del, NS8_Y73C, Spike_N501Y, NS3_E194D, NSP3_I1412T, NS8_R52I, NSP12_P323L, Spike_D614G, Spike_Y144del, N_D3L, Spike_S982A, N_S235F |

| | | | | | | | |
|---|---|---|---|---|---|---|---|
| 357 | CDR69 | EPI_ISL_1970382 | F | B.1.1.7 | O | Apr-21 | Spike_H69del, NSP3_T183I, NSP8_T145I, Spike_T716I, NS8_K68stop, NSP6_S106del, N_R203K, Spike_A570D, Spike_N501Y, NSP3_I1412T, Spike_P681H, Spike_Y144del, NSP6_G107del, NSP3_A890D, Spike_D1118H, NSP6_F108del, NS8_Y73C, N_G204R, Spike_V70del, NS3_E194D, NSP12_P323L, Spike_D614G, Spike_S982A |
| 358 | CDR73 | EPI_ISL_1970386 | F | B.1.1.7 | O | Apr-21 | Spike_H69del, NS8_Q27stop, NSP3_T183I, Spike_T716I, NS8_K68stop, NSP6_S106del, N_R203K, Spike_A570D, NSP14_A320V, Spike_N501Y, NSP3_I1412T, NS8_R52I, Spike_P681H, Spike_Y144del, NSP6_G107del, NSP3_A890D, Spike_D1118H, NSP6_F108del, NS8_Y73C, N_G204R, Spike_V70del, NS3_E194D, NSP12_P323L, Spike_D614G, N_D3L, Spike_S982A |
| 359 | CDR80 | EPI_ISL_1970393 | F | B.1.1.7 | G | Apr-21 | Spike_H69del, NS8_Q27stop, NSP3_T183I, NSP8_T145I, Spike_T716I, NS8_K68stop, NSP6_S106del, Spike_A570D, Spike_N501Y, NSP3_I1412T, NS8_R52I, Spike_P681H, Spike_Y144del, NSP6_G107del, NSP3_A890D, Spike_D1118H, NSP6_F108del, NS8_Y73C, Spike_V70del, NS3_E194D, NSP12_P323L, Spike_D614G, N_D3L |
| 360 | CDR81 | EPI_ISL_1970394 | F | B.1.1.7 | G | Apr-21 | Spike_H69del, NS8_Q27stop, NSP3_T183I, NSP8_T145I, NSP3_A890D, NSP6_G107del, NS8_K68stop, NSP6_S106del, Spike_A570D, Spike_D1118H, NSP6_F108del, NS8_Y73C, Spike_V70del, NS3_E194D, NSP3_I1412T, NS8_R52I, NSP12_P323L, Spike_P681H, Spike_D614G, Spike_Y144del, N_D3L, Spike_S982A |
| 361 | CDR82 | EPI_ISL_1970395 | F | B.1.1.7 | GR | Apr-21 | NS8_Q27stop, NSP3_T183I, NSP8_T145I, NS8_K68stop, NSP6_S106del, N_R203K, Spike_A570D, Spike_N501Y, NSP3_I1412T, NS8_R52I, Spike_Y144del, NSP6_G107del, NSP3_A890D, Spike_D1118H, NSP6_F108del, NS8_Y73C, N_G204R, NS3_E194D, NSP12_P323L, Spike_D614G, N_D3L, Spike_S982A, N_S235F |
| 362 | CDR83 | EPI_ISL_1970396 | F | B.1.1.7 | O | Apr-21 | Spike_H69del, NS8_Q27stop, NSP3_T183I, Spike_T716I, NSP6_S106del, N_R203K, NSP3_D110Y, Spike_A570D, NSP15_E260A, NSP3_R1297del, NSP3_I1412T, NS8_R52I, NSP8_S76F, Spike_P681H, Spike_Y144del, NSP3_V1298del, NSP6_G107del, NSP3_A890D, Spike_D1118H, NSP6_F108del, N_G204R, Spike_V70del, NSP3_S1296del, NSP12_P323L, Spike_D614G, N_D3L, Spike_S982A, N_S235F |
| 363 | CDR84 | EPI_ISL_1970397 | F | B.1.1.7 | GR | Apr-21 | Spike_H69del, NS8_Q27stop, NSP3_T183I, NS8_K68stop, NSP6_S106del, N_R203K, Spike_A570D, NSP3_I1412T, NS3_S180P, NS8_R52I, Spike_P681H, Spike_Y144del, NSP6_G107del, NSP3_A890D, Spike_D1118H, NSP6_F108del, N_G204R, Spike_V70del, NS3_E194D, NSP12_P323L, Spike_D614G, N_D3L, Spike_S982A |
| 364 | CDR85 | EPI_ISL_1970398 | F | B.1.1.7 | G | Apr-21 | Spike_H69del, NS8_Q27stop, NSP3_T183I, NSP8_T145I, Spike_T716I, NS8_K68stop, NSP6_S106del, Spike_A570D, NSP14_A320V, Spike_N501Y, NSP3_I1412T, NS8_R52I, Spike_Y144del, NSP2_Q383H, NSP6_G107del, NSP3_A890D, N_D402V, Spike_D1118H, NSP6_F108del, NS8_Y73C, Spike_V70del, NS3_E194D, NSP12_P323L, Spike_D614G, N_D3L, Spike_S982A |

| | | | | | | | |
|---|---|---|---|---|---|---|---|
| 365 | CDR87 | EPI_ISL_1970400 | F | B.1.1.7 | GR | Apr-21 | Spike_H69del, NS8_Q27stop, NSP3_T183I, NSP8_T145I, Spike_T716I, NS8_K68stop, NSP6_S106del, N_R203K, Spike_A570D, NSP3_I1412T, NS8_R52I, Spike_P681H, Spike_Y144del, NSP6_G107del, NSP3_A890D, Spike_D1118H, NSP6_F108del, NS8_Y73C, N_G204R, Spike_V70del, NS3_E194D, NSP12_P323L, Spike_D614G, N_D3L, Spike_S982A, N_S235F |
| 366 | CDR88 | EPI_ISL_1970401 | F | B.1.1.7 | GR | Apr-21 | Spike_H69del, Spike_V1264L, NSP3_T183I, NSP8_T145I, Spike_T716I, NS8_K68stop, NSP6_S106del, N_R203K, Spike_A570D, NSP3_I1412T, NS8_R52I, Spike_P681H, Spike_Y144del, NSP6_G107del, NSP3_A890D, Spike_D1118H, NSP6_F108del, NS8_Y73C, N_G204R, Spike_V70del, NS3_E194D, NSP12_P323L, Spike_D614G, N_D3L, Spike_S982A, N_S235F |
| 367 | CDR89 | EPI_ISL_1970402 | F | B.1.1.7 | GRY | Apr-21 | Spike_H69del, NS8_Q27stop, NSP3_T183I, NSP8_T145I, Spike_T716I, NSP6_S106del, N_R203K, Spike_N501Y, NSP3_I1412T, NS8_R52I, Spike_P681H, Spike_Y144del, NSP6_G107del, NSP6_F108del, NS8_Y73C, N_G204R, Spike_V70del, NS3_E194D, NSP12_P323L, Spike_D614G, N_D3L, Spike_S982A, N_S235F |
| 368 | CDR98 | EPI_ISL_1970410 | F | B.1.1.7 | G | Apr-21 | Spike_H69del, NS8_Q27stop, NSP3_T183I, NSP8_T145I, NSP6_L148F, Spike_T716I, NS8_K68stop, NSP6_S106del, Spike_A570D, Spike_N501Y, NSP3_I1412T, Spike_P681H, Spike_Y144del, NSP6_G107del, NSP3_A890D, Spike_D1118H, NSP6_F108del, NS8_Y73C, Spike_V70del, NS3_E194D, NSP12_P323L, Spike_D614G, N_D3L, Spike_S982A |
| 369 | CDR99 | EPI_ISL_1970411 | F | B.1.1.7 | GR | Apr-21 | Spike_H69del, NSP3_T183I, Spike_T716I, NSP6_S106del, N_R203K, NSP4_A231V, Spike_A570D, NS3_T89I, NSP13_L581F, NSP3_I1412T, Spike_P681H, Spike_Y144del, NSP12_P227L, NSP3_P67L, NSP6_G107del, NSP3_A890D, Spike_D1118H, NSP6_F108del, N_G204R, NSP3_T275A, Spike_V70del, NSP5_A193V, NSP12_P323L, NSP14_P451S, Spike_D614G, N_D3L, Spike_S982A, N_S235F |
| 370 | CDR86 | EPI_ISL_1970399 | F | B.1.525 | G | Apr-21 | Spike_H69del, NSP12_P323F, M_I82T, Spike_A67V, N_A12G, NSP3_T1189I, NSP6_G107del, E_L21F, NSP2_K443Q, NSP6_S106del, Spike_E484K, N_T205I, NSP6_F108del, Spike_Q52R, Spike_V70del, NS3_S92L, Spike_D614G, Spike_Y144del, Spike_F888L, Spike_Q677H |
| 371 | CDR101 | EPI_ISL_1970351 | F | B.1.1.7 | G | Apr-21 | Spike_H69del, NS8_Q27stop, NSP3_T183I, NSP8_T145I, Spike_T716I, NS8_K68stop, NSP6_S106del, Spike_A570D, NSP14_A320V, Spike_N501Y, NSP3_I1412T, NS8_R52I, Spike_P681H, Spike_Y144del, NSP6_G107del, NSP3_A890D, Spike_E748V, Spike_D1118H, NSP6_F108del, NS8_Y73C, Spike_V70del, NS3_E194D, NSP12_P323L, Spike_D614G, N_D3L, Spike_S982A |
| 372 | CDR102 | EPI_ISL_1972231 | F | B.1.1.7 | GRY | Apr-21 | Spike_H69del, NS8_Q27stop, NSP3_T183I, E_Y57F, Spike_T716I, NSP3_W1196C, NSP6_S106del, N_R203K, NSP15_E260A, Spike_N501Y, NSP3_R1297del, NSP3_I1412T, NS8_R52I, NSP8_S76F, Spike_P681H, Spike_S151del, NSP3_V1298del, NSP6_G107del, NSP3_A890D, Spike_D1118H, NSP6_F108del, E_F56A, N_G204R, Spike_V70del, NSP3_S1296del, NSP12_P323L, Spike_D614G, N_D3L, Spike_S982A, N_S235F |

| | | | | | | | |
|---|---|---|---|---|---|---|---|
| 373 | CMC94262 | EPI_ISL_1970415 | F | B.1.1.7 | GRY | Apr-21 | Spike_H69del, NS8_Q27stop, NSP3_T183I, NSP8_T145I, Spike_T716I, NS8_K68stop, NSP6_S106del, N_R203K, Spike_A570D, Spike_N501Y, NSP3_I1412T, NS8_R52I, Spike_P681H, Spike_Y144del, NSP6_G107del, NSP3_A890D, NSP5_P96L, NSP3_G145D, Spike_D1118H, NSP6_F108del, NS8_Y73C, N_G204R, Spike_V70del, NS3_E194D, NSP12_P323L, Spike_D614G, N_D3L, Spike_S982A |
