## Supplementary table 2 for "Genomic and epidemiological analysis of SARS-CoV-2 viruses in Sri Lanka"

**Supplementary Table 2. Amino acid mutation counts of 231 sequences belonging to B.1.411 lineage.**

| Protein | AA_Substitution | Count | Mutation_ID |
|---|---|---|---|
| Spike | D614G | 228 | Spike_D614G |
| NSP12 | M666I | 224 | NSP12_M666I |
| Spike | H1159Y | 215 | Spike_H1159Y |
| NSP6 | L37F | 205 | NSP6_L37F |
| NSP2 | T166I | 202 | NSP2_T166I |
| NSP12 | P323L | 201 | NSP12_P323L |
| N | T205I | 198 | N_T205I |
| NS8 | Q18stop | 164 | NS8_Q18stop |
| NSP12 | D445G | 154 | NSP12_D445G |
| NS3 | Q57H | 147 | NS3_Q57H |
| NSP2 | T85I | 109 | NSP2_T85I |
| NSP3 | P654S | 19 | NSP3_P654S |
| Spike | A684V | 18 | Spike_A684V |
| NS3 | T151S | 16 | NS3_T151S |
| Spike | T676A | 13 | Spike_T676A |
| NSP6 | C221F | 10 | NSP6_C221F |
| NSP5 | K90R | 9 | NSP5_K90R |
| Spike | M1229I | 9 | Spike_M1229I |
| NS3 | R30H | 8 | NS3_R30H |
| NS3 | L108F | 8 | NS3_L108F |
| NS8 | R52I | 8 | NS8_R52I |
| Spike | S98P | 8 | Spike_S98P |
| N | T265I | 7 | N_T265I |
| NSP16 | R86K | 7 | NSP16_R86K |
| Spike | S640F | 7 | Spike_S640F |
| NS8 | F120V | 6 | NS8_F120V |
| N | K373R | 6 | N_K373R |
| NS8 | I121L | 6 | NS8_I121L |
| NSP6 | E195D | 6 | NSP6_E195D |
| NSP12 | V720I | 6 | NSP12_V720I |
| NSP9 | M101I | 6 | NSP9_M101I |
| Spike | T716I | 5 | Spike_T716I |
| Spike | T33S | 5 | Spike_T33S |
| NSP15 | T48I | 5 | NSP15_T48I |
| NSP14 | S450I | 5 | NSP14_S450I |
| NSP7 | T81I | 5 | NSP7_T81I |
| NSP12 | T85I | 5 | NSP12_T85I |
| Spike | V70del | 5 | Spike_V70del |
| Spike | H69del | 5 | Spike_H69del |
| NSP13 | H290Y | 5 | NSP13_H290Y |
| NSP3 | S1670F | 4 | NSP3_S1670F |
| Spike | N679K | 4 | Spike_N679K |
| NSP13 | L581F | 4 | NSP13_L581F |
| NS8 | W45C | 4 | NS8_W45C |
| NSP14 | P203L | 4 | NSP14_P203L |
| NS8 | P93L | 4 | NS8_P93L |
| Spike | D287N | 4 | Spike_D287N |
| NSP2 | K337E | 4 | NSP2_K337E |

| | | | |
|---|---|---|---|
| NSP3 | K1804N | 4 | NSP3_K1804N |
| NSP16 | Q3L | 4 | NSP16_Q3L |
| Spike | D178G | 4 | Spike_D178G |
| NSP2 | G392R | 4 | NSP2_G392R |
| Spike | G1124V | 4 | Spike_G1124V |
| NSP14 | S134F | 4 | NSP14_S134F |
| NSP9 | T24I | 4 | NSP9_T24I |
| Spike | N439K | 4 | Spike_N439K |
| NSP13 | A505P | 4 | NSP13_A505P |
| NSP13 | A598S | 4 | NSP13_A598S |
| NSP3 | I1683T | 4 | NSP3_I1683T |
| NSP3 | K429N | 3 | NSP3_K429N |
| NSP14 | K349N | 3 | NSP14_K349N |
| NSP12 | Q292H | 3 | NSP12_Q292H |
| Spike | L5F | 3 | Spike_L5F |
| NSP3 | A667T | 3 | NSP3_A667T |
| NSP3 | A690V | 3 | NSP3_A690V |
| NSP2 | H208Y | 3 | NSP2_H208Y |
| NSP3 | H290Y | 3 | NSP3_H290Y |
| N | D144Y | 3 | N_D144Y |
| NS3 | S171L | 3 | NS3_S171L |
| NSP13 | S80G | 3 | NSP13_S80G |
| NSP2 | T388I | 3 | NSP2_T388I |
| NSP8 | T148I | 3 | NSP8_T148I |
| NS3 | V13L | 3 | NS3_V13L |
| NSP3 | Q1884H | 3 | NSP3_Q1884H |
| NS3 | L106R | 3 | NS3_L106R |
| NSP3 | A488V | 3 | NSP3_A488V |
| Spike | A222V | 3 | Spike_A222V |
| N | A134V | 3 | N_A134V |
| NSP3 | A1311V | 3 | NSP3_A1311V |
| NSP3 | S1265del | 2 | NSP3_S1265del |
| NSP3 | T204I | 2 | NSP3_T204I |
| NSP3 | T1072I | 2 | NSP3_T1072I |
| NSP2 | E201A | 2 | NSP2_E201A |
| NS3 | E102Q | 2 | NS3_E102Q |
| NSP6 | V149F | 2 | NSP6_V149F |
| NS3 | V255del | 2 | NS3_V255del |
| NS3 | V202L | 2 | NS3_V202L |
| NSP3 | L1328F | 2 | NSP3_L1328F |
| NSP3 | L1266I | 2 | NSP3_L1266I |
| NSP15 | A94T | 2 | NSP15_A94T |
| NS3 | W193L | 2 | NS3_W193L |
| NSP4 | I383F | 2 | NSP4_I383F |
| NSP1 | G30S | 2 | NSP1_G30S |
| NSP2 | G147D | 2 | NSP2_G147D |
| NS3 | T175I | 2 | NS3_T175I |
| N | T362I | 2 | N_T362I |
| NSP2 | T497K | 2 | NSP2_T497K |
| Spike | T76I | 2 | Spike_T76I |
| NSP3 | K529R | 2 | NSP3_K529R |

| | | | |
|---|---|---|---|
| NSP12 | E254D | 2 | NSP12_E254D |
| NS8 | V62L | 2 | NS8_V62L |
| NSP9 | V76A | 2 | NSP9_V76A |
| NSP7 | V58A | 2 | NSP7_V58A |
| NS3 | L41F | 2 | NS3_L41F |
| NS3 | W131C | 2 | NS3_W131C |
| NSP13 | D260Y | 2 | NSP13_D260Y |
| N | D402Y | 2 | N_D402Y |
| NS8 | I58V | 2 | NS8_I58V |
| NSP3 | G17C | 2 | NSP3_G17C |
| NSP3 | M1556L | 2 | NSP3_M1556L |
| NSP3 | L1096I | 2 | NSP3_L1096I |
| NSP2 | L71F | 2 | NSP2_L71F |
| NS7a | I3F | 2 | NS7a_I3F |
| NSP3 | S1675I | 1 | NSP3_S1675I |
| NSP14 | S503L | 1 | NSP14_S503L |
| NSP3 | S284C | 1 | NSP3_S284C |
| E | S55F | 1 | E_S55F |
| M | S4F | 1 | M_S4F |
| NSP10 | T12I | 1 | NSP10_T12I |
| NSP3 | T423I | 1 | NSP3_T423I |
| N | K387R | 1 | N_K387R |
| NSP3 | K945N | 1 | NSP3_K945N |
| NS8 | E59stop | 1 | NS8_E59stop |
| Spike | E484K | 1 | Spike_E484K |
| NSP1 | E36K | 1 | NSP1_E36K |
| NSP12 | Y546C | 1 | NSP12_Y546C |
| Spike | V213L | 1 | Spike_V213L |
| NS3 | V50I | 1 | NS3_V50I |
| NSP5 | V261F | 1 | NSP5_V261F |
| Spike | V1122L | 1 | Spike_V1122L |
| Spike | V367F | 1 | Spike_V367F |
| NS3 | V55I | 1 | NS3_V55I |
| Spike | V1040F | 1 | Spike_V1040F |
| NSP15 | V320L | 1 | NSP15_V320L |
| NSP16 | Q238H | 1 | NSP16_Q238H |
| Spike | M1237I | 1 | Spike_M1237I |
| NS8 | C83S | 1 | NS8_C83S |
| E | C40L | 1 | E_C40L |
| NS7a | L102P | 1 | NS7a_L102P |
| NSP6 | L125F | 1 | NSP6_L125F |
| NS3 | L53F | 1 | NS3_L53F |
| NS8 | L95F | 1 | NS8_L95F |
| E | L39C | 1 | E_L39C |
| NSP2 | A159S | 1 | NSP2_A159S |
| NS3 | A59T | 1 | NS3_A59T |
| NSP3 | A338H | 1 | NSP3_A338H |
| Spike | A879T | 1 | Spike_A879T |
| E | A41L | 1 | E_A41L |
| NSP12 | W617F | 1 | NSP12_W617F |
| NSP3 | P822L | 1 | NSP3_P822L |

| | | | |
|---|---|---|---|
| NS3 | P262S | 1 | NS3_P262S |
| NSP14 | P140S | 1 | NSP14_P140S |
| NSP3 | P2L | 1 | NSP3_P2L |
| NSP3 | ins336DHNY | 1 | NSP3_ins336DHNY |
| NSP12 | ins617LR | 1 | NSP12_ins617LR |
| NSP14 | H455Y | 1 | NSP14_H455Y |
| NS3 | D210Y | 1 | NS3_D210Y |
| N | D144N | 1 | N_D144N |
| NSP12 | D618N | 1 | NSP12_D618N |
| NSP12 | D284N | 1 | NSP12_D284N |
| NSP3 | D174Y | 1 | NSP3_D174Y |
| Spike | G261V | 1 | Spike_G261V |
| NSP3 | G777V | 1 | NSP3_G777V |
| NSP3 | G145C | 1 | NSP3_G145C |
| NSP1 | G137S | 1 | NSP1_G137S |
| Spike | G1167R | 1 | Spike_G1167R |
| NSP3 | G337S | 1 | NSP3_G337S |
| NSP14 | G481S | 1 | NSP14_G481S |
| NSP9 | G37R | 1 | NSP9_G37R |
| NSP15 | S261L | 1 | NSP15_S261L |
| N | S201N | 1 | N_S201N |
| NSP2 | S378F | 1 | NSP2_S378F |
| NSP15 | S308Y | 1 | NSP15_S308Y |
| NS3 | S92L | 1 | NS3_S92L |
| NSP6 | F35L | 1 | NSP6_F35L |
| NSP6 | F34V | 1 | NSP6_F34V |
| NSP14 | F217del | 1 | NSP14_F217del |
| Spike | F1121L | 1 | Spike_F1121L |
| Spike | T1238S | 1 | Spike_T1238S |
| Spike | T859I | 1 | Spike_T859I |
| NSP2 | T170I | 1 | NSP2_T170I |
| NS8 | T26I | 1 | NS8_T26I |
| NSP3 | T1303I | 1 | NSP3_T1303I |
| NSP6 | T29P | 1 | NSP6_T29P |
| NSP14 | T215K | 1 | NSP14_T215K |
| NSP6 | T172A | 1 | NSP6_T172A |
| NSP3 | N922S | 1 | NSP3_N922S |
| NSP12 | N9Y | 1 | NSP12_N9Y |
| Spike | N751K | 1 | Spike_N751K |
| NS3 | K67R | 1 | NS3_K67R |
| NSP2 | K456R | 1 | NSP2_K456R |
| NS3 | E102K | 1 | NS3_E102K |
| Spike | E180G | 1 | Spike_E180G |
| Spike | E154K | 1 | Spike_E154K |
| Spike | E748D | 1 | Spike_E748D |
| NS3 | Y107C | 1 | NS3_Y107C |
| NSP14 | Y361C | 1 | NSP14_Y361C |
| NSP2 | V308L | 1 | NSP2_V308L |
| NSP10 | V7L | 1 | NSP10_V7L |
| NSP3 | V929I | 1 | NSP3_V929I |
| NSP13 | V266L | 1 | NSP13_V266L |

| | | | |
|---|---|---|---|
| NSP3 | V267F | 1 | NSP3_V267F |
| Spike | V622F | 1 | Spike_V622F |
| NSP1 | V84del | 1 | NSP1_V84del |
| NS3 | Q185H | 1 | NS3_Q185H |
| NSP6 | Q30E | 1 | NSP6_Q30E |
| NSP12 | Q822H | 1 | NSP12_Q822H |
| NSP3 | Q995H | 1 | NSP3_Q995H |
| NSP1 | M85V | 1 | NSP1_M85V |
| NSP14 | C208del | 1 | NSP14_C208del |
| E | C40del | 1 | E_C40del |
| NSP10 | C41S | 1 | NSP10_C41S |
| NSP14 | C210del | 1 | NSP14_C210del |
| Spike | L585F | 1 | Spike_L585F |
| NSP3 | L862F | 1 | NSP3_L862F |
| NSP14 | L209del | 1 | NSP14_L209del |
| Spike | L18F | 1 | Spike_L18F |
| NSP3 | L72F | 1 | NSP3_L72F |
| NSP12 | L8C | 1 | NSP12_L8C |
| E | L39del | 1 | E_L39del |
| NSP6 | L33M | 1 | NSP6_L33M |
| NS7b | L32F | 1 | NS7b_L32F |
| NSP3 | A1321V | 1 | NSP3_A1321V |
| NSP3 | A1105T | 1 | NSP3_A1105T |
| NSP3 | A1280V | 1 | NSP3_A1280V |
| NSP8 | A21V | 1 | NSP8_A21V |
| NS3 | A23S | 1 | NS3_A23S |
| E | A41S | 1 | E_A41S |
| M | A2S | 1 | M_A2S |
| NSP6 | W31Y | 1 | NSP6_W31Y |
| NSP16 | P236L | 1 | NSP16_P236L |
| NSP3 | P1228S | 1 | NSP3_P1228S |
| NSP3 | P395L | 1 | NSP3_P395L |
| NSP13 | P326L | 1 | NSP13_P326L |
| NSP12 | ins9stop | 1 | NSP12_ins9stop |
| NSP6 | ins35VF | 1 | NSP6_ins35VF |
| NSP6 | ins171MTARTVYDDG | 1 | NSP6_ins171MTARTVYDDG |
| Spike | H1101Y | 1 | Spike_H1101Y |
| NSP3 | H1880Y | 1 | NSP3_H1880Y |
| NSP1 | H83del | 1 | NSP1_H83del |
| Spike | D936Y | 1 | Spike_D936Y |
| Spike | D253N | 1 | Spike_D253N |
| NSP14 | D496Y | 1 | NSP14_D496Y |
| Spike | D80Y | 1 | Spike_D80Y |
| NSP14 | D211del | 1 | NSP14_D211del |
| NSP16 | D26H | 1 | NSP16_D26H |
| NSP12 | D135Y | 1 | NSP12_D135Y |
| NS3 | D199Y | 1 | NS3_D199Y |
| NSP12 | R10C | 1 | NSP12_R10C |
| NSP14 | R212H | 1 | NSP14_R212H |
| N | R209K | 1 | N_R209K |
| NSP14 | R213H | 1 | NSP14_R213H |

| | | | |
|---|---|---|---|
| NSP3 | I967T | 1 | NSP3_I967T |
| NSP13 | I333V | 1 | NSP13_I333V |
| NSP1 | G82del | 1 | NSP1_G82del |
| Spike | F186L | 1 | Spike_F186L |
| NSP13 | F373Y | 1 | NSP13_F373Y |
| Spike | F92H | 1 | Spike_F92H |
| NSP3 | S1206L | 1 | NSP3_S1206L |
| NSP14 | S255I | 1 | NSP14_S255I |
| NSP2 | S211H | 1 | NSP2_S211H |
| Spike | S94P | 1 | Spike_S94P |
| NSP3 | S1443F | 1 | NSP3_S1443F |
| Spike | T63A | 1 | Spike_T63A |
| NS6 | N39T | 1 | NS6_N39T |
| NSP3 | N1680D | 1 | NSP3_N1680D |
| NSP16 | N235R | 1 | NSP16_N235R |
| M | K15M | 1 | M_K15M |
| M | K14I | 1 | M_K14I |
| NS3 | Y264H | 1 | NS3_Y264H |
| NSP3 | Y519N | 1 | NSP3_Y519N |
| NSP2 | E210D | 1 | NSP2_E210D |
| NS8 | E106stop | 1 | NS8_E106stop |
| NSP3 | V1936S | 1 | NSP3_V1936S |
| Spike | V1176F | 1 | Spike_V1176F |
| N | V72I | 1 | N_V72I |
| NSP10 | V108del | 1 | NSP10_V108del |
| NSP3 | V1935L | 1 | NSP3_V1935L |
| NSP2 | V447F | 1 | NSP2_V447F |
| N | V270L | 1 | N_V270L |
| NSP13 | V371F | 1 | NSP13_V371F |
| NSP13 | V372A | 1 | NSP13_V372A |
| NSP5 | Q306R | 1 | NSP5_Q306R |
| Spike | Q14H | 1 | Spike_Q14H |
| NS8 | Q72H | 1 | NS8_Q72H |
| N | M210V | 1 | N_M210V |
| NSP2 | M404V | 1 | NSP2_M404V |
| N | M234V | 1 | N_M234V |
| NSP12 | M380L | 1 | NSP12_M380L |
| NSP4 | C296F | 1 | NSP4_C296F |
| Spike | L841I | 1 | Spike_L841I |
| NSP2 | L410F | 1 | NSP2_L410F |
| M | L16Q | 1 | M_L16Q |
| Spike | L1063F | 1 | Spike_L1063F |
| NSP4 | L353F | 1 | NSP4_L353F |
| NSP2 | L213S | 1 | NSP2_L213S |
| NSP2 | L550I | 1 | NSP2_L550I |
| NS7a | A79V | 1 | NS7a_A79V |
| NSP7 | A80V | 1 | NSP7_A80V |
| NS7b | A15S | 1 | NS7b_A15S |
| NSP14 | A425V | 1 | NSP14_A425V |
| Spike | A783S | 1 | Spike_A783S |
| NSP4 | A380V | 1 | NSP4_A380V |

| | | | |
|---|---|---|---|
| NSP4 | A231V | 1 | NSP4_A231V |
| Spike | A93Y | 1 | Spike_A93Y |
| NSP3 | P985L | 1 | NSP3_P985L |
| NSP3 | P1261S | 1 | NSP3_P1261S |
| NSP13 | P504S | 1 | NSP13_P504S |
| Spike | P1263L | 1 | Spike_P1263L |
| NSP10 | P107del | 1 | NSP10_P107del |
| N | P199S | 1 | N_P199S |
| NS7a | P99S | 1 | NS7a_P99S |
| NSP16 | ins234MstopM | 1 | NSP16_ins234MstopM |
| NSP13 | D369E | 1 | NSP13_D369E |
| N | D128Y | 1 | N_D128Y |
| NSP3 | I617V | 1 | NSP3_I617V |
| NSP2 | R222C | 1 | NSP2_R222C |
| NS3 | G174V | 1 | NS3_G174V |
| NSP2 | G212T | 1 | NSP2_G212T |
| NS7a | G38stop | 1 | NS7a_G38stop |
